# Hip joint space width is causally related to hip osteoarthritis risk via distinct protective and susceptibility mechanisms: findings from a genome-wide association study meta-analysis

**DOI:** 10.1101/2023.03.01.23286618

**Authors:** Monika Frysz, Benjamin G. Faber, Cindy G. Boer, Daniel S. Evans, Raja Ebsim, Kaitlyn A. Flynn, Mischa Lundberg, Lorraine Southam, April Hartley, Fiona R. Saunders, Claudia Lindner, Jennifer S. Gregory, Richard M. Aspden, Nancy E. Lane, Nicholas C. Harvey, David M. Evans, Eleftheria Zeggini, George Davey Smith, Timothy Cootes, Joyce Van Meurs, John P. Kemp, Jonathan H. Tobias

## Abstract

**Objective:** Minimum joint space width (mJSW) from 2-dimensional images provides a proxy for cartilage thickness. This study aimed to conduct a genome-wide association study (GWAS) of mJSW to (i) identify new genetic determinants of mJSW and use them to (ii) examine causal effects of mJSW on hip osteoarthritis (HOA) risk.

**Methods:** GWAS meta-analysis of hip mJSW derived from plain X-rays (four cohorts) or DXA (one cohort) was performed, stratified by sex and adjusted for age and ancestry principal components. Mendelian randomisation (MR) and cluster analyses were used to examine causal effect of mJSW on HOA.

**Results:** 50,745 individuals were included in the meta-analysis. 42 SNPs, which mapped to 39 loci (35 novel), were identified. Mendelian randomisation (MR) revealed little evidence of a causal effect of mJSW on HOA (*β*_*IVW*_ -0.01 [95% CI -0.19, 0.17]). However, MR-Clust analysis suggested the null MR estimates reflected the net effect of two distinct causal mechanisms cancelling each other out, one of which was protective, whereas the other increased HOA susceptibility. For the latter mechanism, all loci were positively associated with height, suggesting mechanisms leading to greater height and mJSW increase the risk of HOA in later life.

**Conclusions:** GWAS and MR analyses suggested one group of mJSW loci reduces HOA risk via increased mJSW, suggesting possible utility as targets for chondroprotective therapies. The second group of mJSW loci increased HOA risk, despite increasing mJSW, but were also positively related to height, suggesting they contribute to mJSW and HOA risk via a growth-related mechanism.

## Introduction

Hip osteoarthritis (HOA) is the commonest cause of pain and loss of function of the hip worldwide (1). It is a disease of the whole joint with multiple biological pathways implicated in its pathogenesis, involving cartilage, bone and synovium (2). The prevalence of HOA is approximately 10% and is predicted to increase (1, 3). Currently there are no known means of preventing disease and/or symptomatic progression, leaving total hip replacement (THR) as the treatment of choice for those with end-stage disease. As a result, HOA costs European countries over €400 billion/year in both direct and indirect healthcare costs illustrating its substantial health and economic burden (4). A better understanding of the pathogenesis of HOA may uncover new opportunities for treatment, prevention and early diagnosis.

The heritability of HOA is estimated at 50%, indicating a considerable genetic role in its pathogenesis (5, 6). In an attempt to identify specific genetic associations, large-scale genome-wide association studies (GWAS) have been used to identify genes and their biological pathways which might offer potential treatment targets (7, 8). To date, 45 independent genetic loci have been associated with HOA, but the underlying genetic pathways causing disease remain largely unclear (9). A more focused GWAS, which investigates heritable components of HOA (endophenotypes), may shed light on these mechanisms. One such endophenotype is minimum joint space width (mJSW), which is defined as the narrowest distance across the joint and acts as proxy for cartilage thickness, cartilage loss being a key feature of HOA (10). Therefore, genetic factors associated with mJSW might help to identify pathways involved in cartilage homeostasis. As well as giving more insight into biology, using a continuous endophenotype such as mJSW increases statistical power for GWAS compared with a case-control definition of end-stage disease.

An earlier GWAS found four independent loci associated with mJSW obtained from antero-posterior (AP) radiographs, many of which showed nominal associations with HOA (11). Larger sample sizes and updated genotype reference panels provide the opportunity for comprehensive characterization of mJSW genetic architecture. These data can also provide genetic instruments for Mendelian randomisation (MR) to test causal associations between exposures and outcomes (12), such as mJSW and HOA risk. The UK Biobank study (UKB) has recently conducted over 40,000 high-resolution dual-energy X-ray absorptiometry (DXA) scans of the hip that have been automatically annotated for mJSW (3). The present study aimed to conduct a GWAS meta-analysis of hip mJSW combining X-ray and DXA cohorts to maximize the study power to identify novel loci, and then explore the genetic architecture of mJSW and its relationship with HOA. Subsequently, we aimed to evaluate causal effects of mJSW on HOA risk using MR and cluster analyses, to allow for the possibility that distinct sets of SNPs associated with mJSW map to directionally opposite causal pathways (13).

## Methods

### Cohort descriptions

GWAS cohorts comprised the UKB, The Rotterdam Study (RS) I&II, Osteoporotic Fractures in Men (MrOS) Study and Study of Osteoporotic Fractures (SOF). mJSW was measured automatically in UKB and manually in RS, MrOS and SOF (see Supplementary Methods).

UKB study (application number 17295) is overseen by the Ethics Advisory Committee and received approval from the National Information Governance Board for Health and Social Care and Northwest Multi-Centre Research Ethics Committee (11/NW/0382), all participants provided informed consent for this study. The Rotterdam Study has been approved by the Medical Ethics Committee of the Erasmus MC (registration number MEC 02.1015) and by the Dutch Ministry of Health, Welfare and Sport (Population Screening Act WBO, license number 1071272-159521-PG). The Rotterdam Study has been entered into the Netherlands National Trial Register (NTR; www.trialregister.nl) and into the WHO International Clinical Trials Registry Platform (ICTRP; www.who.int/ictrp/network/primary/en/) under shared catalogue number NTR6831. All participants provided written informed consent to participate in the study and to have their information obtained from treating physicians.

### Genome-wide association study and meta-analysis

GWAS for mJSW were conducted separately in UKB, RS I&II, SOF and MrOS. In each cohort, mJSW was stratified by sex and adjusted for age, ancestry principal components, and in addition study site in the case of MrOS and SOF. Given potential relationships between mJSW and height, a further GWAS was performed including height adjustment for each cohort. Residuals resulting from female and male analysis were standardised to mean=0, SD=1, and then combined into a single outcome for GWAS. UKB used a linear mixed model for GWAS, implemented in BOLT-LMM (v2.3) (14), SOF, MrOS used an OLS linear regression model implemented in PLINK (15) and RS1 and RS2 used RVtests (5). RS I&II were imputed to Haplotype Reference Consortium (HRC v.1.1), UK Biobank release V3 was imputed to 3 reference panels (UK10K, 1000 Genomes and HRC) and SOF and MrOS were imputed to 1000 Genomes. All cohorts used the hg19 build. A subsequent inverse variance weighted fixed effects meta-analysis was performed with METAL (16) (see Supplementary Methods).

### Linkage disequilibrium score regression and genome-wide conditional and joint complex trait analysis (GCTA-COJO)

Linkage disequilibrium (LD) score regression (LDSC) v1.0.1 was used to estimate SNP heritability, and the genetic correlation between mJSW and several other traits, including HOA, height, and body mass index (BMI) (see Supplementary Methods) (9, 17). In addition, the genetic correlation between mJSW_DXA_ and mJSW_X-ray_ was examined. A European based LD reference panel was used, and analysis was limited to HapMap3 SNPs (therefore excluding major histocompatibility regions) (17). Conditional and joint analysis (GCTA-COJO) was performed in conjunction with a UKB reference panel to identify statistically independent mJSW associated signals (18).

### Mendelian randomisation and MR Cluster Analysis

The conditionally independent mJSW lead SNPs were used as genetic instruments for MR analyses to investigate the causal effect of mJSW on HOA, using the TwoSampleMR package v0.5.6 in R (19). The HOA GWAS was a meta-analysis combining the latest genetics of osteoarthritis consortium HOA GWAS without UKB (9) and an updated UKB HOA GWAS removing those individuals with mJSW measures to avoid sample overlap (see Supplementary Methods). Steiger filtering was applied to demonstrate the exposure instruments were upstream of the outcome. Inverse variance weighted (IVW) analysis was used as the primary method, with MR Egger, weighted median, simple mode and weighted mode approaches as sensitivity analyses (12). MR-Clust was applied in relation to HOA to group variants into distinct groups with similar causal estimates (13). This method, which may help to identify different causal mechanisms underlying HOA, is used when heterogeneity in causal effect estimates for a complex trait is observed, and different biological mechanisms are suspected. Two sample MR was then used to quantify cluster specific effects on HOA and height.

### Gene prioritisation and Downstream analyses

Initially, the independent mJSW lead SNPs were looked-up in a previous GWAS of height and BMI in UKB available via the IEU open GWAS project (20) and HOA. SNPs were prioritised based on MR-Clust results and a look-up in previous height and HOA summary statistics. In these fine mapping analyses that used the coloc R package, we compared 100kb regions on either side of the lead mJSW SNP in the mJSW and HOA GWAS to look for shared signals (21). Then generalised gene-set analysis of GWAS data (MAGMA v1.08) (22) was implemented in Functional Mapping and annotation of GWAS (FUMA) tool (23). Briefly, SNPs were mapped to the protein coding genes using default settings (SNP-wise (mean) model for gene test) and gene-set analysis was performed using 10894 gene sets obtained from MsigDB v5.2. In addition, the list of mapped genes was annotated for overlapping gene ontology biological processes genes using PANTHER (24). Subsequently, the expression quantitative trait loci (eQTL) database GTEx was searched for each leading SNP to identify cis-acting effects, with cultured fibroblasts considered the most relevant tissue. LocusFocus was used to conduct Bayesian colocalisation with all expressed genes over 100kb either side of the sentinel SNP (21, 25, 26). To further identify which cis-genes share the same causal variants, we used colocalisation to look at eQTL data assessed on highly degraded (diseased) and less degraded (healthy) cartilage, and synovial tissue retrieved following knee and hip joint replacements (27). We considered a SNP to colocalise with an eQTL if the posterior probability (PP) was >80%. In addition, regulatory elements of non-coding human genome were identified using RegulomeDB (28).

## Results

### Genome wide association analysis

We conducted a GWAS meta-analysis of hip mJSW in 50,745 participants from 5 cohorts, of whom 24,429 were males and 26,316 females with a mean age of 65.1 years (range 45-97 years), height of 169.7 cm (135-204 cm), weight of 75.5 kg (34-171 kg) and mJSW of 3.05 mm (0.0-7.4 mm), (Supplementary Table 1). Following conditional analyses, 42 independent SNPs were identified at genome-wide significance (*P* ≤5·0×10^−8^), together accounting for 4.6% of mJSW variance (Table 1, Figure 1) (11). The identified SNPs mapped to 39 loci, of which 35 were novel (defined as >1MB from previously reported variants (11)). mJSW SNP heritability (h^2^) was 0.20 (95% CI 0.16, 0.25), and there was moderate genomic inflation (λ=1.11; UKB λ= 1.10, MrOS 1.02, SOF 1.00, RS1 1.01, RS2 1.00). However, the intercept from LDSC, and the ratio attenuation statistic (Intercept=1.01 [Standard error=0.01] / RPS=0.28 [0.15]) suggested that most of the inflation reflected polygenicity rather than confounding due to population stratification or relatedness (Supplementary Figure 2). Equivalent results were obtained in a further GWAS following height adjustment (Supplementary Figure 3).

**Table 1.**
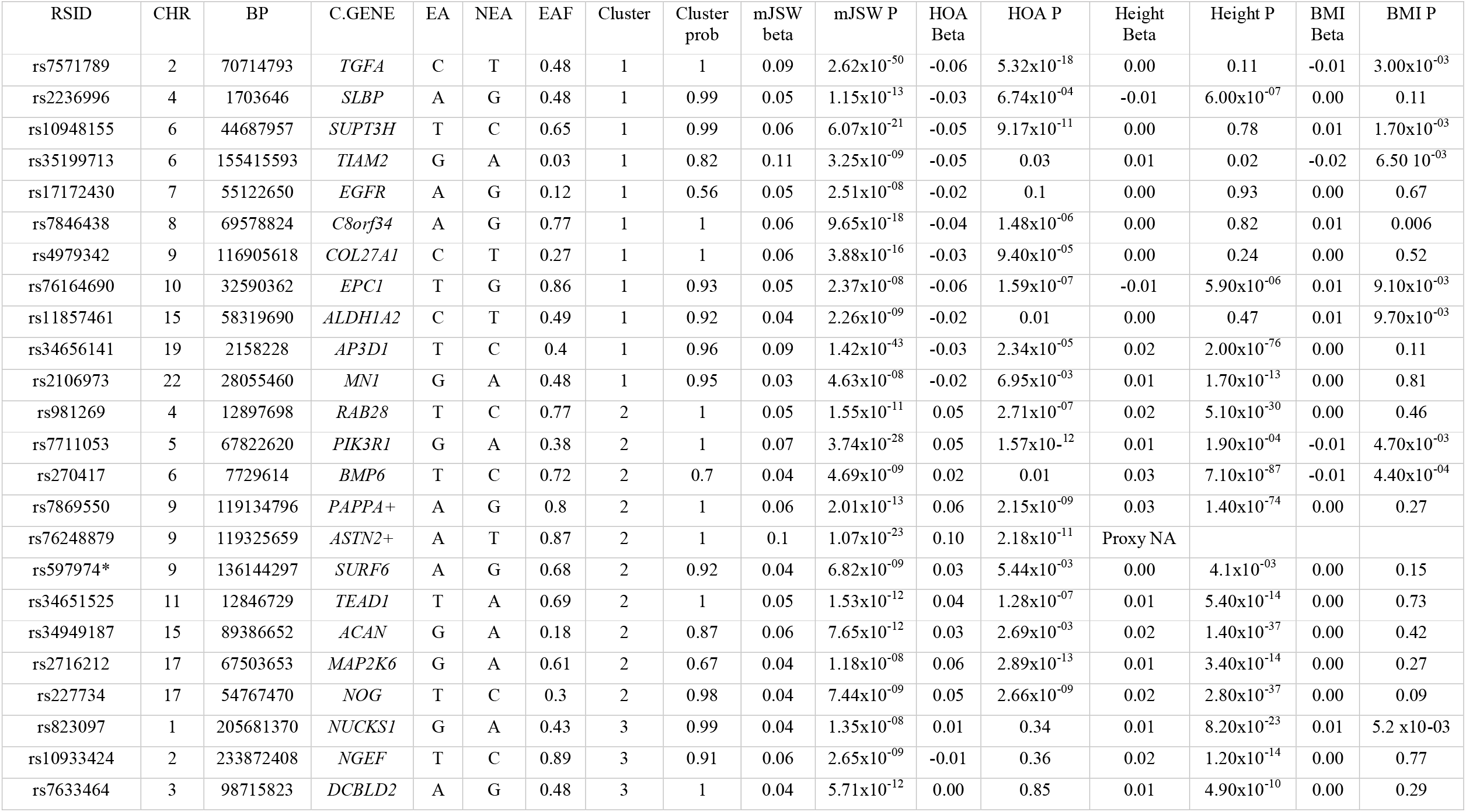
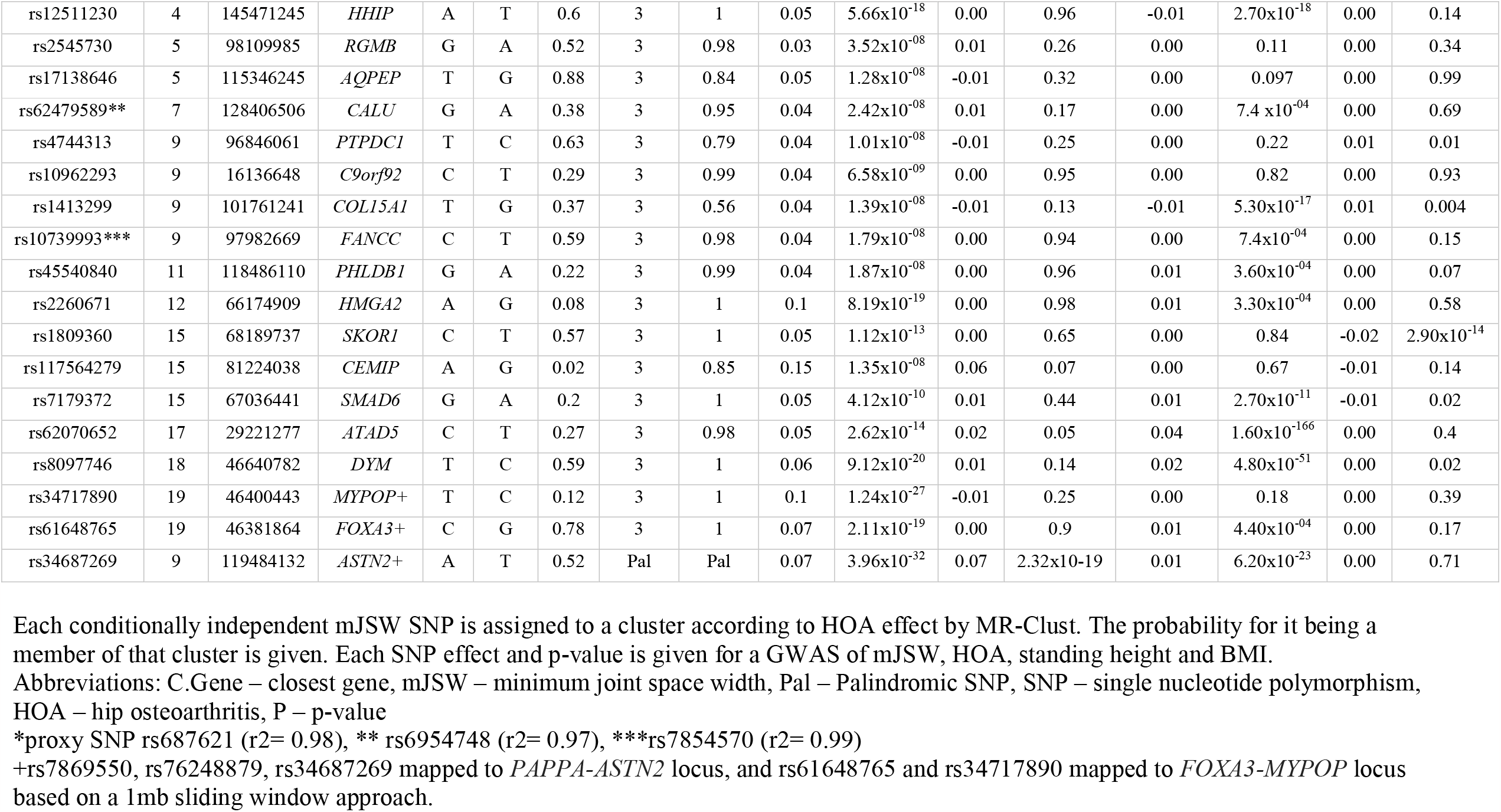
Conditionally independent minimum joint space width single nucleotide polymorphisms, and their associations with height, BMI, and HOA risk.

**Figure 1.**
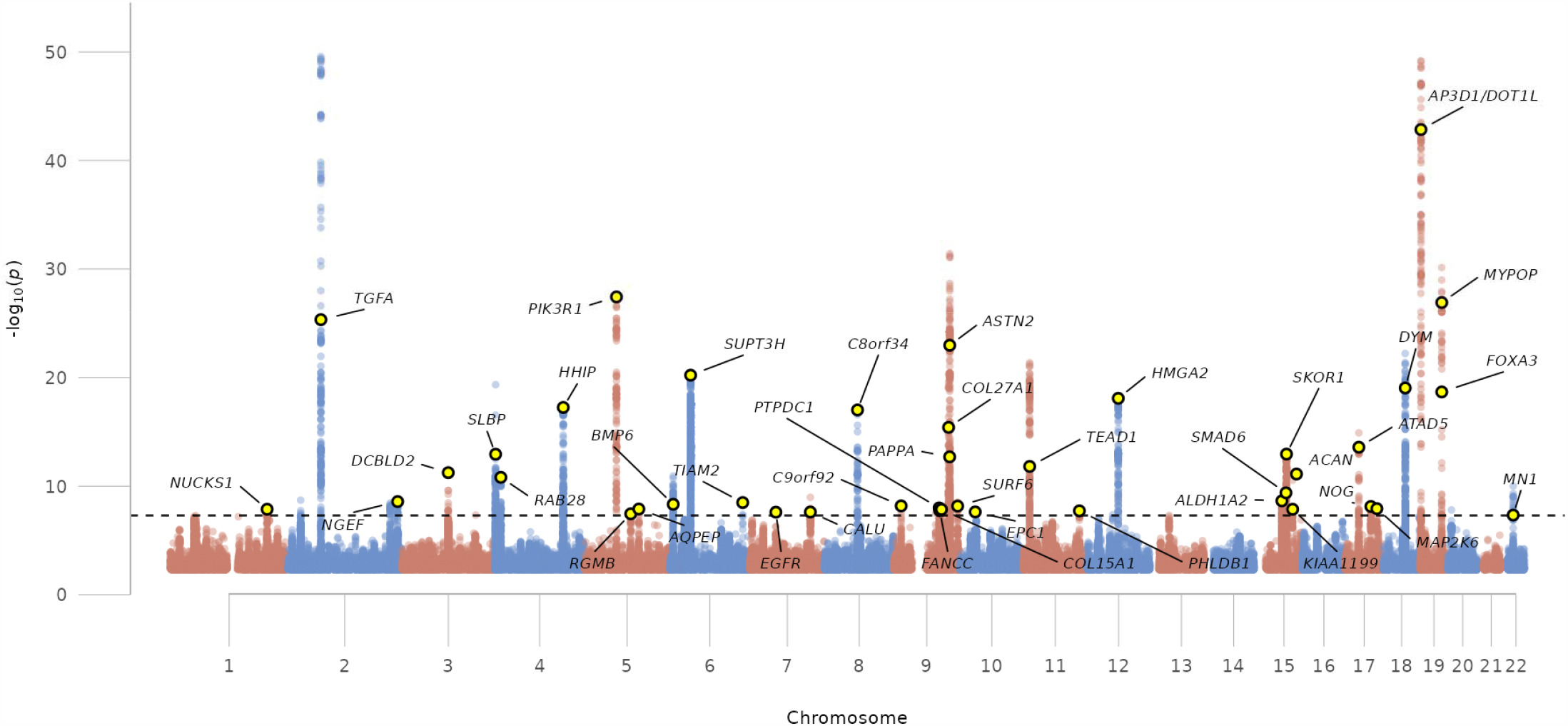
Manhattan plot showing mJSW genome-wide meta-analysis results. The dashed black line denotes the threshold for declaring genome-wide significance (5.0×10-8). Yellow circles represent novel mJSW loci (defined as > 1MB from previously reported genome-wide significant mJSW variants).

### Genetic correlation

LDSC provided estimates of genetic correlation. A strong genetic correlation was seen between mJSW_DXA_ (N=38,175) versus mJSW_X-ray_ (N=12,570) (r_g_ 0.87 [95% CI 0.59, 1.14]). While the SNP heritability z-score for mJSW_DXA_ was 10.8, the SNP heritability z-score for mJSW_X-ray_ was 3.3, which is below the threshold of 4 that is suggested for reliable LDSC estimates (17). There was a moderate correlation between mJSW_combined_ (mJSW_DXA_ and mJSW_X-ray_ combined) versus height (r_g_ 0.28 [0.22, 0.33]) and between mJSW_DXA_ versus height (r_g_ 0.34 [0.28, 0.39]). There was minimal evidence of a genetic correlation of mJSW_combined_ versus BMI (r_g_ 0.06 [0.01, 0.12]) and versus HOA (r_g_ 0.10 [-0.01, 0.21]). However, when examined separately, there was some evidence of correlation between mJSW_DXA_ and BMI (r_g_ 0.08 [0.03, 0.14]) and HOA (r_g_ 0.14 [0.04, 0.25]) (Supplementary Table 3).

### Mendelian Randomisation and MR-Cluster

To examine the causal relationship between mJSW and osteoarthritis, we performed a two sample MR. 41 of the 42 independent mJSW lead SNPs were used as genetic instruments (mean F-statistic = 61). Rs34687269 was not included in the MR analyses because its alleles are palindromic. Despite good instrument strength, two sample MR showed no causal effect of mJSW on HOA (IVW: β -0.01 [95% CI -0.19, 0.17], MR Egger: β -0.37 [-0.91, 0.12] and Weighted Median: β -0.02 [-0.13, 0.09]) (Supplementary Figure 4 A). Subsequent cluster analysis of the mJSW genetic instruments displayed three distinct clusters, with two sample MR used to quantify each cluster’s effects: (i) Cluster one SNPs (n=11) were associated with a higher mJSW and a decreased risk of HOA (IVW: β -0.60 [95% CI -0.72, -0.48]); (ii) Cluster two SNPs (n=10) were associated with both greater mJSW and an increased risk of HOA (IVW: β 0.88 [0.72, 1.04]); and (iii) Cluster three SNPs (n=20) had no clear association with HOA (IVW: β 0.03 [-0.05, 0.11]) (Table 2 and Figure 2). Heterogeneity of SNP effects between mJSW and HOA identified by cluster analysis illustrated why no net causal effect between these traits was detected. To further understand these SNP clusters, SNP associations with other traits were investigated.

**Table 2.** Two sample Mendelian randomisation results.

| Exposure | Outcome | SNPs | IVW |  | MR Egger |  | Weighted median |  | Simple Mode |  | Weighted mode |  |
| --- | --- | --- | --- | --- | --- | --- | --- | --- | --- | --- | --- | --- |
|  |  |  | Beta (95% CI) | P | Beta (95% CI) | P | Beta (95% CI) | P | Beta (95% CI) | P | Beta (95% CI) | P |
| mJSW | HOA | 41 | -0.01 (-0.19-0.17) | 0.87 | -0.37 (-0.91-0.17) | 0.18 | -0.02 (-0.13-0.09) | 0.72 | -0.03 (-0.23-0.18) | 0.80 | -0.07 (-0.29-0.14) | 0.49 |
| Cluster 1 | HOA | 11 | -0.60 (-0.71- -0.49) | $2.47 \times 10^{-25}$ | -0.52 (-0.88- -0.16) | 0.02 | -0.58 (-0.72- -0.44) | $1.61 \times 10^{-16}$ | -0.55 (-0.81- -0.29) | $1.94 \times 10^{-03}$ | -0.66 (-0.90- -0.41) | $3.64 \times 10^{-04}$ |
| Cluster 2 | HOA | 10 | 0.88 (0.71-1.04) | $2.98 \times 10^{-26}$ | 0.68 (0.07-1.29) | 0.06 | 0.86 (0.68-1.04) | $8.62 \times 10^{-21}$ | 0.88 (0.59-1.16) | $1.99 \times 10^{-04}$ | 0.84 (0.62-1.07) | $3.71 \times 10^{-05}$ |
| Cluster 3 | HOA | 20 | 0.03 (-0.05-0.11) | 0.47 | 0.01 (-0.23-0.26) | 0.91 | 0.01 (-0.10-0.12) | 0.86 | 0.00 (-0.22-0.21) | 0.98 | -0.01 (-0.20-0.18) | 0.90 |
| Cluster 1 | Height | 11 | 0.06 (-0.02-0.14) | 0.16 | 0.20 (-0.05-0.45) | 0.16 | 0.01 (-0.01-0.03) | 0.17 | 0.01 (-0.01-0.03) | 0.45 | 0.01 (0.00-0.03) | 0.16 |
| Cluster 2 | Height | 10 | 0.25 (0.14-0.36) | $1.36 \times 10^{-05}$ | -0.07 (-0.40-0.26) | 0.69 | 0.10 (0.07-0.13) | $2.75 \times 10^{-11}$ | 0.30 (0.11-0.50) | 0.01 | 0.11 (0.08-0.13) | $1.01 \times 10^{-05}$ |
| Cluster 3 | Height | 20 | 0.09 (-0.01-0.19) | 0.07 | 0.09 (-0.22-0.40) | 0.58 | 0.02 (-0.01-0.04) | 0.22 | 0.02 (-0.01-0.06) | 0.25 | 0.01 (-0.01-0.03) | 0.22 |

Abbreviations: IVW – inverse variance weighted, MR – Mendelian randomisation, mJSW – minimum joint space width, SNP – single nucleotide polymorphism, CI – confidence interval, P – p-value, HOA – hip osteoarthritis

**Figure 2.**
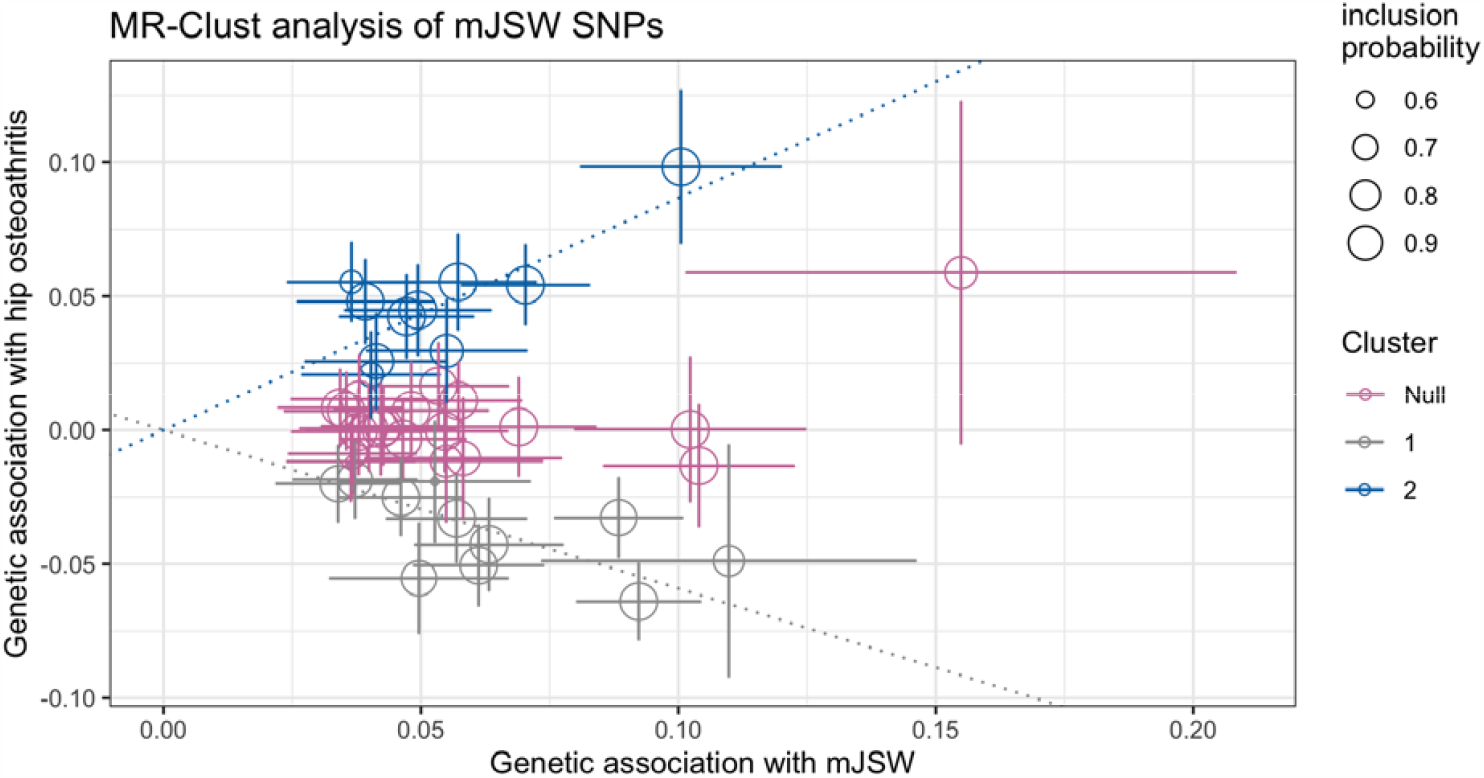
MR-Clust results Each independent minimum joint space width (mJSW) single nucleotide polymorphism (SNP) is plotted comparing their mJSW and hip osteoarthritis (HOA) effects. Three clusters are identified: Cluster one SNPs show a protective effect on HOA with increasing mJSW, Cluster two SNPs show an increasing risk of HOA with increasing mJSW and the null cluster SNPs show no effect on HOA.

### Trait look-ups and SNP prioritisation

The 42 independent mJSW-associated SNPs were examined in previous GWAS of HOA, height, and BMI (Table 1). SNPs in Cluster one (n=11), which were associated with a decreased risk of HOA with increasing mJSW, showed mixed associations with height; MR showed limited evidence of a small causal effect on height overall (IVW: β 0.06 [95% CI - 0.02, 0.14]) (Table 2). SNPs in Cluster two (n=10), which were associated with a higher HOA risk with increasing mJSW, all (except one for which a proxy SNP was not found) showed strong and consistent positive associations with height and some evidence of association with BMI, and MR showed a strong causal effect of these SNPs on height (IVW: β 0.25 [95% CI 0.14, 0.36]) (Table 2), Supplementary Figure 4E-F). Colocalisation analysis revealed that two SNPs in Cluster two (near *BMP6* and *MAP2K6*) shared common signals with height (PP=99% and 97%, respectively) (Supplementary Table 4).

The palindromic SNP (rs34687269) showed strong positive associations with mJSW, HOA and height, in keeping with rs76248879, a Cluster two SNP that was also situated close to the *ASTN2* locus (Table 1). There is a clear null cluster outlier (rs117564279) in Figure 2, which shows a strong mJSW effect (β 0.15, *P* 1.35×10^−8^) and weaker HOA (β 0.06, *P* 0.07) effect. Interestingly, it has no association with height (β 0.002, *P* 0.67). Rs117564279 is a rare allele with a MAF 0.02 (Table 1).

### Identification of candidate osteoarthritis pathogenesis genes

SNPs in Cluster one, which were thought to increase HOA risk through reduced mJSW and hence are potential targets for chondro-protective therapies, were assessed further. Colocalisation was used to compare GWAS signals between mJSW and HOA. Loci closest to *TGFA, COL27A1, C8orf34* and *SLBP* showed strong evidence of a shared signal (PP 1.00, 1.00, 0.99 & 0.97, respectively). No other loci showed such evidence (Table 3).

**Table 3.** Cluster one candidate gene identification

| C.Gene | RSID | HOA Beta | HOA P | HOA Coloc (PP) | MAGMA - Top Genes (p-value) |
| --- | --- | --- | --- | --- | --- |
| *TGFA* | rs7571789 | -0.06 | $5.32 \times 10^{-18}$ | 1.00 | *TGFA* ($4.83 \times 10^{-17}$) |
| *SUPT3H* | rs10948155 | -0.05 | $9.17 \times 10^{-11}$ | 0.03 | *SUPT3H* ($6.93 \times 10^{-15}$), *RUNX2* ($1.55 \times 10^{-12}$) |
| *EPC1* | rs76164690 | -0.06 | $1.59 \times 10^{-07}$ | 0.00 | *EPC1* ($3.41 \times 10^{-06}$) |
| *C8orf34* | rs7846438 | -0.04 | $1.48 \times 10^{-06}$ | 0.99 | *C8orf34* ($3.83 \times 10^{-09}$) |
| *AP3D1* | rs34656141 | -0.03 | $2.34 \times 10^{-05}$ | 0.40 | *DOT1L* ($3.11 \times 10^{-15}$) |
| *COL27A1* | rs4979342 | -0.03 | $9.40 \times 10^{-05}$ | 1.00 | *COL27A1* ($2.21 \times 10^{-12}$) |
| *SLBP* | rs2236996 | -0.03 | $6.74 \times 10^{-04}$ | 0.97 | *TMEM129* ($3.45 \times 10^{-12}$), *TACC3* ($9.78 \times 10^{-11}$), *SLBP* ($1.05 \times 10^{-14}$) |
| *MN1* | rs2106973 | -0.02 | $6.95 \times 10^{-03}$ | 0.16 | N/A |
| *ALDH1A2* | rs11857461 | -0.02 | 0.01 | 0.25 | *ALDH1A2* ($8.45 \times 10^{-09}$) |
| *TIAM2* | rs35199713 | -0.05 | 0.03 | 0.00 | N/A |
| *EGFR* | rs17172430 | -0.02 | 0.10 | 0.05 | N/A |

Cluster one SNPs were labelled with the closest gene. They were looked up in a HOA GWAS. Colocalisation was used to assess whether mJSW and HOA GWAS signals share common genetic causal variant in a given region with the posterior probability (PP) reported (H4: both traits are associated and share a single causal variant). Gene set analysis (MAGMA) was used to identify further candidate genes. The p-value threshold was $2.65 \times 10^{-06}$ . eQTL signals were assessed in GTEx using LocusFocus to conduct colocalisation with posterior probabilities reported. Cultured fibroblasts or if not present the tissue with the largest evidence of expression are reported. Abbreviations: C.Gene – closest gene, HOA – hip osteoarthritis, PP – posterior probability, Coloc – colocalisation, GTEx – genotyping expression project, N/A – not applicable, MAGMA – generalised gene-set analysis of GWAS data.

Subsequently, attempts were made to identify the underlying causal gene responsible for the SNP association. MAGMA assigned *TGFA, SUPT3H-RUNX2, C8orf34, EPC1, COL27A1, SLBP*-*TMEM129-TACC3, ALDH1A2* and *DOT1L* as candidate genes (Table 3). *TGFA* and *SUPT3H* mJSW association signals colocalised with GTEx expression in amygdala and basal ganglia respectively, but not in fibroblasts (Supplementary Table 5). The outlier SNP (rs117564279) with the largest effect size near *CEMIP* was also examined in GTEx and colocalised with eQTL SNPs in skeletal muscle (PP 0.98). RegulomeDB suggested the SNPs nearest to *TGFA, AP3D1, EGFR* and *TIAM2* were non-coding regulatory regions with probability scores >0.5 (Supplementary Table 6). Colocalisation between mJSW SNPs and human cartilage eQTL data provided no further gene-SNP evidence for our prioritised SNPs (Supplementary Table 7). However, it did show evidence of colocalisation for two null cluster SNPs; rs62479589 with *OPN1SW* in both highly and less degraded cartilage (PP 0.97 & 0.90 respectively) and rs823097 with *RAB7L1* in highly degraded cartilage (PP 0.96) (Supplementary Table 7).

### Gene ontology biological process annotations

PANTHER and FUMA GeNE2FUNC analyses showed that three Cluster two SNPs (which mapped to *ACAN, NOG, BMP6*) overlapped with skeletal system morphogenesis and development (Supplementary Tables 8 and 9).

## Discussion

In the largest GWAS of hip mJSW to date, we identified 42 conditionally independent SNPs, mapping to 39 loci of which 35 were novel. MR analysis revealed little evidence for a causal effect of mJSW on HOA risk, however, cluster analysis identified three groups of SNPs with distinct effects. One cluster comprised 11 SNPs which increase mJSW leading to a decrease in HOA risk. In contrast, a second cluster comprised 10 SNPs which increased mJSW but led to an increase in HOA risk. The latter set of SNPs was also related to height, a known risk factor for HOA. A null cluster comprised 20 SNPs with no association with HOA risk. Taken together, these findings suggest that SNPs associated with mJSW may exert distinct effects on HOA risk according to whether this is instrumented by SNPs which are also related to height.

Of the 11 loci identified by MR-Clust, which were protective for HOA with increasing mJSW (Cluster one), *TGFA, C8orf34, COL27A1* and *SLBP-TMEM129-TACC3* colocalised with the same causal signal for HOA in a large-scale HOA GWAS. The present findings suggest that these previously identified loci cause HOA through reduced cartilage thickness, suggesting potential utility as therapeutic targets for chondro-protective therapy. *TGFA* was implicated in mJSW by a previous much smaller GWAS and is known to be involved in endochondral bone formation (11, 33-35). Likewise, *COL27A1* is established in cartilage regulation and formation, and mutations are associated with osteochondrodysplasias in humans such as Steel syndrome which feature early hip dislocations and OA (36, 37). There is little known about *C8orf34* regulation of joint tissues such as cartilage but it has been implicated in vertebral disc disease (32). In addition, MAGMA suggested *TMEM129, SLBP* and *TACC3* might be the genes responsible for the association with mJSW at rs2236996locus but this was not supported by eQTL findings. *TMEM129* mutations can lead to facial dysmorphias such as Wolf-Hirschhorn syndrome and has been suggested to be a genetic risk factor for OA through disrupted protein degradation in the endoplasmic reticulum (29-31).

The other loci identified in Cluster one, *SUPT3H-RUNX2, AP3D1, EPC1, MN1, ALDH1A2, TIAM2* and *EGFR* did not colocalise with corresponding HOA GWAS signals but nonetheless showed at least a nominal HOA association. The *SUPT3H-RUNX2* locus was identified in the previous mJSW GWAS and has been implicated in chondrocyte and osteoblast differentiation respectively (11, 35). MAGMA suggested *EPC1* as a candidate for rs76164690, however this signal did not colocalise with eQTL expression in fibroblasts. Pigment epithelium derived factor (PEDF) is the product of the *EPC1* gene and is known to be anti-angiogenic. Previously PEDF has been shown to be preferentially expressed in OA cartilage contributing to OA pathogenesis by upregulating matrix degrading factors (38, 39). Whilst there was evidence of an association between rs34656141 with eQTL expression for *AP3D1, DOT1L* and *AMH* in fibroblasts these signals did not colocalise. *DOT1L* was previously implicated in mJSW in a smaller GWAS and is known to regulate cartilage homeostasis and protect against OA (11, 40). Anti-Mullerian Hormone, the product of *AMH*, is associated with knee OA in women (41). *ALDH1A2, TIAM2* and *EGFR* showed less evidence of an association with HOA, that said, *ALDH1A2* and *EGFR* have previously been identified as potential treatment targets for OA (42, 43). Less is known about *AP3D1, TIAM2* and *MN1* in the context of cartilage and HOA.

One locus showed a SNP effect of increased mJSW and HOA risk that was not associated with height; rs117564279 (*CEMIP*) is a rare variant with a MAF 0.02 and large effect size for both mJSW and HOA (β 0.15 & β 0.06 respectively). *CEMIP* is the closest gene and showed colocalisation between eQTL expression (skeletal muscle) and the mJSW GWAS signal. *CEMIP* has recently been shown to be expressed in cartilage from osteoarthritic joints, and to induce a fibrosis type response within chondrocytes (44). Therefore, *CEMIP* warrants further investigation to understand if altered expression leads to thicker more fibrous cartilage which in turn could lead to a wider joint space and a higher risk of HOA.

The opposing effects of SNPs in clusters one and two, as shown by the MR analyses of each cluster, presumably lead to a net null effect of mJSW on HOA. This may help to explain why mJSW, when examined observationally, displays little or no associations with HOA and symptoms, yet a decreased mJSW is often seen clinically in severely symptomatic individuals (45). Our observation that Cluster two SNPs are related to both height and HOA is consistent with previous findings that height GWAS signals overlap with OA (46, 47). This also corresponds with findings from observational studies that taller individuals are at an increased risk of HOA (48, 49). Whereas Cluster two SNPs are related to height, mJSW GWAS results showed little attenuation following height adjustment. Therefore, Cluster two SNPs appear to increase HOA risk through co-association with greater height, although height itself does not appear to be on the causal pathway for mJSW, suggesting the role of an intermediary growth-related mechanism (Figure 3). Consistent with this suggestion, gene ontology annotation suggested that three Cluster two SNPs (*ACAN, NOG, BMP6*) have a role in skeletal development. Extra-skeletal endocrine actions that influence growth might also play a role, given two loci, *PAPPA* and *PIK3R1*, are involved in the action of IGF-1 and insulin (50, 51).

**Figure 3.**
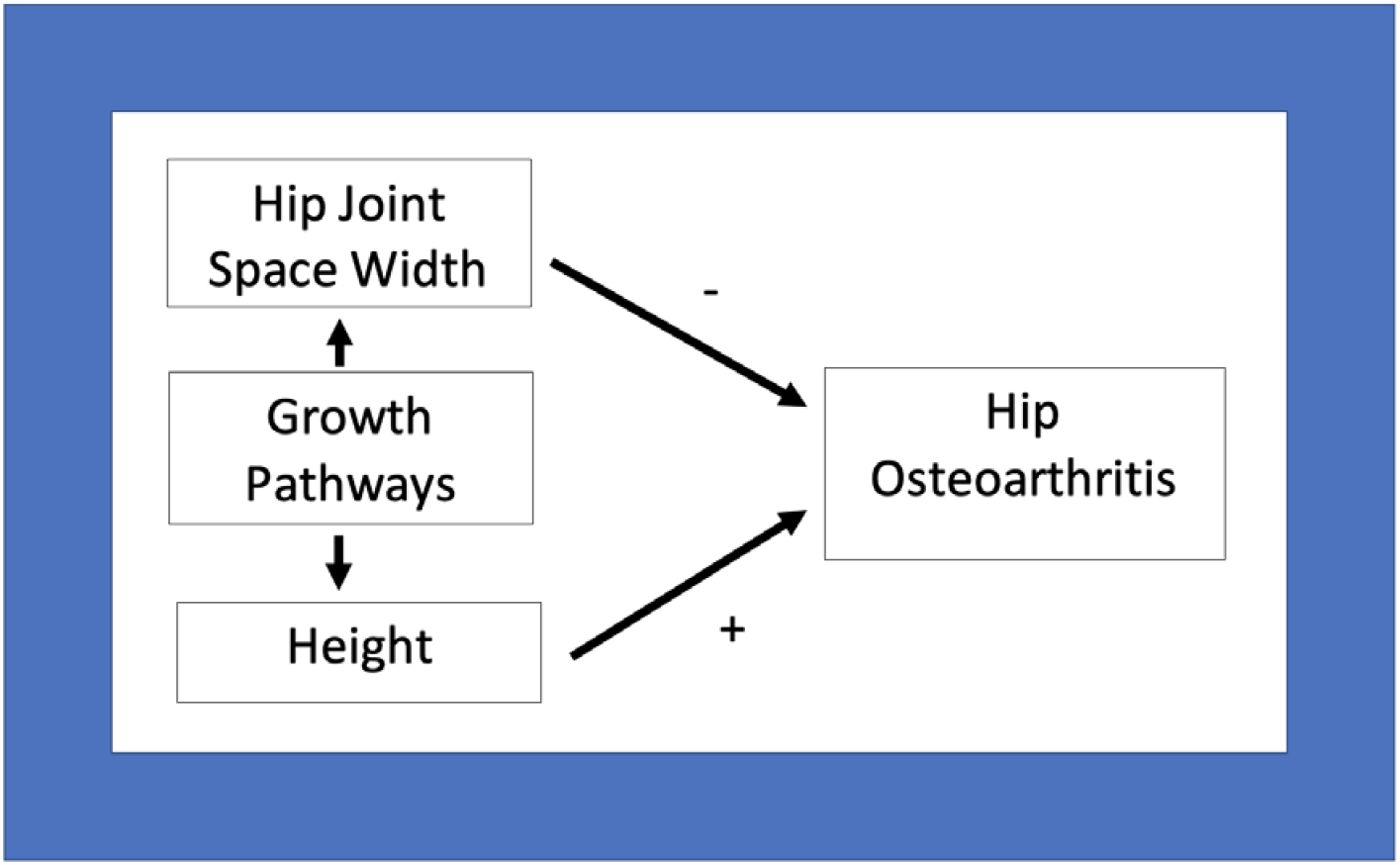
A directed acyclic graph to represent the proposed relationships between hip minimum joint space width, height and hip osteoarthritis.

The strengths of this study include its large sample size which has afforded the power to identify 35 novel loci. In addition, by combining a GWAS meta-analysis with other genetic analyses such as LDSC, MR and MR-Clust we have been able to tease out different causal pathways related to mJSW. In terms of limitations, as this is a GWAS of individuals with European ancestry this limits generalisability to other ancestries. In addition, we combined DXA and X-ray based cohorts, despite certain differences in mJSW measurements using these methods. For example, unlike DXA scans where only the superior joint space can be evaluated, mJSW can also be measured on X-rays at other sites. That said, X-ray based mJSW measurements were based solely on the superior joint space in SOF and MrOS. In contrast, in RS, mJSW measurements were also obtained laterally, axially and medially, with the smallest value used. That said, genetic correlation between mJSW obtained using these two methods was relatively high, albeit the mJSW_X-ray_ GWAS was underpowered for LDSC analysis. Furthermore, there was limited evidence of colocalisation between GWAS and eQTL data which hinders the identification of effector genes. However, it is increasingly recognised that many true GWAS signals fail to colocalise with eQTL signals (52, 53).

In conclusion, we present findings from a GWAS meta-analysis of hip mJSW which identified 35 novel loci and replicated 4 known loci. Subsequently, we showed that mJSW SNPs act on HOA in two distinct clusters; those that decrease HOA risk with increasing mJSW and those that increase HOA risk via increasing mJSW. We postulate the first group of SNPs may act via cartilage mediated pathways, suggesting possible utility as targets for chondroprotective therapies. In contrast, the latter group of SNPs are associated with greater height and likely act through growth-related mechanisms which require further clarification.

## Data Availability

The UK Biobank mJSW data from this study will be available in a forthcoming data release. Users must be registered with UK Biobank to access their resources (https://bbams.ndph.ox.ac.uk/ams/).

## Acknowledgements

This work has been conducted using the UK Biobank resource (application number 17295). The authors would like to thank the study participants, the staff from the Rotterdam Study and the participating general practitioners and pharmacists. The generation and management of GWAS genotype data for the Rotterdam Study (RSI & RSII) was executed by the Human Genotyping Facility of the Genetic Laboratory of the Department of Internal Medicine, Erasmus MC, Rotterdam, The Netherlands. The authors would like to thank Pascal Arp, Mila Jhamai, Marijn Verkerk, Lizbeth Herrera and Marjolein Peters, MSc, and Carolina Medina-Gomez, MSc, for their help in creating the GWAS database, and Linda Broer PhD, for the creation of the imputed data.

## Funding and grant award information

MF, RE, FS are supported, and this work is funded by a Wellcome Trust collaborative award (reference number 209233). BGF is supported by a Medical Research Council (MRC) clinical research training fellowship (MR/S021280/1). ML is supported by a University of Queensland Research Training Scholarship from The University of Queensland (UQ). ML thanks the Commonwealth Scientific and Industrial Research Organisation for the support through a Postgraduate Top-Up Scholarship. CL was funded by the MRC (MR/S00405X/1) as well as a Sir Henry Dale Fellowship jointly funded by the Wellcome Trust and the Royal Society (223267/Z/21/Z). This research was funded in whole, or in part, by the Wellcome Trust [Grant numbers 080280/Z/06/Z, 20378/Z/16/Z, 223267/Z/21/Z]. For the purpose of open access, the authors have applied a CC BY public copyright licence to any Author Accepted Manuscript version arising from this submission. NCH acknowledges support from the MRC (MC_PC_21003; MC_PC_21001) and NIHR Southampton Biomedical Research Centre, University of Southampton and University Hospital Southampton. BGF, MF, AEH, GDS, JHT work in the MRC Integrative Epidemiology Unit at the University of Bristol, which is supported by the MRC (MC_UU_00011/1). JPK is funded by a National Health and Medical Research Council (Australia) Investigator grant (GNT1177938). DSE acknowledges funding from NIH/NIA U24AG051129. The Osteoporotic Fractures in Men (MrOS) Study is supported by National Institute of Health funding from the following institutes: the National Institute on Aging (NIA), the National Institute of Arthritis and Musculoskeletal and Skin Diseases (NIAMS), the National Center for Advancing Translational Sciences (NCATS), and NIH Roadmap for Medical Research under the following grant numbers: R01 AR052000, K24 AR048841, U01 AG027810, U01 AG042124, U01 AG042139, U01 AG042140, U01 AG042143, U01 AG042145, U01 AG042168, U01 AR066160, and UL1 TR000128. The Study of Osteoporotic Fractures (SOF) is supported by National Institutes of Health funding. The National Institute on Aging (NIA) provides support under the following grant numbers: R01 AG005407, R01 AR35582, R01 AR35583, R01 AR35584, R01 AG005394, R01 AG027574, and R01 AG027576.

## Author contributions

Study design: MF, BGF; Data Analysis: MF, BF, RE, KAF, ML, AH, FRS.; Interpretation of results: MF, BGF, JPK, JT; Replication data: CGB, DSE.; Manuscript drafting: MF, BGF, JPK, JT.; Manuscript reviewing and editing: all authors.

