## Supplementary Methods for "Hip joint space width is causally related to hip osteoarthritis risk via distinct protective and susceptibility mechanisms: findings from a genome-wide association study meta-analysis"

The UK Biobank Study

UKB recruited 500,000 adults prospectively between 2006-2010 who have undergone comprehensive genetic and physical phenotyping. A comprehensive catalogue of variables is available here (http://biobank.ctsu.ox.ac.uk/crystal/) and extensive details of the cohort have been previously published here (59). The UKB extended imaging study has conducted hip DXA scans on ~50,000 individuals to date. Demographic information was obtained on the same day as the DXA scans. mJSW was measured automatically from outline points as has been described previously (3).

The Rotterdam Study

The Rotterdam Study (RS) is a prospective population-based cohort consisting of elderly inhabitants, 45 years and older, of the Ommoord district in the city of Rotterdam, the Netherlands. The design of the RS, data collection and genotyping has been described in detail in (1). Two phases of recruitment (I – 1989, those aged >55 years old & II – 2000, those aged >55 years old) are included in this study. The mJSW was assessed on pelvic radiographs taken in the anterior-posterior position, and have been described in (2,3). Briefly, the mJSW was measured in mm, along a radius from the centre of the femoral head, and defined as the shortest distance found from the femoral head to the acetabulum. Within the Rotterdam Study, we used a 0.5 mm graduated magnifying glass laid directly over the radiograph to measure the minimal joint space width of the hip joints (3).

Osteoporotic Fractures in Men Study (MrOS)

The Osteoporotic Fractures in Men Study (MrOS) is a prospective study of 5,994 men recruited between 2000-2002 at 6 centres around the United States (Birmingham, Alabama; Minneapolis, Minnesota; Palo Alto, California; the Monongahela Valley near Pittsburgh, Pennsylvania; Portland, Oregon; and San Diego, California). To be eligible, men had to be ≥ 65 years old, ambulatory, and without bilateral hip replacements. A full description of the MrOS cohort has been previously published (60, 61). Pelvic X-rays from which mJSW was obtained took place from March 2005 to May 2006.

Study of Osteoporotic Fractures (SOF)

The Study of Osteoporotic Fractures (SOF) is a multicentre cohort study started in 1986 (62). Participants were all female and aged ≥65 years at baseline and were recruited between September 1986 and October 1988 from population-based listings in 4 metropolitan areas of the US (62). Extensive information on this cohort has previously been published (62). Anteroposterior X-rays were obtained in 5,928 women from which mJSW was derived. Subjects with a diagnosis of rheumatoid arthritis, Paget's disease, or prior hip fracture or hip surgery were excluded from this analysis (62). The X-ray was done supine with the participants feet were placed at 15–30 degrees of internal rotation, the central ray was cantered on the symphysis pubis, and a 101-cm focal film distance at 70–80 kV (peak) was used. Using a caliper and reticule, mJSW from the acetabular roof to the femoral head according to previously published methods (20).

Minimum joint space width

In UKB, mJSW_DXA_ was measured automatically using a machine-learning trained algorithm that placed points around the femoral head and superior acetabulum on high resolution iDXA scans. Points were checked manually and corrected if necessary (~90% images required no point correction). Custom Python 3.0 script was subsequently applied to these points to derive mJSW (3, 14). Only superior joint space was examined in UKB. In the Rotterdam Study (RS) I&II, a 0.5 mm graduated magnifying glass was laid onto AP hip radiographs to measure mJSW_X-ray_ (15). In the Study of Osteoporotic Fractures (SOF) and Osteoporotic Fractures in Men Study (MrOS), a manual electronic caliper was used to measure the superior mJSW_X-ray_ to the nearest 0.1mm (16, 17).

GWAS in UK Biobank

*Minimum joint space width*

Genotyping, imputation and quality control (QC) were performed by UKB as previously described (63). Samples were genotyped using two genotyping arrays; Applied Biosystems UK BiLEVE Axiom Array by Affymetrix (49,950 participants) and Applied Biosystems UK Biobank Axiom Array (438,427 participants). Data were imputed using the HRC reference panel, and the merged UK10K and 1000 Genomes phase 3 reference panels in IMPUTE4.

A subset of European individuals from UKB (59) was used for GWAS. Ancestry assignment of UK-Biobank participants was performed as follows: The UKB sample was projected onto the first 20 principal components estimated from the 1000 Genomes Phase 3 (1000G) project (where ancestry was known) using GCTA version 1.93.2. Projections used a curated set of 38,512 LD-pruned HapMap 3 Release 3 (HM3) REF bi-allelic SNPs that were shared between the 1000G and UKB genotyped datasets (i.e. MAF > 1%, minor allele count > 5, genotyping call rate > 95%, Hardy-Weinberg P > 1x10^-6^, and regions of extensive LD removed). Uniform Manifold Approximation and Projection for Dimension Reduction (UMAP) was used in conjunction with the first 20 principal components to cluster 486,445 individuals using the following parameters: min_dist=0.0001, n_components=3, n_neighbors=45, random_state=10293082. UKB participants that clustered together with the 1000G European sub-populations were manually identified by visual inspection (N=461,920) and used for downstream genetic analyses.

We tested associations between genetic variants and sex stratified standardised (mean=1, SD=1) mJSW residuals assuming an additive allelic effect using linear mixed model (LMM) implemented in BOLT-LMM (v2.3) to account for cryptic population structure and relatedness, correcting each trait for age, sex, genotyping array and ancestry informative principal components 1 - 20 as previously described. GWAS involved high quality genome-wide imputed v3 genetic data (i.e. ~12 million SNPs, INFO > 0.3, MAF > 0.01%) measured in 38,175 related Europeans (for mJSW) from UKB.

*Hip Osteoarthritis*

An additional GWAS for HOA was completed to reduce sample overlap for the genetic correlation, two-sample MR, MR-Clust and colocalisation analyses. We conducted a GWAS of hospital diagnosed HOA (HES OA) n=323,948 in UKB, removing all individuals with a mJSW measure. HOA controls were selected as previously described (8). For example, individuals with inflammatory arthritis or OA at other sites were excluded from being controls. BOLT-LMMv2.34 was used for a GWAS of HES OA adjusting for age, sex, genotyping array and the first 20 ancestry principal components (cases=21,467, controls=302,481). Then the HOA GWAS meta-analysis was conducted using inverse variance weighted meta-analysis performed in METAL that combined the results of our HES OA GWAS in UKB with those from the Genetics of OA-consortium (GO) HOA GWAS meta-analysis, having excluded UKB (cases=46,704 & controls=574,765) (25). There is a small amount of sample overlap (<3%) between mJSW and HOA GWAS due to the presence of RS in both.

GWAS in Rotterdam

Genotypes from RS participates were imputed to the Haplotype Reference Consortium reference panel (V.1.0) using the Michigan Imputation Server.(4) We assessed genetic associations in each RS sub-cohort (RSI and RSII) using linear regression models of the standardised mJSW residuals (adjusted for age and sex) and additionally adjusted for the first four genetic principal components. RVtests(5) was used for the GWAS analyses and results were quality controlled using EasyQC.(6) Variants with an imputation quality <0.3, minor allele frequency <0.05 or effective allele count <5 were excluded and genomic control correction was applied to all SE and p values.

GWAS in SOF and MrOS

DNA extraction SOF:

In collaboration with Roche Molecular Systems (Alameda, CA), DNA from participants of the Study of Osteoporotic Fractures (SOF) was extracted from either buffy coat or whole blood samples collected at either visit 2 (1989–1990) or visit 6 (1997–1998). Among the 9704 SOF participants enrolled at the baseline visit, 6795 participants provided blood samples and consented to genetic testing. Among these 6795 SOF participants, DNA samples from 4117 participants had sufficient DNA quantity and were submitted to the Broad Institute for whole-genome genotyping. Among the 4117 SOF DNA samples, 3924 had sufficient DNA quantity and DNA solution volume and underwent whole-genome genotyping.

DNA extraction MrOS:

Genomic DNA from participants in the Osteoporotic Fractures in Men (MrOS) Study was extracted from whole blood samples collected at the baseline visit using the Flexigene protocol (Qiagen, Valencia, CA, USA) at the University of Pittsburgh. Among the 5994 MrOS participants enrolled at the baseline visit, 5530 participants provided blood samples, had DNA extracted, and consented to genetic testing. DNA samples from these 5530 participants were submitted to the Broad Institute for whole-genome genotyping. At the Broad Institute, DNA concentration was assayed by Picogreen and DNA solution volume was measured. Among the 5530 MrOS DNA samples, 5485 had sufficient DNA quantity and DNA solution volume and underwent whole-genome genotyping. All DNA samples eligible for whole-genome genotyping were genotyped using Sequenom iPLEX technology for a 24-SNP “fingerprint” panel.

Genotyping:

The Illumina HumanOmni1_Quad_v1-0 B genotyping array, containing 1,140,419 probes, was used for whole-genome genotyping. Samples from SOF and MrOS were randomized to 96-well genotyping plates by sex and clinic site. Eighty-one samples were plated twice to assess reproducibility. Pairwise concordance was 100%. 119 replicates of samples from HapMap trios of CEU and YRI populations and singletons from CHB and JPT populations were genotyped alongside MrOS and SOF samples, and compared to published HapMap genotypes. Concordance was 99.7% for CEU and YRI samples and was 95.0-99.7% for CHB and JPT samples.

Genotype quality control

Genotypes were called using a clustering algorithm in Illumina’s BeadStudio software at the Broad Institute. All genotype quality control was performed using custom scripts generated in the R statistical Language (version 2.12 and using the packages netCDF, v. 4.11 and GWASTools). Samples with call rates < 97% were excluded. SNPs with GenTrain scores <0.6, cluster separation scores <0.4, call rates <97.5%, or MAF <0.01 were excluded. Autosomal SNPs with HWE P-value <10^-4^ were excluded. In addition, genotype clusters for SNPs on chrX, chrY, chrXY and chrMT were reviewed manually. Heterozygous X-chromosome genotypes in MrOS were set to missing. For MrOS and SOF samples, 740,713 SNPs passed QC.

Additional samples were excluded based on: (1) genotypic sex mismatch using X and Y chromosome probe intensities, (2) relatedness among genotyped samples using the kinship coefficient that estimates probability that alleles are identical-by-descent, and (3) gross chromosomal abnormalities detected using the LogR Ratio and B allele frequency.

Among the 3924 SOF samples that underwent whole-genome genotyping, 3682 samples had acceptable call rates. Among these 3682 SOF samples, 4 were removed due to relatedness and 53 were removed due to gross chromosomal abnormalities, leaving 3625 SOF samples with whole genome genotyping data that passed QC. Among the 5485 MrOS samples that underwent whole-genome genotyping, 5168 samples had acceptable call rates. Among these 5168 MrOS samples, 1 was removed due to relatedness and 37 were removed due to gross chromosomal abnormalities, leaving 5130 MrOS samples with whole genome genotyping data that passed QC.

Population structure

Principal component analysis (PCA) was performed to detect evidence for population structure, exclude genetic ancestry outliers, and produce principal components to include in regression models to adjust for genetic ancestry (64). Unrelated samples from the CEU, YRI, CHB, and JPT HapMap populations were used to generate principal components. As there are three datasets in this study, Principal Components for the following were provided: MrOS all ethnicities, MrOS and SOF all ethnicities, MrOS white only, SOF white only, MrOS and MrOS Sweden white only, MrOS and SOF and MrOS Sweden white only, and MrOS Sweden white only.

The eigenvectors were calculated after removal of samples determined to be twins, sample with multiple cousins, and replicate samples as determined by IBD analysis. SNPs used for the calculation of the final eigenvectors had MAF>0.001 and linkage disequilibrium values < 0.05. For MrOS and SOF, eigenvectors were calculated using all ethnicities and again using those samples within 4SD of the mean after population analysis with HapMap samples

Genotype imputation

Before imputation, it was necessary to convert genotypes to plus-strand alleles. Minor alleles were then compared between HapMap samples and MrOS and SOF samples. SNPs and samples that passed QC filters underwent SNP genotype imputation using MaCH (v 1.0.17) for haplotype phasing and Minimac v 2011-08-12 beta for missing genotype imputation. 739,477 genotyped SNPs (714,543 autosomal SNPs) were available for imputation. HapMap phase II release 22 build 36 consensus phased haplotypes from a combined panel of CEU, YRI, CHB, and JPT HapMap samples were used as a reference panel. Genotype imputation was also performed using the 1000 genomes reference panel, genome build 37. For genome build-37, the X chromosome was imputed separately for males and females. For imputation done with the HapMap2 reference panel, 3,020,488 SNPs were imputed. For the 1000 Genomes reference panel, 40,314,357 SNPs were imputed.

Association analysis

Genetic association analysis was performed among individuals of European descent based on self-report and PCA-based genetic ancestry. Linear regression models adjusted for the effects of age, study site, and other covariates specified for the analysis. Potential population stratification was adjusted for by including the first four principal components in regression models. Imputed allele dosages were modeled with an additive mode of inheritance.

Minimum joint space width GWAS meta-analysis

Quality control of summary statistics was performed using EasyQC (20). Briefly, missing data, mono-allelic SNVs, implausible values (P > 1, infinite SE, beta >10, EAF>1.) and duplicates were removed from the data. We excluded variants with poor imputation quality (INFO <0.4) and minor allele frequency ≤0.005. Allele coding was harmonized across cohorts (A/T/C/G or I/D) and allele frequency checked against HRC imputed reference (http://www.haplotype-reference-consortium.org/) to identify possible allele coding errors. P-Z scatter plots were inspected for problems with beta estimates, standard errors and *P* values. Cleaned files were used to perform an inverse variance weighted fixed effects meta-analysis was performed with METAL (21). Following the meta-analysis SNPs were only considered if they were in more than one cohort and a SNP heterogeneity below a prespecified threshold (I^2^ ≤30). A separate GWAS meta-analysis was conducted excluding UKB so that mJSW_DXA_ and mJSW_X-ray_ could be compared.
