## Supplementary Figures for "Hip joint space width is causally related to hip osteoarthritis risk via distinct protective and susceptibility mechanisms: findings from a genome-wide association study meta-analysis"

### Supplementary Figure 1. Locus zoom plots for 42 leading mJSW SNPs

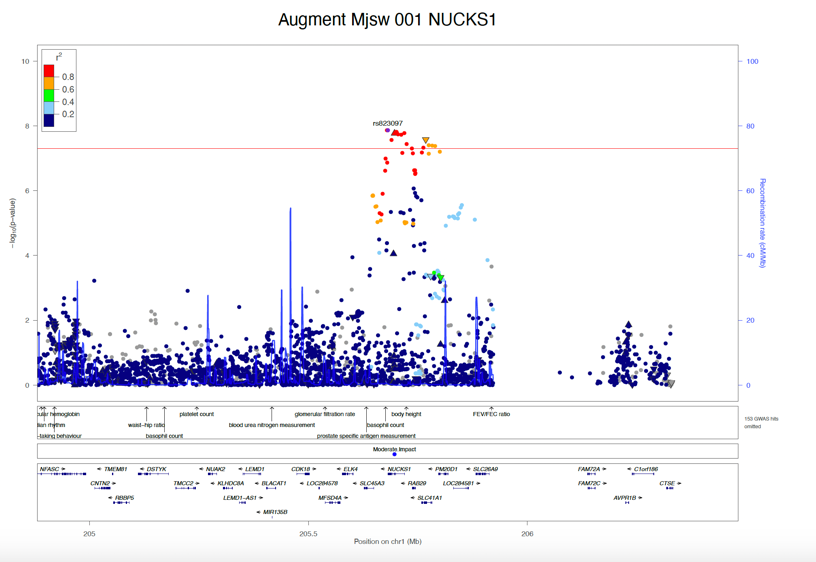

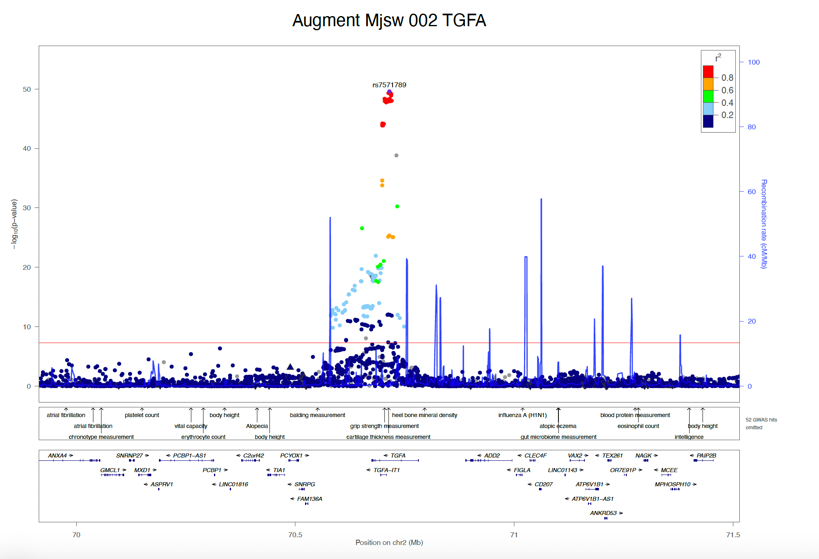

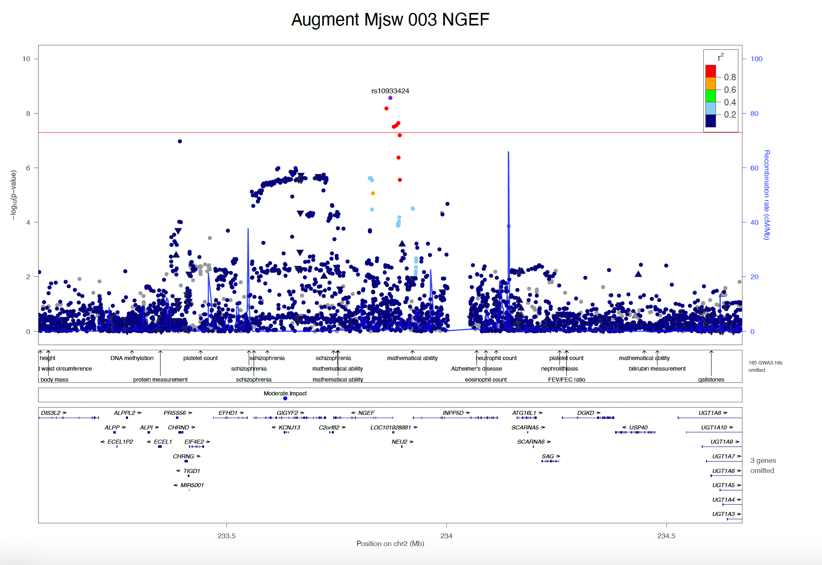

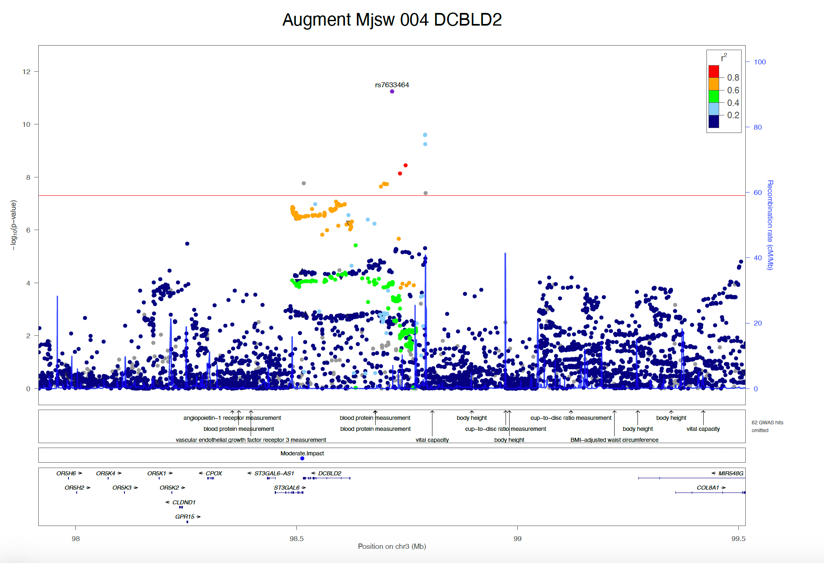

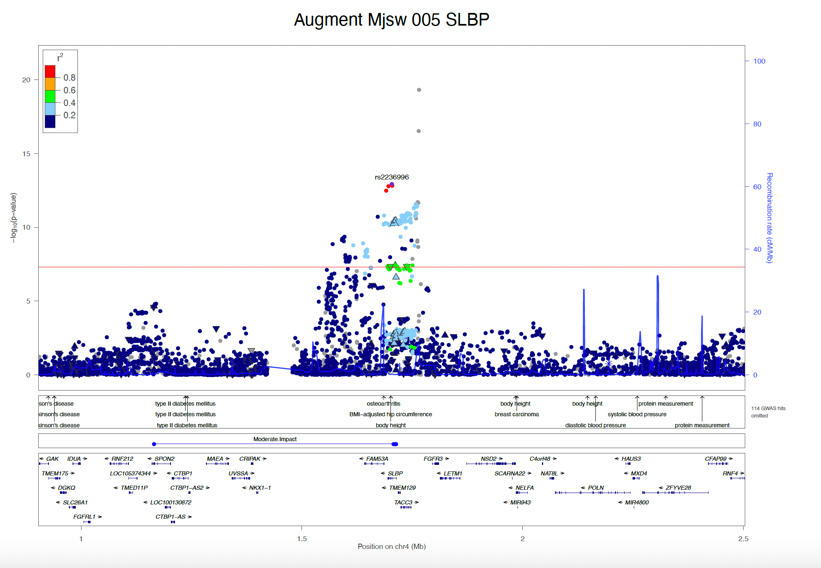

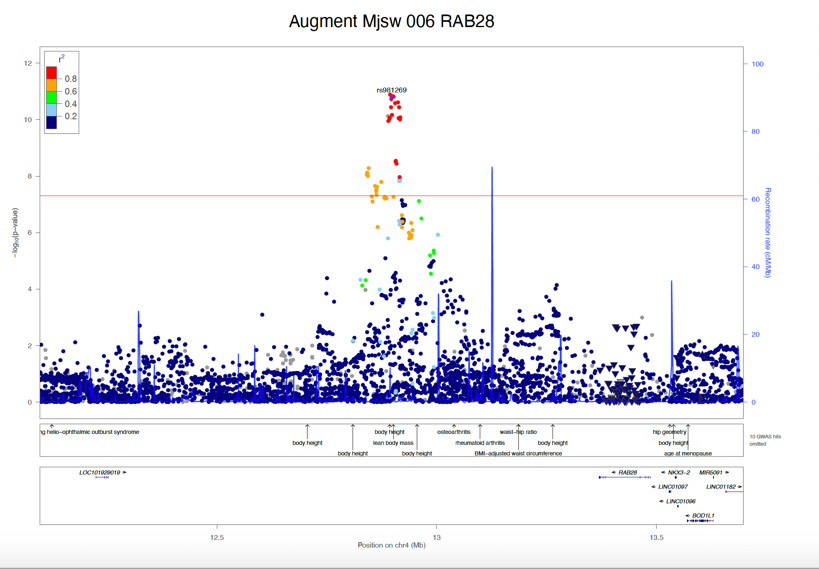

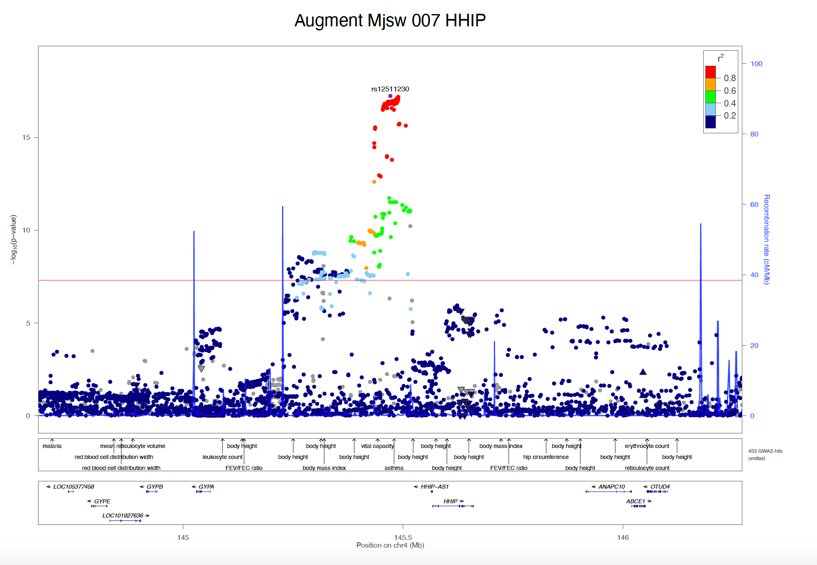

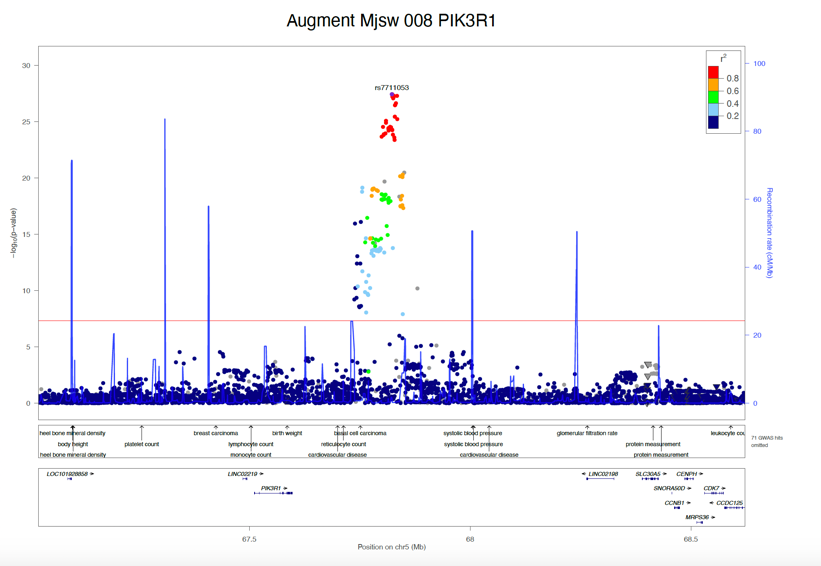

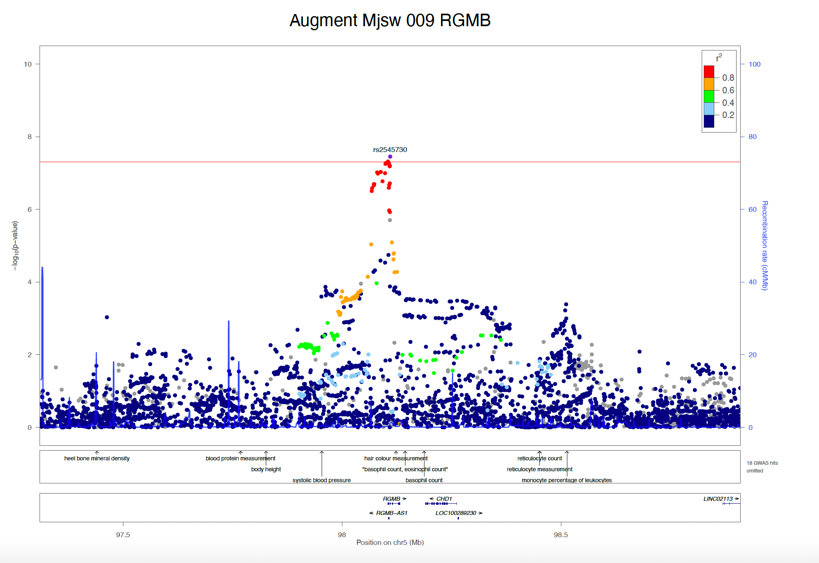

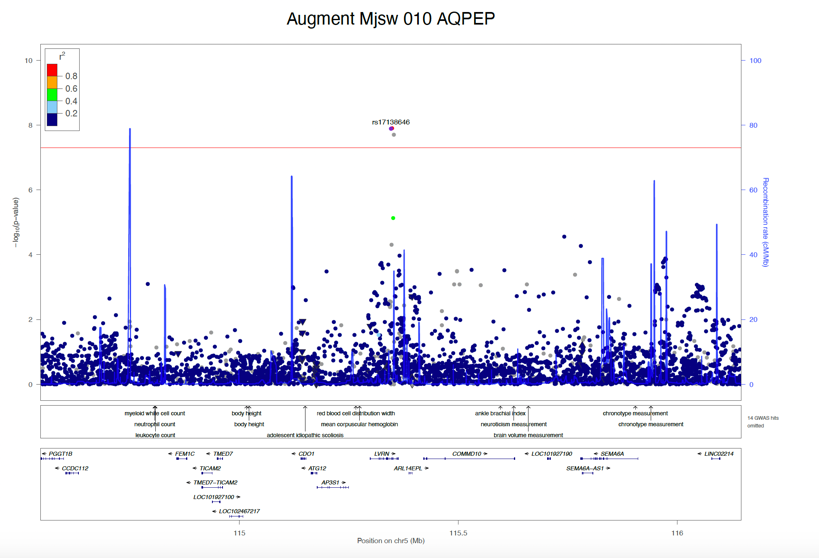

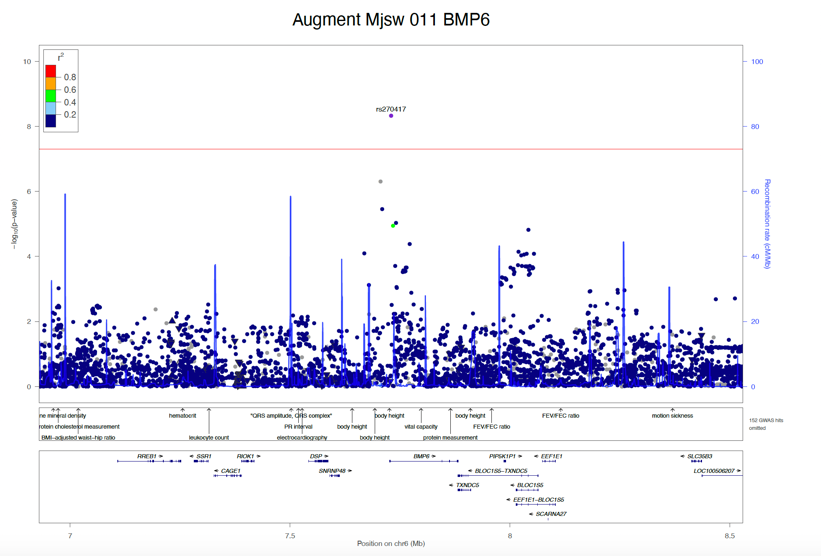

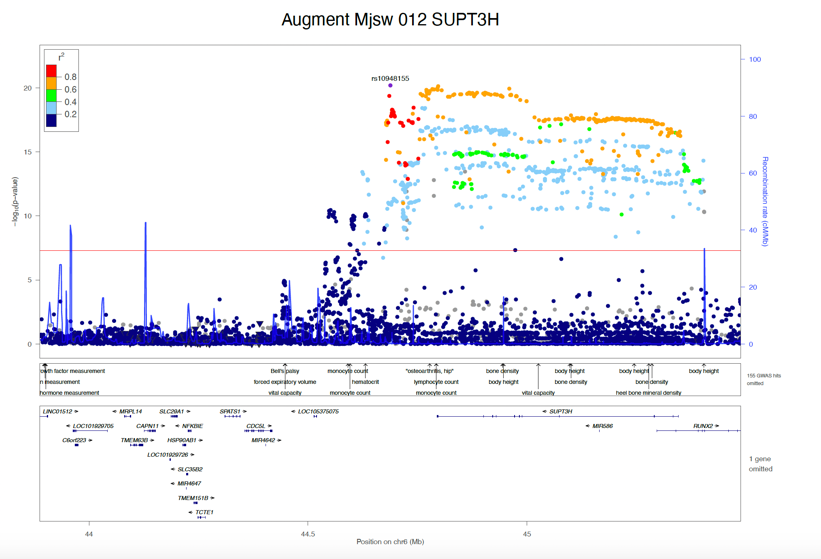

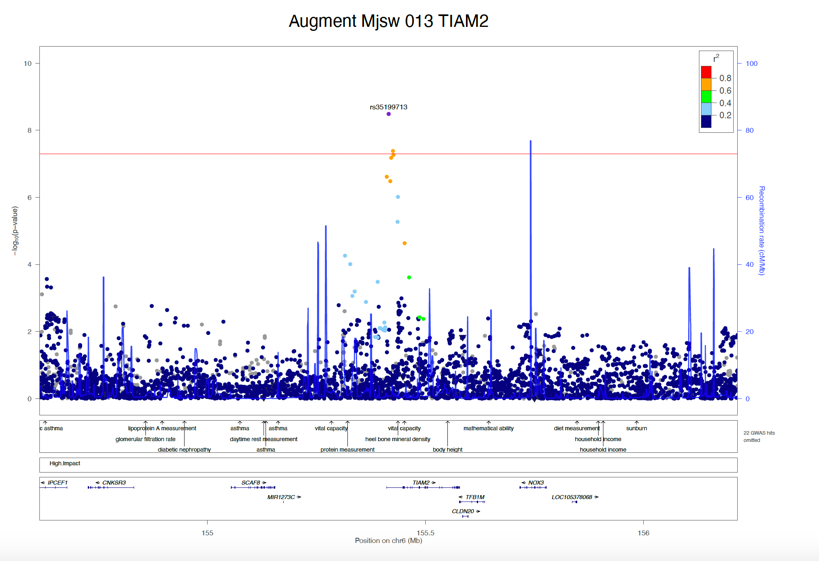

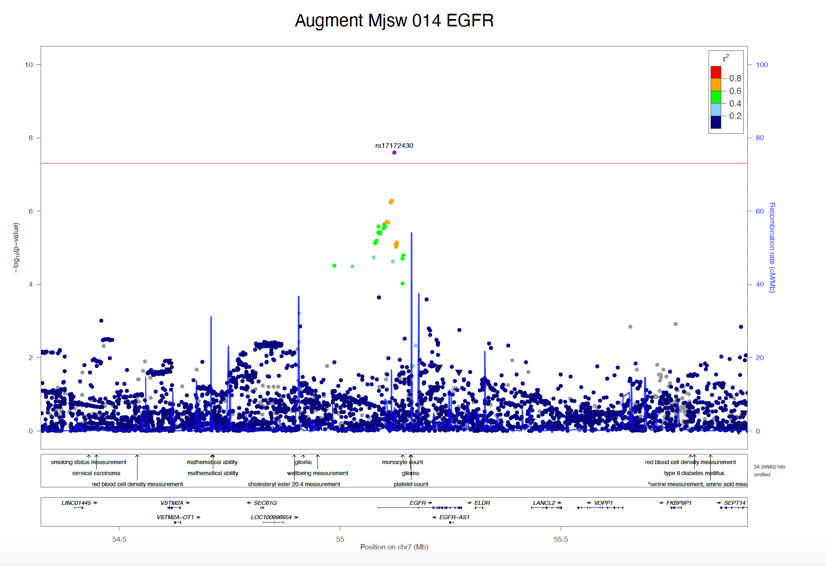

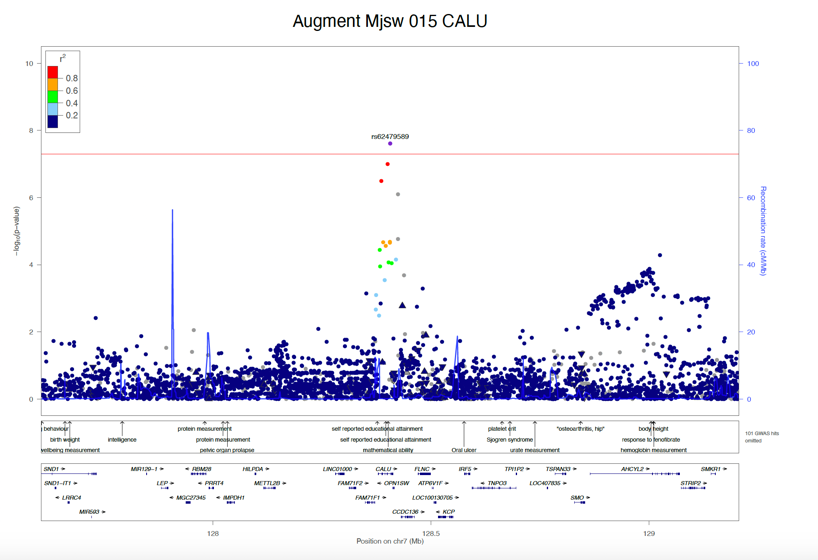

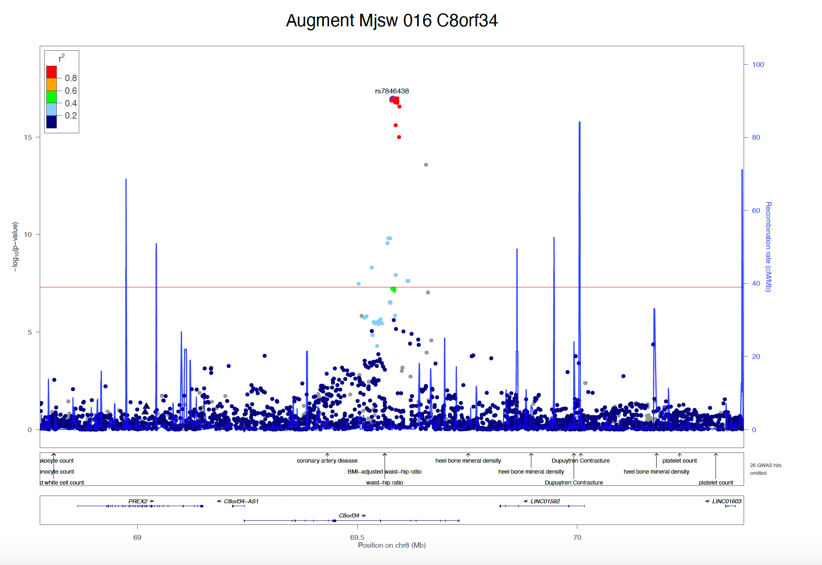

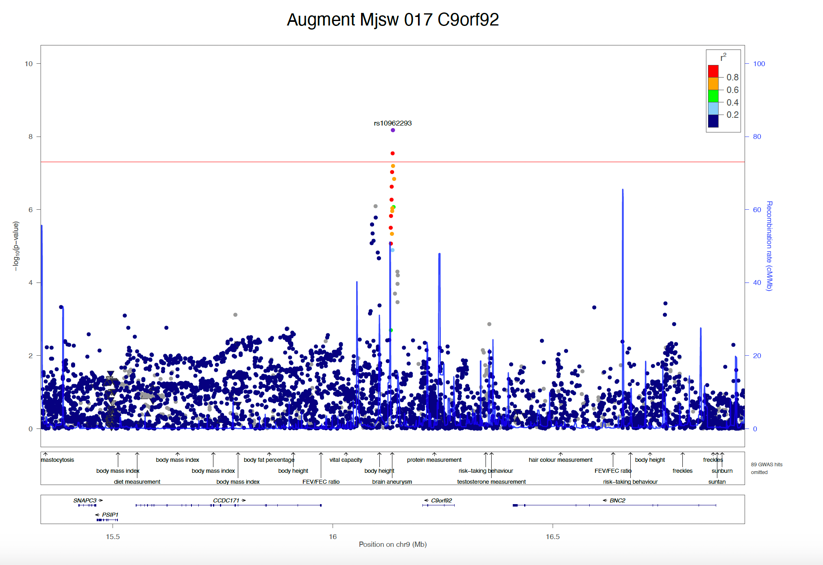

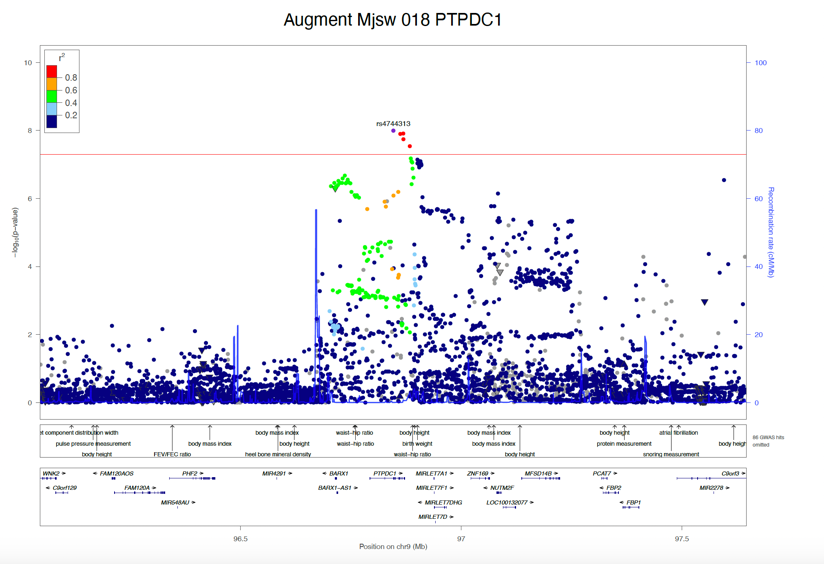

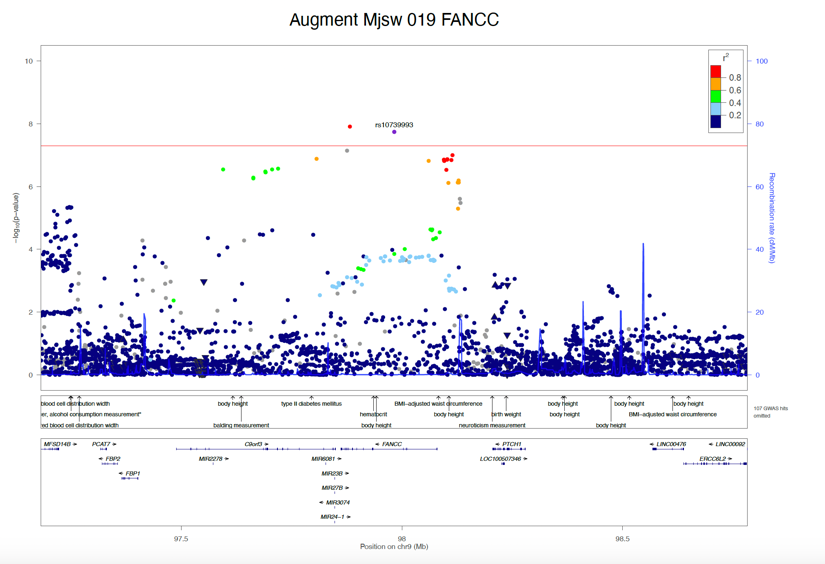

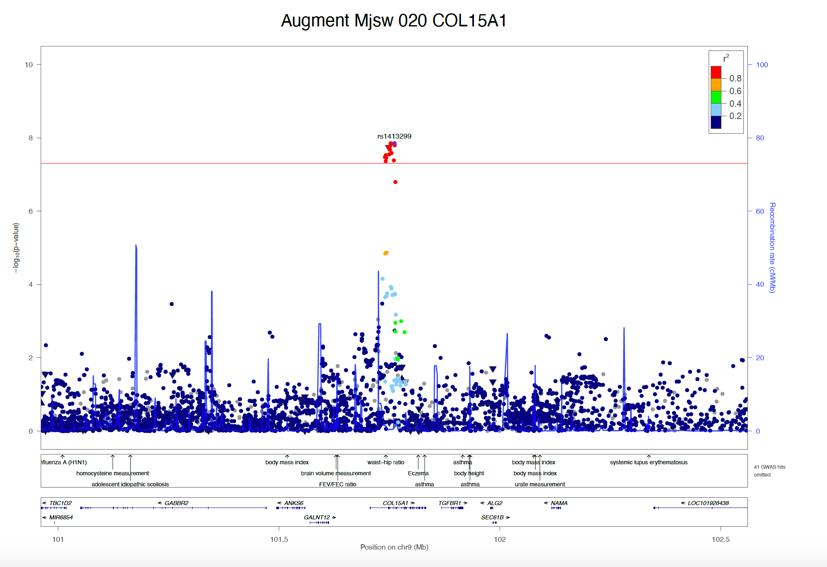

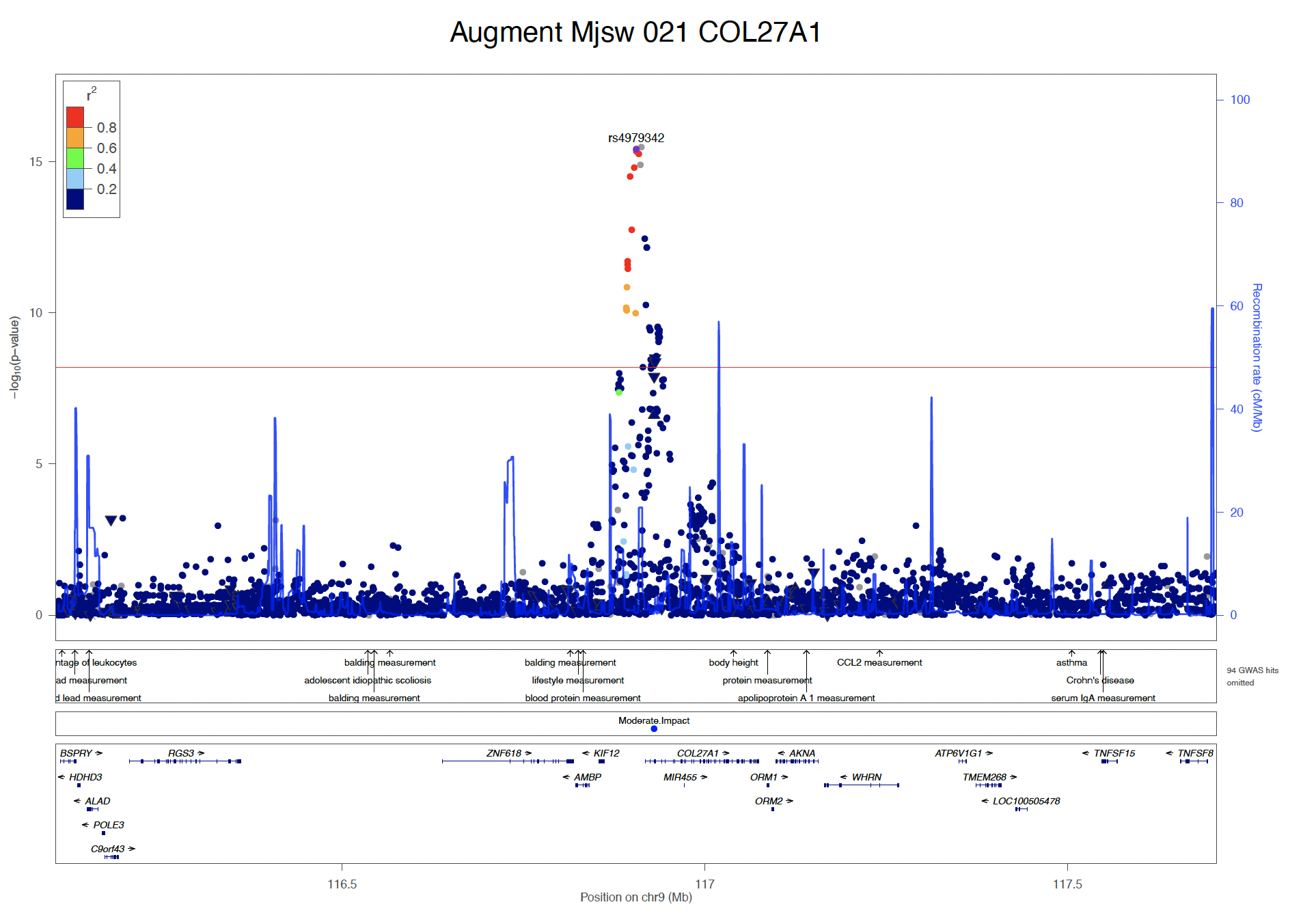

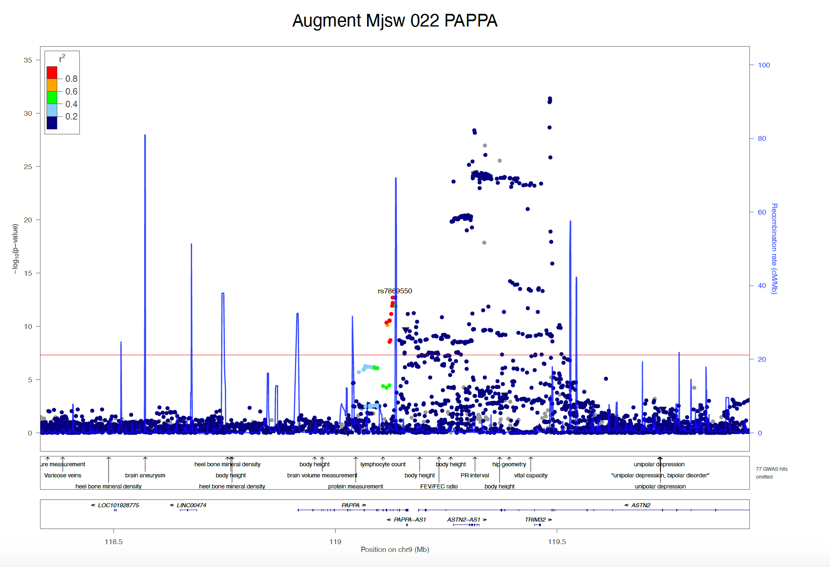

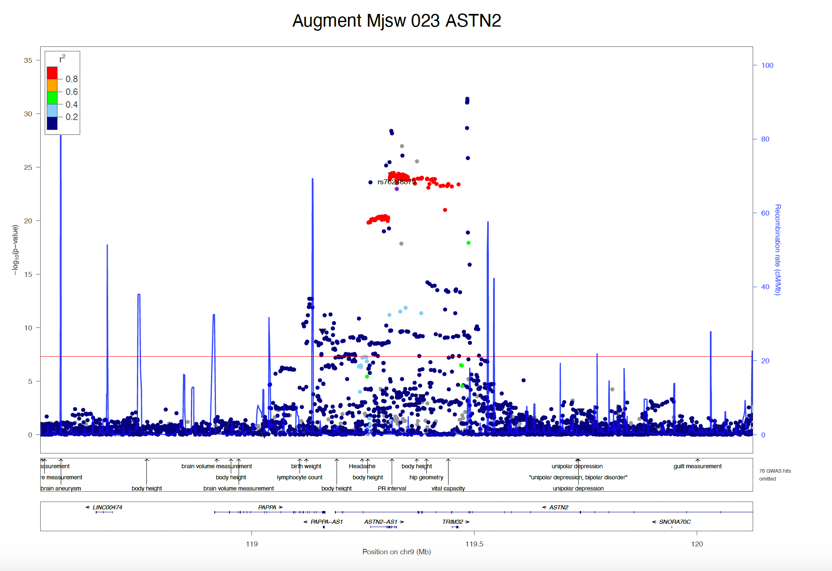

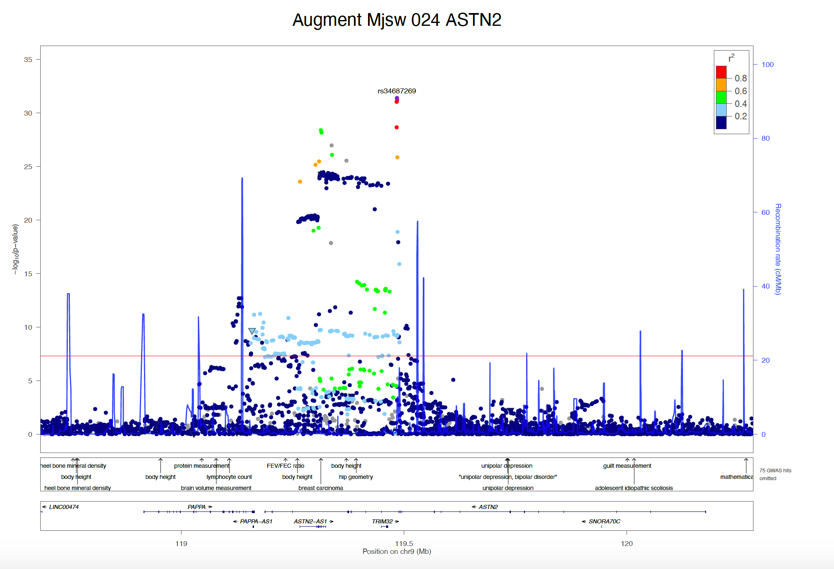

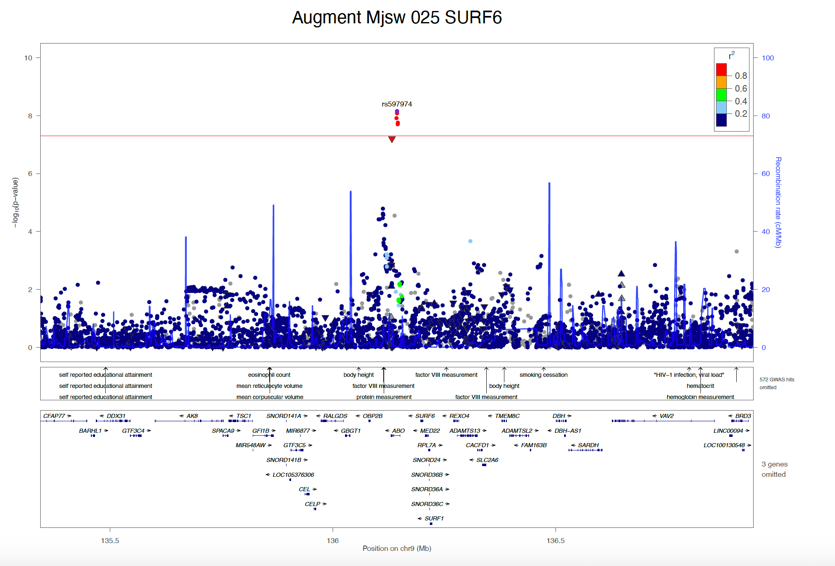

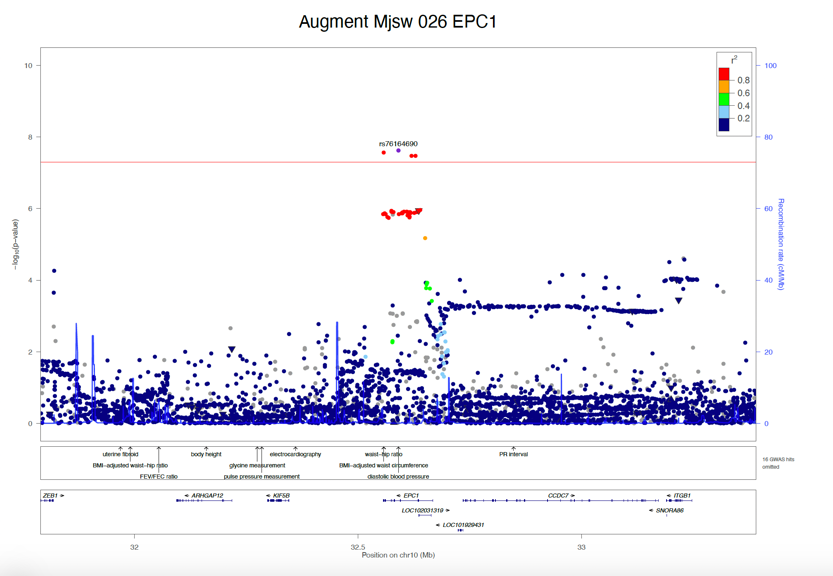

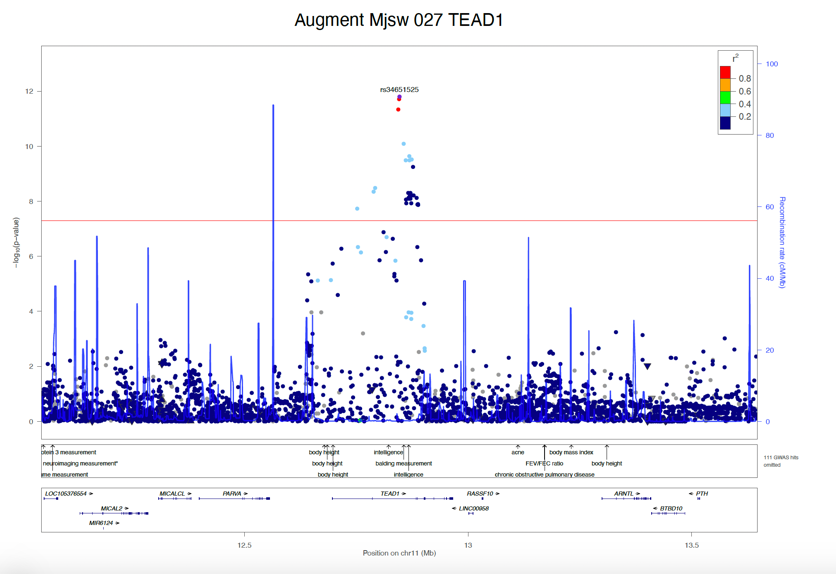

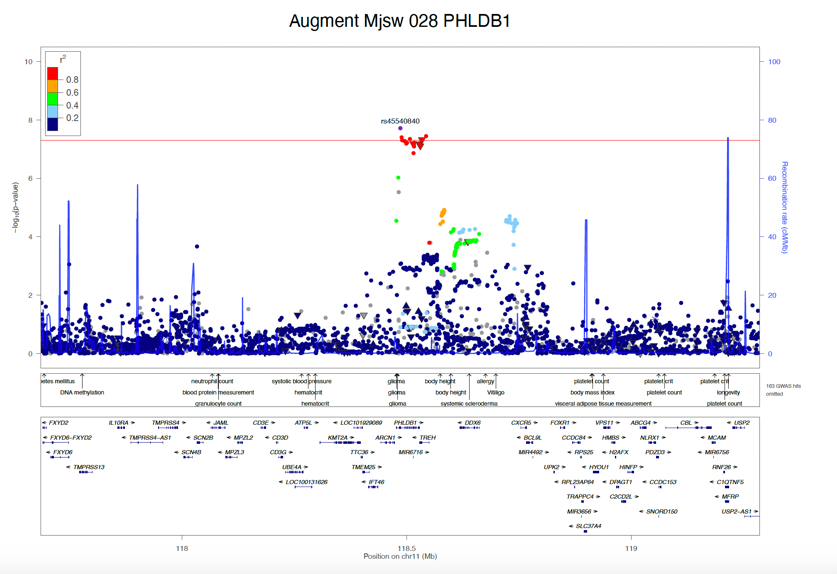

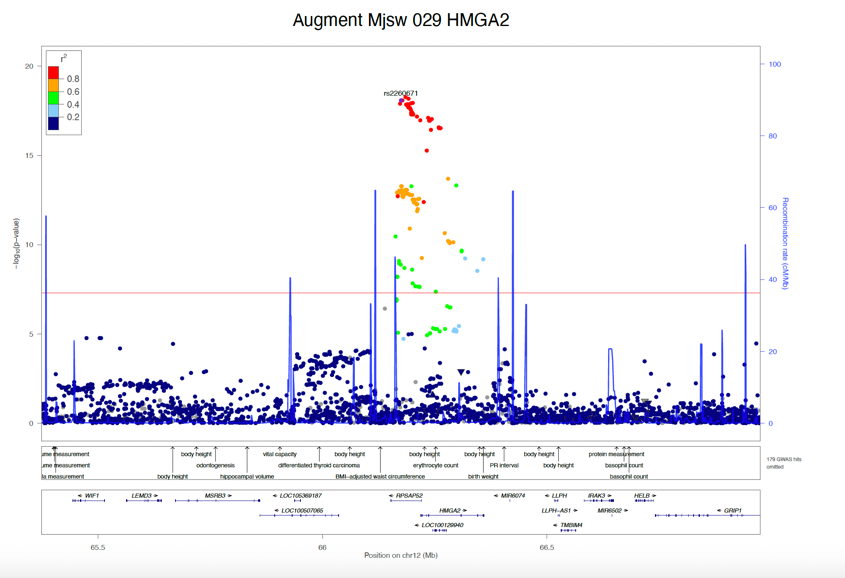

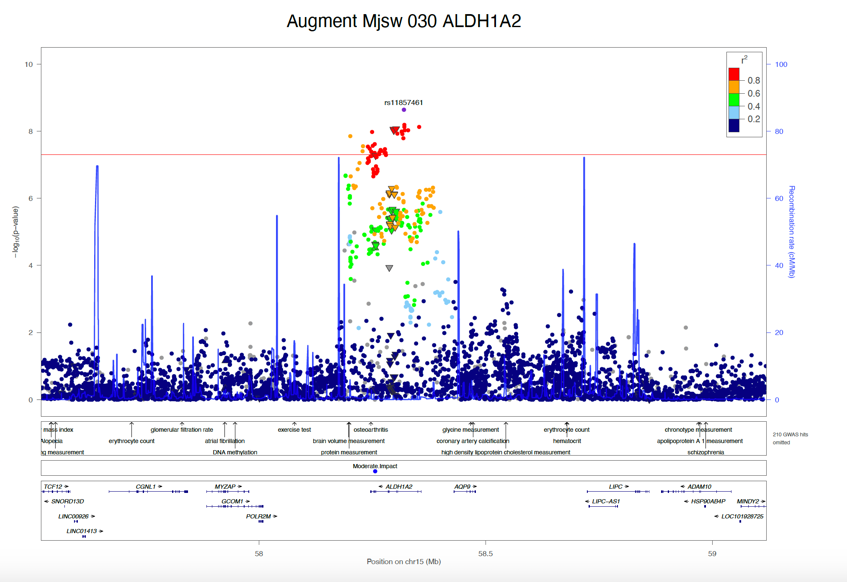

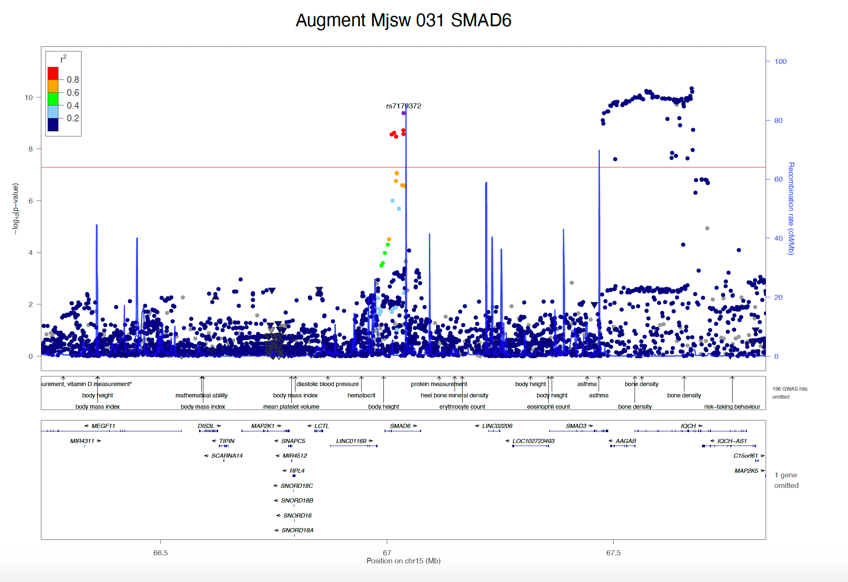

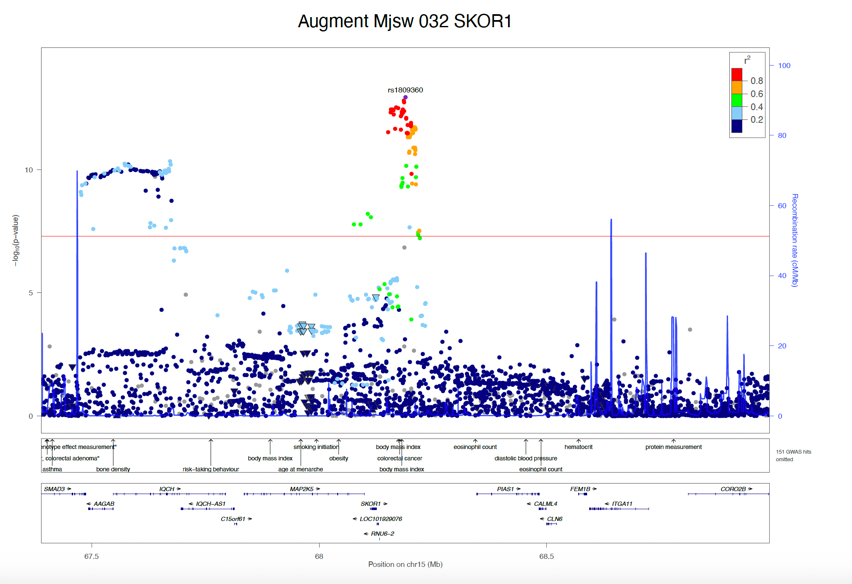

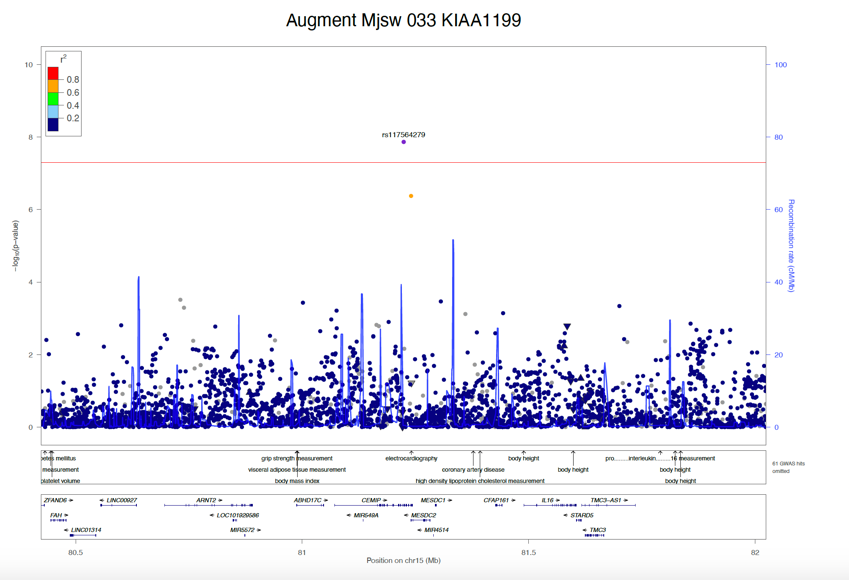

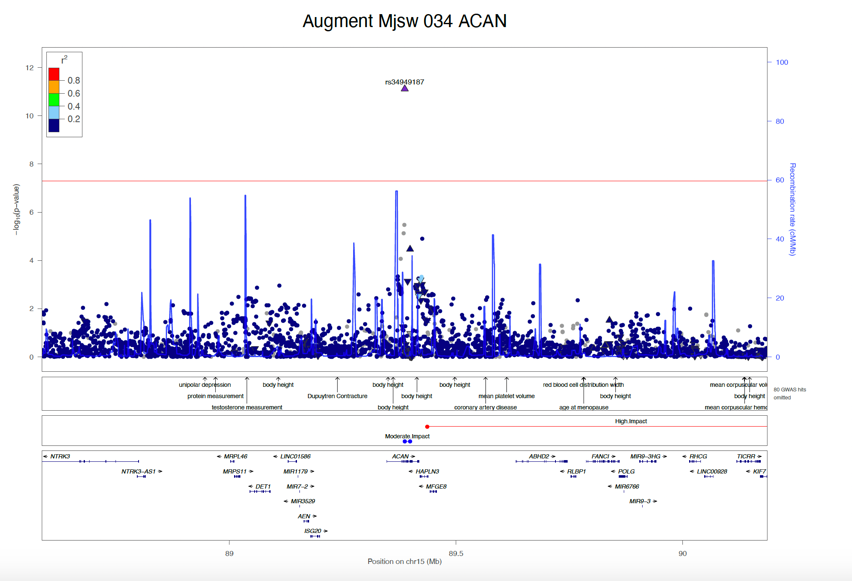

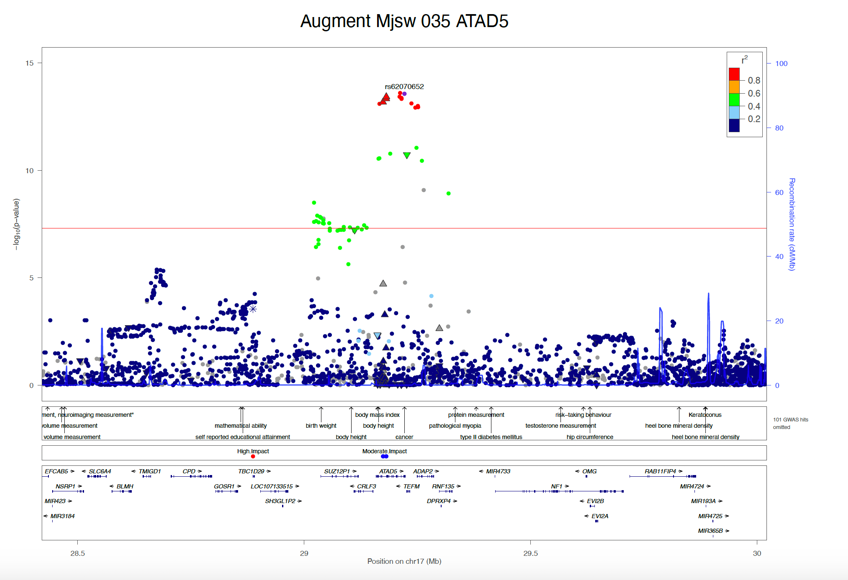

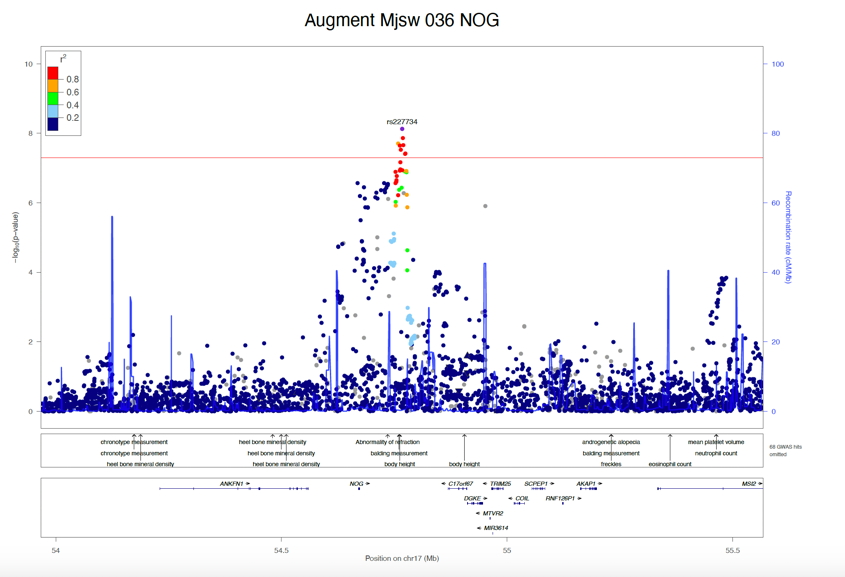

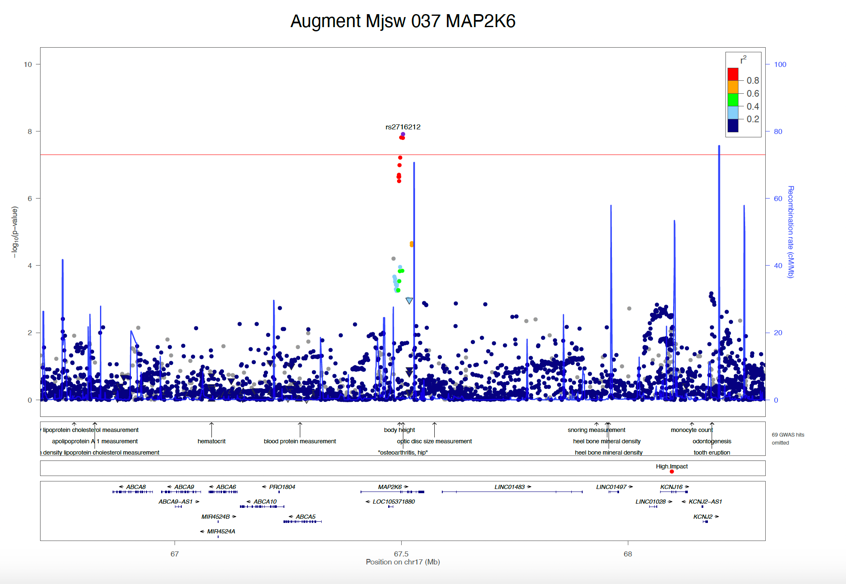

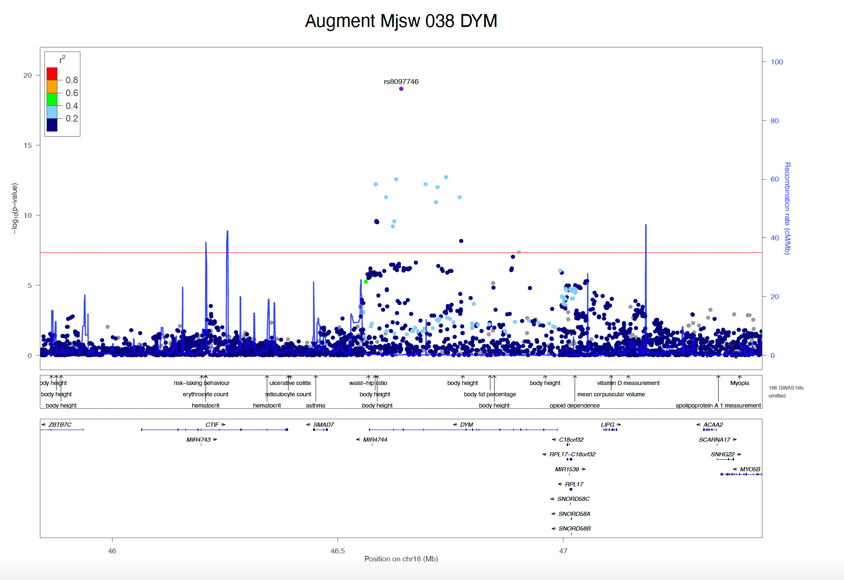

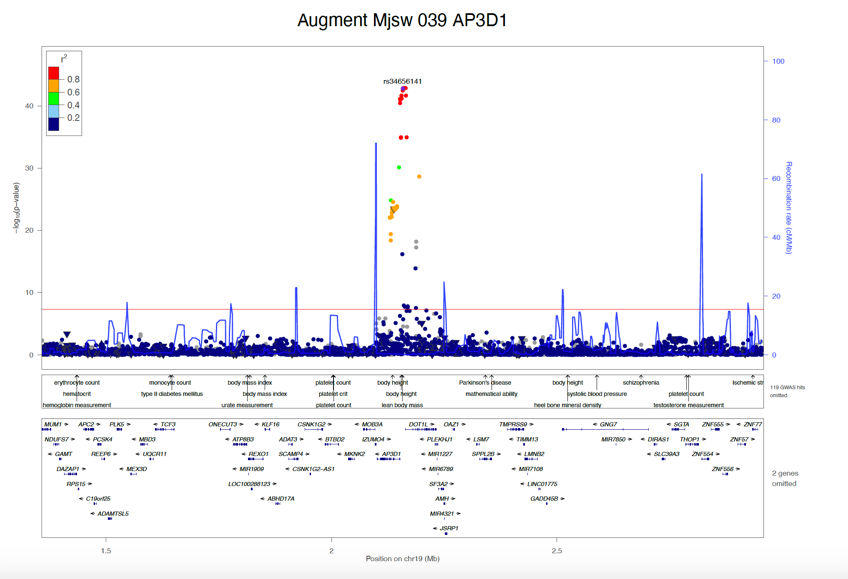

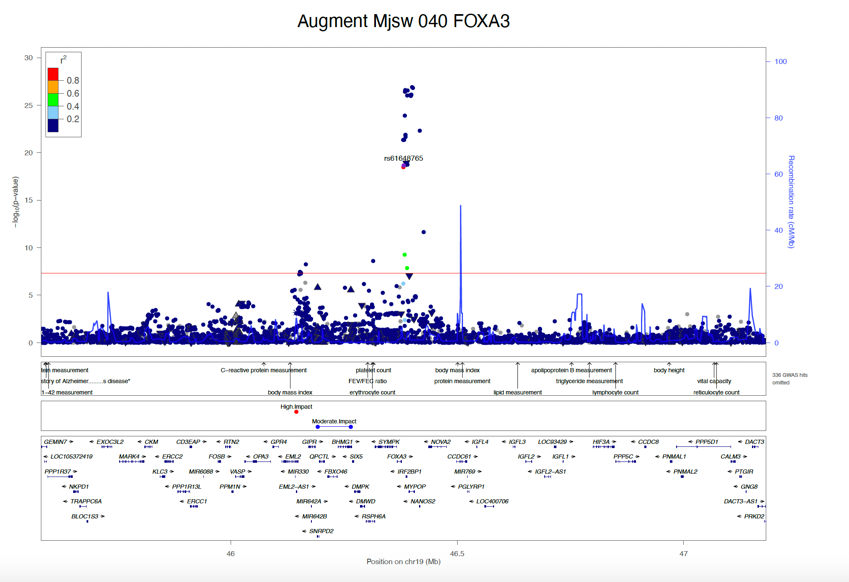

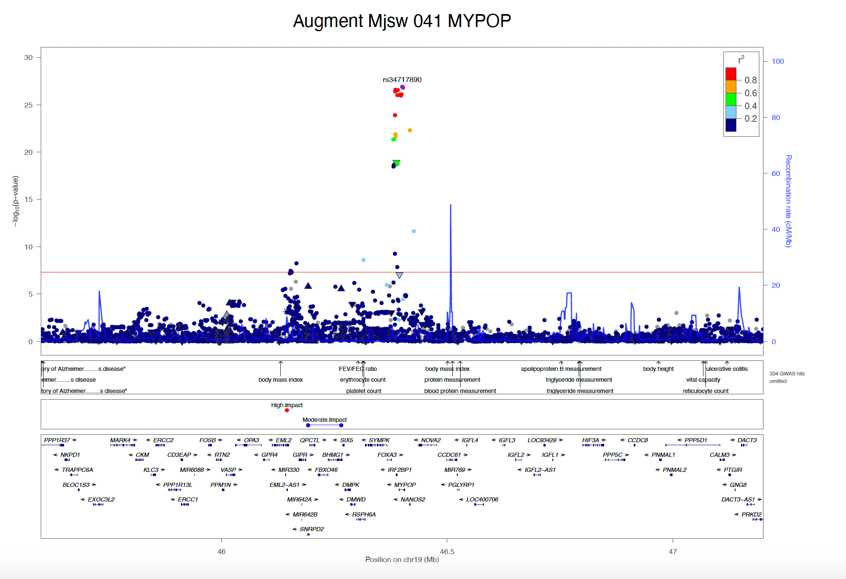

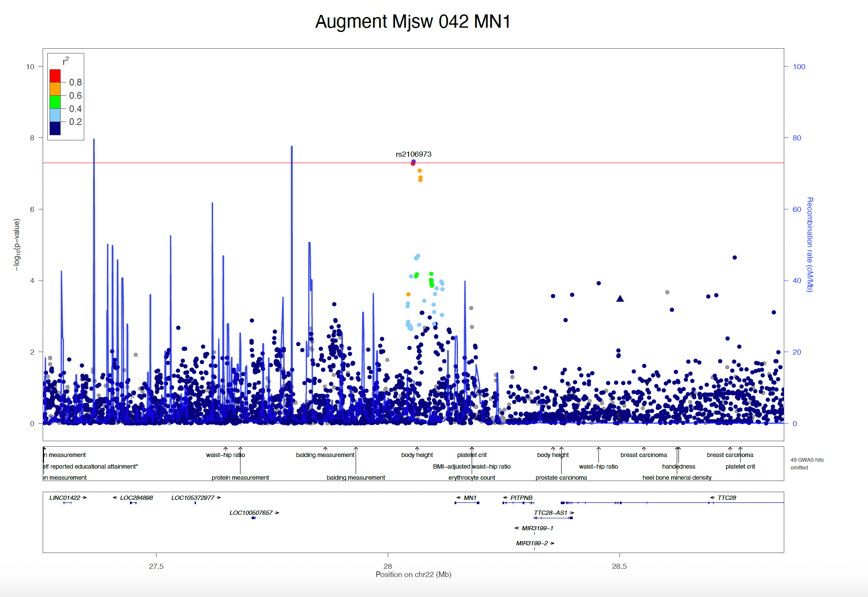

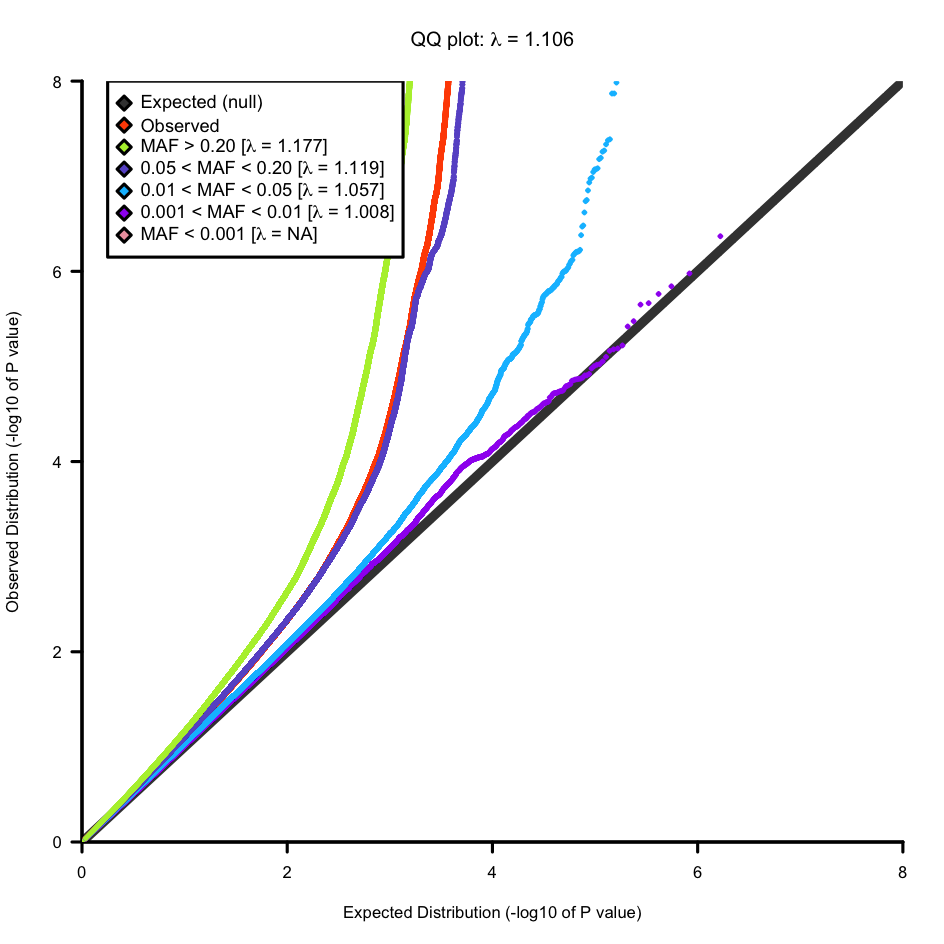

### Supplementary Figure 2: QQ plot of minimum joint space meta-analysis

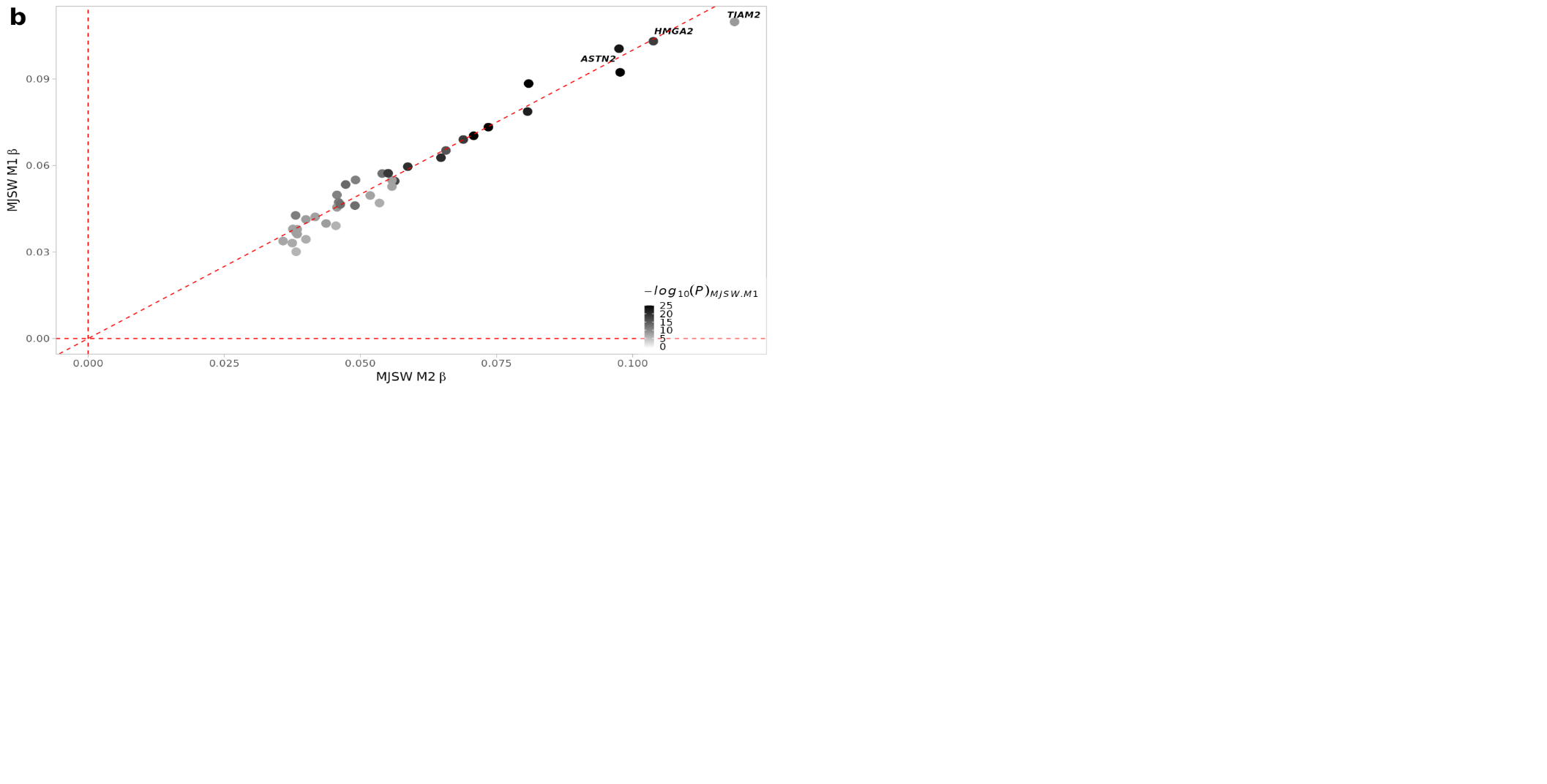

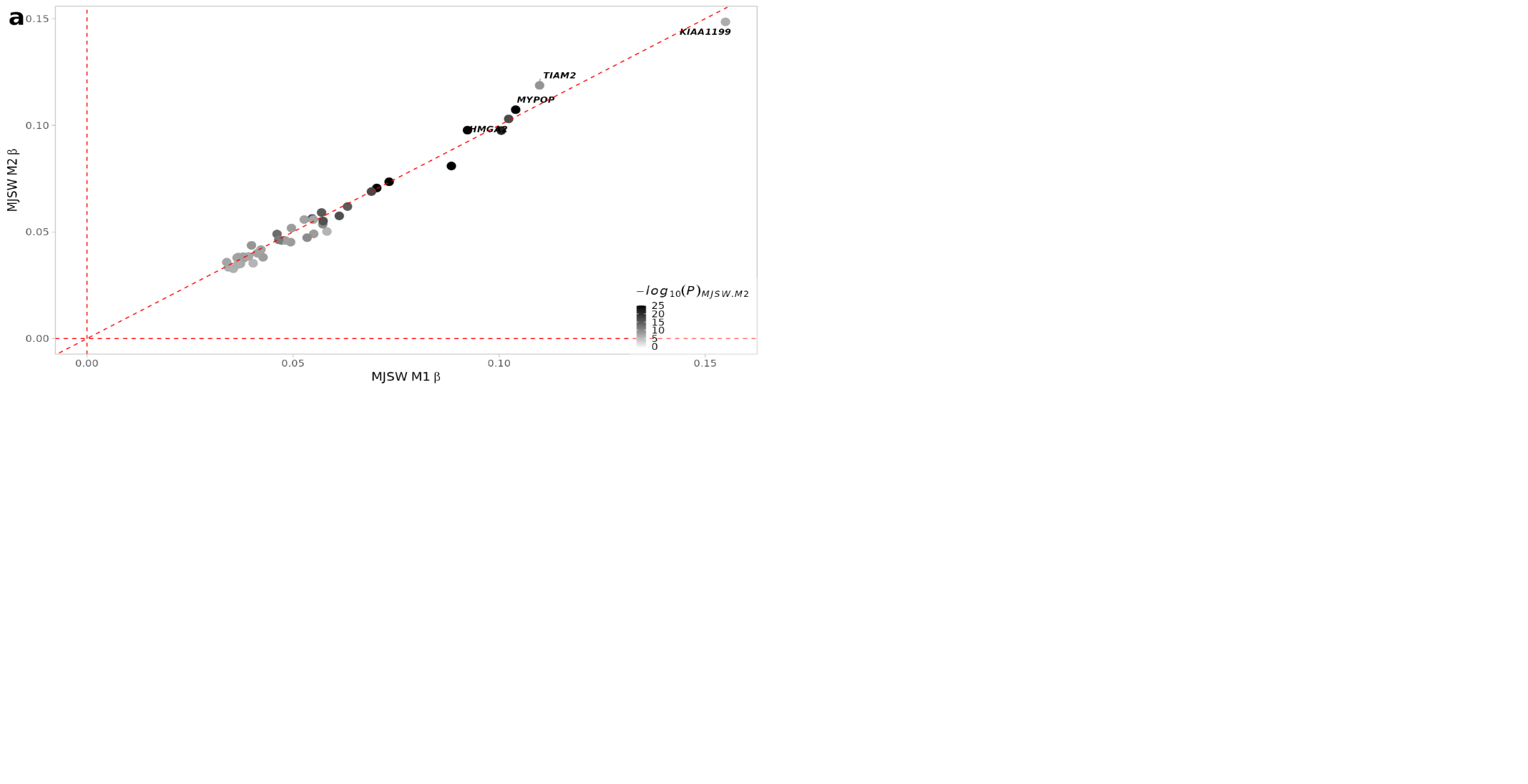

### Supplementary figure 3: Look up of a) Model 1 (adjusted for age and sex) mJSW meta-analysis results in Model 2 mJSW meta analysis results (adjusted for height) b) Look up of Model 2 results in Model 1

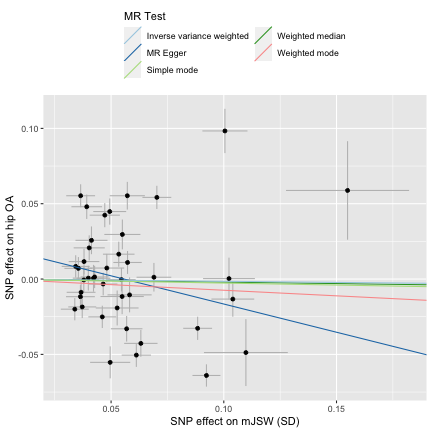

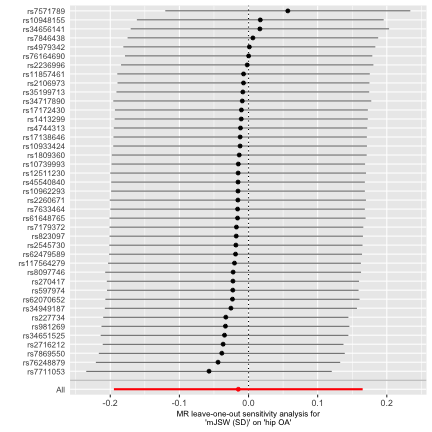

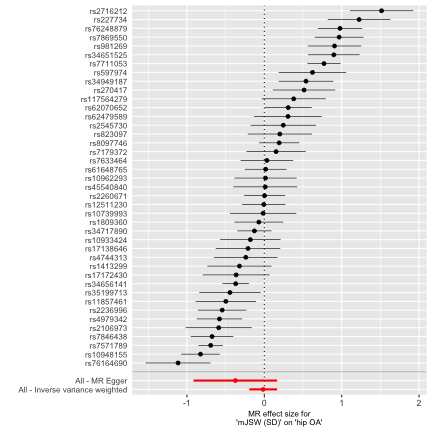

### Supplementary Figure 4A. Exposure – mJSW, Outcome – hip osteoarthritis. Left – Mendelian randomisation plot comparing the 5 different methods. Centre – leave one out analysis. Right – Single SNP analysis.

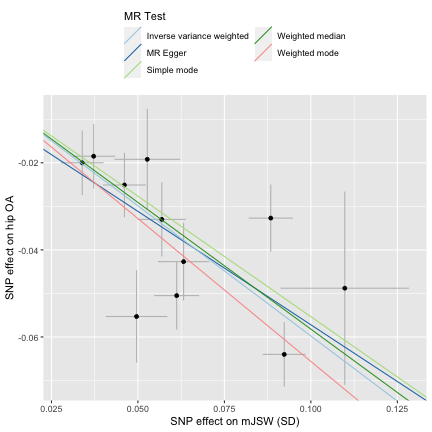

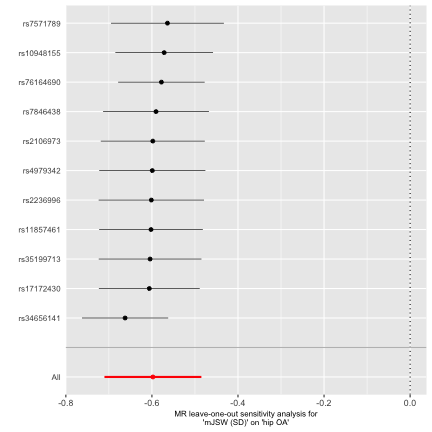

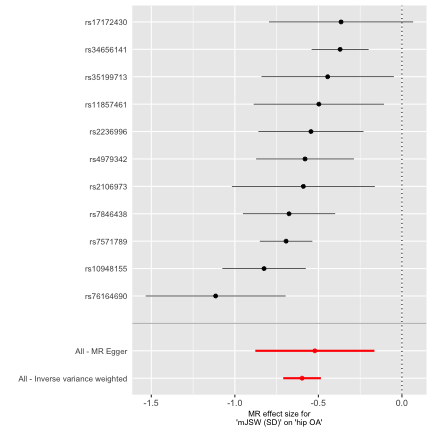

### Supplementary Figure 4B. Exposure – Cluster one mJSW SNPs, Outcome – hip osteoarthritis. Left – Mendelian randomisation plot comparing the 5 different methods. Centre – leave one out analysis. Right – Single SNP analysis.

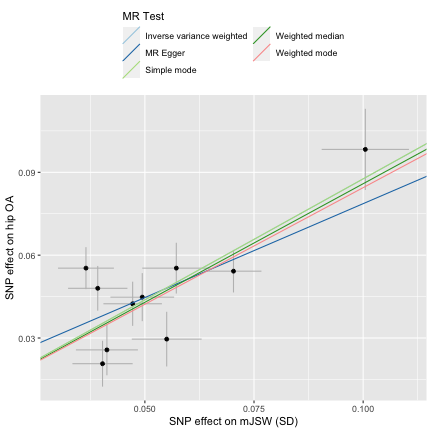

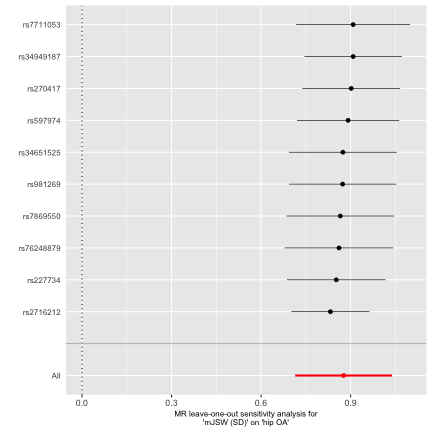

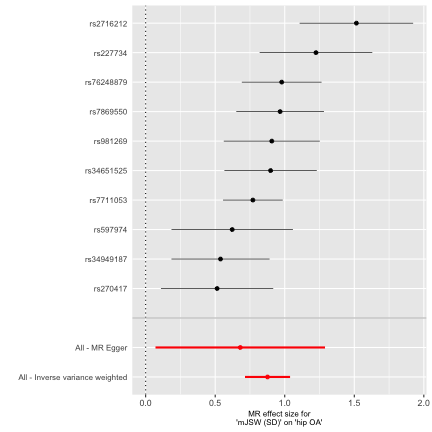

### Supplementary Figure 4C. Exposure – Cluster two mJSW SNPs, Outcome – hip osteoarthritis. Left – Mendelian randomisation plot comparing the 5 different methods. Centre – leave one out analysis. Right – Single SNP analysis.

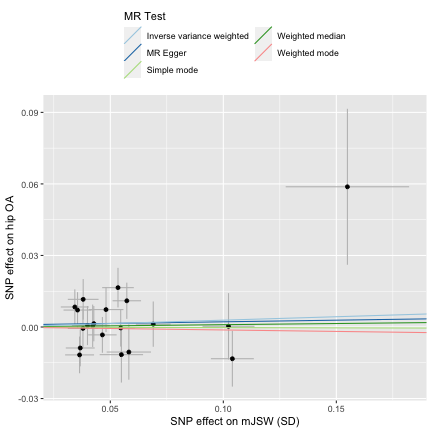

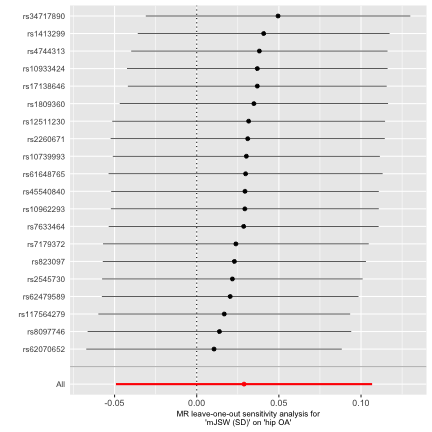

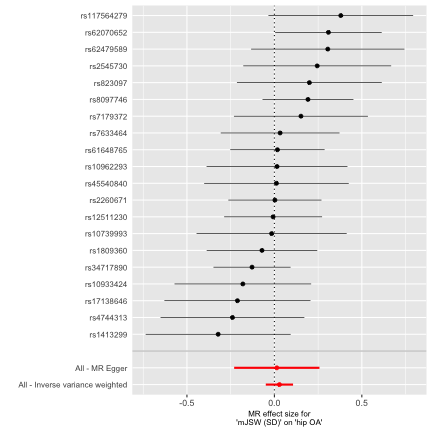

### Supplementary Figure 4D. Exposure –Cluster three mJSW SNPs, Outcome – hip osteoarthritis. Left – Mendelian randomisation plot comparing the 5 different methods. Centre – leave one out analysis. Right – Single SNP analysis.

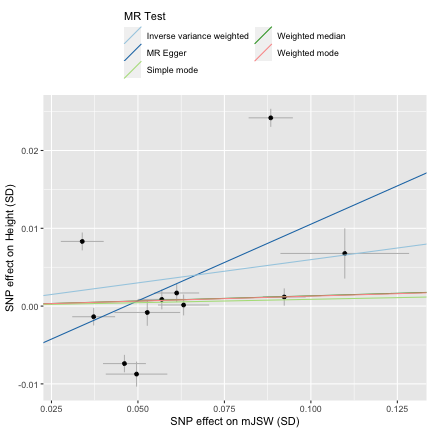

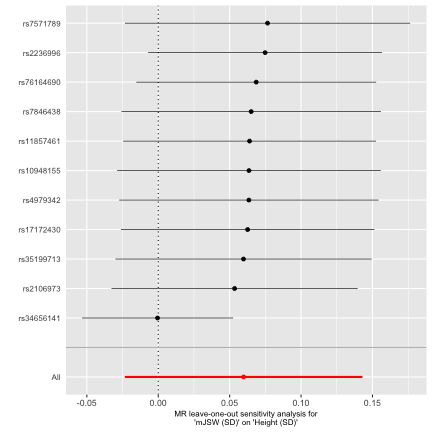

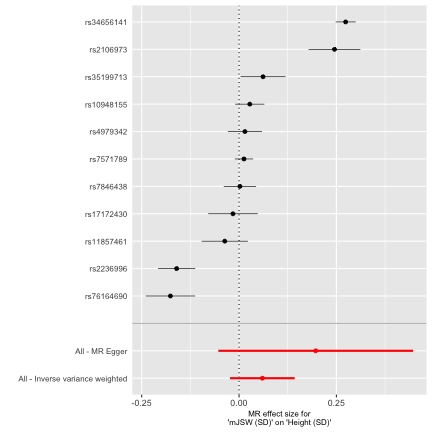

### Supplementary Figure 4E. Exposure – Cluster one mJSW SNPs, Outcome – height. Left – Mendelian randomisation plot comparing the 5 different methods. Centre – leave one out analysis. Right – Single SNP analysis.

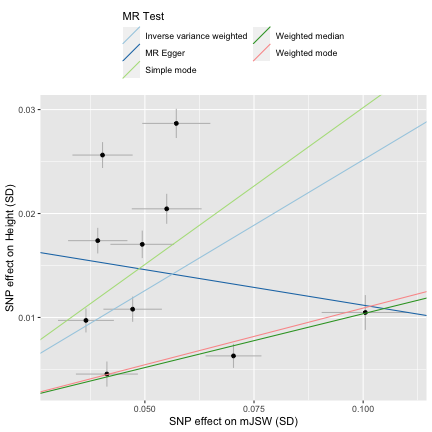

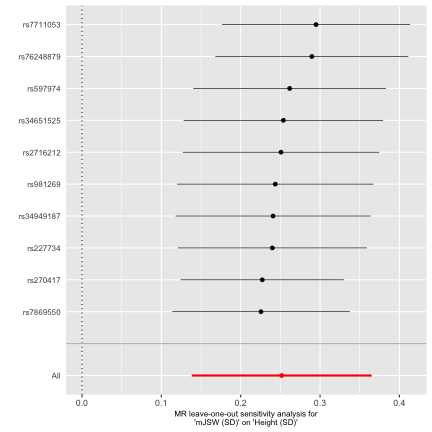

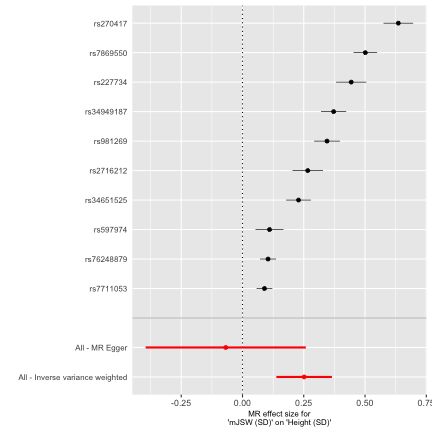

### Supplementary Figure 4F. Exposure – Cluster two mJSW SNPs, Outcome – height. Left – Mendelian randomisation plot comparing the 5 different methods. Centre – leave one out analysis. Right – Single SNP analysis.
