## Supplementary Tables for "Hip joint space width is causally related to hip osteoarthritis risk via distinct protective and susceptibility mechanisms: findings from a genome-wide association study meta-analysis"

### Supplementary Table 1. Descriptive statistics for each cohort included in the meta-analysis.

|  | **UKB** | **RS1** | **RS2** | **MrOS** | **SOF** | **Combined** |
| --- | --- | --- | --- | --- | --- | --- |
| Age - mean in years (range) | 63.8 (45-82) | 64.0 (55-86) | 64.5 (55-93) | 77.6 (69 - 97) | 70.7 (65-91) | 65.1 (45-97) |
| Height - mean in cm (range) | 170.2 (135-204) | 169.0 (141 -198) | 168.8(142-203) | 173.7 (150-198) | 159.3 (139-179) | 169.7 (135-204) |
| Weight - mean in kg (range) | 75.4 (34-171) | 74.8 (41-131) | 77.6 (36-127) | 82.6 (47-141) | 68.3 (40-129) | 75.5 (34-171) |
| Sex - male frequency (percent) | 18,317 (48%) | 2,075 (43%) | 801 (46%) | 3,236 (100%) | 0 (0%) | 24,429 (48%) |
| Sex - female frequency (percent) | 19,858 (52%) | 2,749 (57%) | 930 (54%) | 0 (0%) | 2,779 (100%) | 26,316 (52%) |
| mJSW -mean in mm (range) | 2.89 (0.0-5.9) | 3.8 (0.0-7.0) | 4.38 (0.5-7.0) | 3.27 (0.0-7.4) | 2.81 (0.0-6.0) | 3.05 (0.0-7.4) |
| **N** | 38,175 | 4,824 | 1,731 | 3,236 | 2,779 | 50,745 |

Abbreviations: UKB - UK Biobank, RS - Rotterdam Study, MrOS - Osteoporotic Fractures in Men Study, SOF - study of osteoporotic fractures, cm – centimetre, kg – kilogram, mm – millimetre, N – total number

### Supplementary Table 2: COJO results for minimum joint space width meta-analysis

| **RSID** | **C.GENE** | **EA** | **NEA** | **EAF** | **BETA** | **SE** | **P** | **DIR** | **HET.I2** | **N** | **EAF.J** | **BETA.J** | **SE.J** | **P.J** | **N.J** | **LD.R** | **T.VAR** | **T.DIST** | **T.GENE** | **PMID** | **TRAIT** | **STATUS** |
| --- | --- | --- | --- | --- | --- | --- | --- | --- | --- | --- | --- | --- | --- | --- | --- | --- | --- | --- | --- | --- | --- | --- |
| rs823097 | *NUCKS1* | A | G | 0.43 | -0.04 | 0.01 | 1.35E-08 | ----- | 0 | 50745 | 0.42 | -0.04 | 0.01 | 1.04E-08 | 52420 | 0 | . | . | . | - | - | NOVEL |
| rs7571789 | *TGFA* | T | C | 0.48 | -0.09 | 0.01 | 2.62E-50 | ----- | 14.3 | 50745 | 0.48 | -0.09 | 0.01 | 6.45E-50 | 51189 | 0 | rs2862851 | 1991 | TGFA | 27701424 | Hip-mJSW | KNOWN |
| rs10933424 | *NGEF* | T | C | 0.89 | 0.06 | 0.01 | 2.65E-09 | +++++ | 26.6 | 50745 | 0.89 | 0.06 | 0.01 | 2.91E-09 | 50623 | 0 | . | . | . | - | - | NOVEL |
| rs7633464 | *DCBLD2* | A | G | 0.48 | 0.04 | 0.01 | 5.71E-12 | +++++ | 0 | 50745 | 0.48 | 0.04 | 0.01 | 5.82E-12 | 51349 | 0 | . | . | . | - | - | NOVEL |
| rs2236996 | *SLBP* | A | G | 0.48 | 0.05 | 0.01 | 1.15E-13 | +++++ | 0 | 50745 | 0.48 | 0.05 | 0.01 | 1.07E-13 | 51319 | 0 | . | . | . | - | - | NOVEL |
| rs981269 | *RAB28* | T | C | 0.77 | 0.05 | 0.01 | 1.55E-11 | +++++ | 0 | 50745 | 0.77 | 0.05 | 0.01 | 1.34E-11 | 51542 | 0 | . | . | . | - | - | NOVEL |
| rs12511230 | *HHIP* | A | T | 0.60 | 0.05 | 0.01 | 5.66E-18 | +++++ | 0 | 50745 | 0.61 | 0.05 | 0.01 | 4.70E-18 | 51825 | 0 | . | . | . | - | - | NOVEL |
| rs7711053 | *PIK3R1* | A | G | 0.38 | -0.07 | 0.01 | 3.74E-28 | ----- | 0 | 50745 | 0.38 | -0.07 | 0.01 | 5.24E-28 | 50900 | 0 | rs10471753 | 3668 | PIK3R1 | 27701424 | Hip-mJSW | KNOWN |
| rs2545730 | *RGMB* | A | G | 0.52 | -0.03 | 0.01 | 3.52E-08 | ----- | 0 | 50745 | 0.52 | -0.03 | 0.01 | 3.19E-08 | 51342 | 0 | . | . | . | - | - | NOVEL |
| rs17138646 | *AQPEP* | T | G | 0.88 | 0.05 | 0.01 | 1.28E-08 | ++-++ | 15.2 | 50745 | 0.88 | 0.05 | 0.01 | 1.08E-08 | 51282 | 0 | . | . | . | - | - | NOVEL |
| rs270417 | *BMP6* | T | C | 0.72 | 0.04 | 0.01 | 4.69E-09 | ++++- | 0 | 50745 | 0.72 | 0.04 | 0.01 | 5.26E-09 | 51365 | 0 | . | . | . | - | - | NOVEL |
| rs10948155 | *SUPT3H* | T | C | 0.65 | 0.06 | 0.01 | 6.07E-21 | +++++ | 22 | 50745 | 0.65 | 0.06 | 0.01 | 5.09E-21 | 51309 | 0 | rs10948155 | 0 | SUPT3H | 27701424 | Hip-mJSW | KNOWN |
| rs35199713 | *TIAM2* | A | G | 0.03 | -0.11 | 0.02 | 3.25E-09 | ----- | 0 | 50745 | 0.03 | -0.11 | 0.02 | 3.61E-09 | 50226 | 0 | . | . | . | - | - | NOVEL |
| rs17172430 | *EGFR* | A | G | 0.12 | 0.05 | 0.01 | 2.51E-08 | +++++ | 7.2 | 50745 | 0.12 | 0.05 | 0.01 | 2.93E-08 | 50110 | 0 | . | . | . | - | - | NOVEL |
| rs62479589 | *CALU* | A | G | 0.38 | -0.04 | 0.01 | 2.42E-08 | -+??- | 28.7 | 44190 | 0.39 | -0.04 | 0.01 | 2.32E-08 | 45128 | 0 | . | . | . | - | - | NOVEL |
| rs7846438 | *C8orf34* | A | G | 0.77 | 0.06 | 0.01 | 9.65E-18 | ++-++ | 20 | 50745 | 0.77 | 0.06 | 0.01 | 1.41E-17 | 50940 | 0 | . | . | . | - | - | NOVEL |
| rs10962293 | *C9orf92* | T | C | 0.29 | -0.04 | 0.01 | 6.58E-09 | ----- | 10 | 50745 | 0.28 | -0.04 | 0.01 | 7.44E-09 | 50393 | 0 | . | . | . | - | - | NOVEL |
| rs4744313 | *PTPDC1* | T | C | 0.63 | 0.04 | 0.01 | 1.01E-08 | +++++ | 0 | 50745 | 0.63 | 0.04 | 0.01 | 1.81E-08 | 51553 | -0.0273 | . | . | . | - | - | NOVEL |
| rs10739993 | *FANCC* | T | C | 0.59 | -0.04 | 0.01 | 1.79E-08 | --??- | 0 | 44190 | 0.59 | -0.04 | 0.01 | 4.29E-08 | 45436 | -0.00567 | . | . | . | - | - | NOVEL |
| rs1413299 | *COL15A1* | T | G | 0.37 | 0.04 | 0.01 | 1.39E-08 | +++++ | 7.5 | 50745 | 0.36 | 0.04 | 0.01 | 1.23E-08 | 51582 | 0 | . | . | . | - | - | NOVEL |
| rs4979342 | *COL27A1* | T | C | 0.27 | -0.06 | 0.01 | 3.88E-16 | ----- | 0 | 50745 | 0.27 | -0.06 | 0.01 | 1.12E-16 | 51067 | 0.005683 | . | . | . | - | - | NOVEL |
| rs7869550 | *PAPPA* | A | G | 0.80 | 0.06 | 0.01 | 2.01E-13 | ++++- | 25.7 | 50745 | 0.80 | 0.06 | 0.01 | 1.46E-12 | 50658 | -0.01829 | . | . | . | - | - | NOVEL |
| rs76248879 | *ASTN2* | A | T | 0.87 | 0.10 | 0.01 | 1.07E-23 | ++??+ | 21.9 | 44190 | 0.88 | 0.07 | 0.01 | 5.14E-11 | 42349 | 0.385652 | . | . | . | - | - | NOVEL |
| rs34687269 | *ASTN2* | A | T | 0.52 | 0.07 | 0.01 | 3.96E-32 | +++++ | 0 | 50745 | 0.53 | 0.06 | 0.01 | 1.60E-17 | 51277 | 0 | . | . | . | - | - | NOVEL |
| rs597974 | *SURF6* | A | G | 0.68 | 0.04 | 0.01 | 6.82E-09 | ++??+ | 4.6 | 44190 | 0.68 | 0.04 | 0.01 | 6.07E-09 | 45105 | 0 | . | . | . | - | - | NOVEL |
| rs76164690 | *EPC1* | T | G | 0.86 | 0.05 | 0.01 | 2.37E-08 | +++++ | 19.7 | 50745 | 0.86 | 0.05 | 0.01 | 2.53E-08 | 51377 | 0 | . | . | . | - | - | NOVEL |
| rs34651525 | *TEAD1* | A | T | 0.69 | -0.05 | 0.01 | 1.53E-12 | ----- | 19 | 50745 | 0.68 | -0.05 | 0.01 | 1.90E-12 | 50979 | 0 | . | . | . | - | - | NOVEL |
| rs45540840 | *PHLDB1* | A | G | 0.22 | -0.04 | 0.01 | 1.87E-08 | ----- | 0 | 50745 | 0.22 | -0.04 | 0.01 | 1.86E-08 | 50934 | 0 | . | . | . | - | - | NOVEL |
| rs2260671 | *HMGA2* | A | G | 0.08 | 0.10 | 0.01 | 8.19E-19 | +++++ | 0 | 50745 | 0.08 | 0.10 | 0.01 | 6.17E-19 | 51625 | 0 | . | . | . | - | - | NOVEL |
| rs11857461 | *ALDH1A2* | T | C | 0.49 | -0.04 | 0.01 | 2.26E-09 | ----- | 0 | 50745 | 0.50 | -0.04 | 0.01 | 2.94E-09 | 51268 | 0.001472 | . | . | . | - | - | NOVEL |
| rs7179372 | *SMAD6* | A | G | 0.20 | -0.05 | 0.01 | 4.12E-10 | ----- | 0 | 50745 | 0.20 | -0.05 | 0.01 | 4.97E-10 | 51364 | 0.002142 | . | . | . | - | - | NOVEL |
| rs1809360 | *SKOR1* | T | C | 0.57 | -0.05 | 0.01 | 1.12E-13 | ----- | 0 | 50745 | 0.57 | -0.05 | 0.01 | 2.48E-13 | 50580 | 0 | . | . | . | - | - | NOVEL |
| rs117564279 | *CEMIP* | A | G | 0.02 | 0.15 | 0.03 | 1.35E-08 | +++++ | 0 | 50745 | 0.01 | 0.15 | 0.03 | 1.72E-08 | 38219 | -0.00563 | . | . | . | - | - | NOVEL |
| rs34949187 | *ACAN* | A | G | 0.18 | -0.06 | 0.01 | 7.65E-12 | ----- | 0 | 50745 | 0.19 | -0.05 | 0.01 | 7.68E-12 | 51055 | 0 | . | . | . | - | - | NOVEL |
| rs62070652 | *ATAD5* | T | C | 0.27 | -0.05 | 0.01 | 2.62E-14 | ----- | 26.4 | 50745 | 0.27 | -0.05 | 0.01 | 2.45E-14 | 51415 | 0 | . | . | . | - | - | NOVEL |
| rs227734 | *NOG* | T | C | 0.30 | 0.04 | 0.01 | 7.44E-09 | ++++- | 28.9 | 50745 | 0.30 | 0.04 | 0.01 | 8.27E-09 | 51162 | 0 | . | . | . | - | - | NOVEL |
| rs2716212 | *MAP2K6* | A | G | 0.61 | -0.04 | 0.01 | 1.18E-08 | --+-- | 5 | 50745 | 0.62 | -0.04 | 0.01 | 1.19E-08 | 50772 | 0 | . | . | . | - | - | NOVEL |
| rs8097746 | *DYM* | T | C | 0.59 | 0.06 | 0.01 | 9.12E-20 | +++++ | 0 | 50745 | 0.59 | 0.06 | 0.01 | 1.01E-19 | 51338 | 0 | . | . | . | - | - | NOVEL |
| rs34656141 | *AP3D1* | T | C | 0.40 | 0.09 | 0.01 | 1.42E-43 | +++++ | 12.1 | 50745 | 0.40 | 0.09 | 0.01 | 3.08E-43 | 50044 | 0 | rs11880992 | 18175 | DOT1L | 27701424 | Hip-mJSW | KNOWN |
| rs61648765 | *FOXA3* | C | G | 0.78 | 0.07 | 0.01 | 2.11E-19 | +++++ | 0 | 50745 | 0.79 | 0.06 | 0.01 | 1.48E-12 | 48558 | 0.18768 | . | . | . | - | - | NOVEL |
| rs34717890 | *MYPOP* | T | C | 0.12 | 0.10 | 0.01 | 1.24E-27 | +++++ | 29.7 | 50745 | 0.12 | 0.09 | 0.01 | 3.10E-21 | 51341 | 0 | . | . | . | - | - | NOVEL |
| rs2106973 | *MN1* | A | G | 0.48 | -0.03 | 0.01 | 4.63E-08 | ----+ | 18 | 50745 | 0.48 | -0.03 | 0.01 | 4.60E-08 | 51312 | 0 | . | . | . | - | - | NOVEL |

| Legend |  |
| --- | --- |
| **RSID** | Reference SNP cluster ID |
| **C.GENE** | Closest gene to reported variant |
| **EA** | Effect allele |
| **NEA** | Non-effect allele |
| **EAF** | Effect allele frequency |
| **BETA** | Per allele effect in standard deviations of **mJSW** |
| **SE** | Standard error of the β |
| **P** | Strength of evidence against the null hypothesis of no association between variant and outcome |
| **DIR** | Direction of effect in each participating cohort |
| **HET.I2** | Heterogeneity |
| **N** | Sample size |
| **EAF.J** | Frequency of the effect allele in the reference sample used for conditional and joint genome-wide association analysis |
| **MAF.J** | Minor allele frequency of variant in reference databases used for conditional and joint genome-wide association analysis |
| **BETA.J** | Per allele effect estimated from joint analysis of conditionally associated snps in standard deviations of outcome |
| **SE.J** | Standard error of β.J |
| **P.J** | Strength of evidence against the null hypothesis of no association between the variant and outcome as estimated by conditional and joint genome-wide association analysis (i.e. P-value) |
| **N.J** | Estimated effective sample size |
| **LD.R** | LD correlation between the SNP i and SNP i + 1 |
| **T.VAR** | Closest published mJSW associated variant |
| **T.DIST** | Distance between conditionally independent mJSW associated variant and closest previously published mJSW associated variant |
| **T.GENE** | Closest gene to published mJSW associated variant |
| **PMID** | Publication ID |
| **TRAIT** | Trait previously published |

### Supplementary Table 3: Genetic correlations - mJSW meta-analysis compared against various traits

| Trait | rg | se | lci | uci | z | p | h2 |
| --- | --- | --- | --- | --- | --- | --- | --- |
| Height | 0.28 | 0.03 | 0.22 | 0.33 | 9.74 | 2.04E-22 | 0.50 |
| Body Mass Index | 0.06 | 0.03 | 0.01 | 0.12 | 2.22 | 0.03 | 0.21 |
| Hip Osteoarthritis | 0.10 | 0.06 | -0.01 | 0.21 | 1.75 | 0.14 | 0.08 |

rg - correlation coefficient, se - standard error, lci - lower 95% confidence interval, uci - upper 95% confidence interval, p - p-value, h2 - heritability of trait

### Supplementary Table 4: Colocalisation analysis to assess if mJSW and HOA GWAS signals share common genetic causal variant in a given region with the posterior probability (PP) reported (H4: both traits are associated, and share a single causal variant).

| RSID | CHR | BP | C.GENE | EA | NEA | Cluster | Height Beta | Height P | Height Coloc (PP4) |
| --- | --- | --- | --- | --- | --- | --- | --- | --- | --- |
| rs7571789 | 2 | 70714793 | *TGFA* | C | T | 1 | 0.00 | 0.11 | 0.00 |
| rs2236996 | 4 | 1703646 | *SLBP* | A | G | 1 | -0.01 | 6.00E-07 | 0.00 |
| rs10948155 | 6 | 44687957 | *SUPT3H* | T | C | 1 | 0.00 | 0.78 | 0.00 |
| rs35199713 | 6 | 155415593 | *TIAM2* | G | A | 1 | 0.01 | 0.02 | 0.04 |
| rs17172430 | 7 | 55122650 | *EGFR* | A | G | 1 | 0.00 | 0.93 | 0.00 |
| rs7846438 | 8 | 69578824 | *C8orf34* | A | G | 1 | 0.00 | 0.82 | 0.00 |
| rs4979342 | 9 | 116905618 | *COL27A1* | C | T | 1 | 0.002 | 0.24 | 0.00 |
| rs76164690 | 10 | 32590362 | *EPC1* | T | G | 1 | -0.008 | 5.90E-06 | 0.79 |
| rs11857461 | 15 | 58319690 | *ALDH1A2* | C | T | 1 | -0.001 | 0.47 | 0.00 |
| rs34656141 | 19 | 2158228 | *AP3D1* | T | C | 1 | 0.025 | 2.00E-76 | 0.00 |
| rs2106973 | 22 | 28055460 | *MN1* | G | A | 1 | 0.010 | 1.70E-13 | 0.98 |
| rs981269 | 4 | 12897698 | *RAB28* | T | C | 2 | 0.018 | 5.10E-30 | 0.00 |
| rs7711053 | 5 | 67822620 | *PIK3R1* | G | A | 2 | 0.005 | 1.90E-04 | 0.58 |
| rs270417 | 6 | 7729614 | *BMP6* | T | C | 2 | 0.029 | 7.10E-87 | 0.99 |
| rs7869550 | 9 | 119134796 | *PAPPA* | A | G | 2 | 0.030 | 1.40E-74 | 0.00 |
| rs76248879* | 9 | 119325659 | *ASTN2* | A | T | 2 | *proxy SNP not found | | 0.38 |
| rs597974 | 9 | 136144297 | *SURF6* | A | G | 2 | -0.004 | 0.004 | 0.07 |
| rs34651525 | 11 | 12846729 | *TEAD1* | T | A | 2 | 0.011 | 5.40E-14 | 0.33 |
| rs34949187 | 15 | 89386652 | *ACAN* | G | A | 2 | 0.022 | 1.40E-37 | 0.00 |
| rs2716212 | 17 | 67503653 | *MAP2K6* | G | A | 2 | 0.010 | 3.40E-14 | 0.97 |
| rs227734 | 17 | 54767470 | *NOG* | T | C | 2 | 0.018 | 2.80E-37 | 0.02 |
| rs823097 | 1 | 205681370 | *NUCKS1* | G | A | 3 | 0.013 | 8.20E-23 | 0.14 |
| rs10933424 | 2 | 233872408 | *NGEF* | T | C | 3 | 0.016 | 1.20E-14 | 0.98 |
| rs7633464 | 3 | 98715823 | *DCBLD2* | A | G | 3 | 0.008 | 4.90E-10 | 0.01 |
| rs12511230 | 4 | 145471245 | *HHIP* | A | T | 3 | -0.012 | 2.70E-18 | 0.00 |
| rs2545730 | 5 | 98109985 | *RGMB* | G | A | 3 | -0.002 | 0.11 | 0.00 |
| rs17138646 | 5 | 115346245 | *AQPEP* | T | G | 3 | 0.003 | 0.10 | 0.00 |
| rs62479589 | 7 | 128406506 | *CALU* | G | A | 3 | -0.005 | 0.001 | 0.21 |
| rs4744313 | 9 | 96846061 | *PTPDC1* | T | C | 3 | -0.002 | 0.22 | 0.00 |
| rs10962293 | 9 | 16136648 | *C9orf92* | C | T | 3 | 0.000 | 0.82 | 0.00 |
| rs1413299 | 9 | 101761241 | *COL15A1* | T | G | 3 | -0.011 | 5.30E-17 | 0.36 |
| rs10739993 | 9 | 97982669 | *FANCC* | C | T | 3 | -0.004 | 0.001 | 0.02 |
| rs45540840 | 11 | 118486110 | *PHLDB1* | G | A | 3 | 0.006 | 3.60E-04 | 0.00 |
| rs2260671 | 12 | 66174909 | *HMGA2* | A | G | 3 | 0.009 | 3.30E-04 | 0.00 |
| rs1809360 | 15 | 68189737 | *SKOR1* | C | T | 3 | 0.000 | 0.84 | 0.00 |
| rs117564279 | 15 | 81224038 | *CEMIP* | A | G | 3 | 0.002 | 0.67 | 0.01 |
| rs7179372 | 15 | 67036441 | *SMAD6* | G | A | 3 | 0.011 | 2.70E-11 | 0.00 |
| rs62070652 | 17 | 29221277 | *ATAD5* | C | T | 3 | 0.041 | 1.60E-166 | 0.99 |
| rs8097746 | 18 | 46640782 | *DYM* | T | C | 3 | 0.020 | 4.80E-51 | 0.97 |
| rs34717890 | 19 | 46400443 | *MYPOP* | T | C | 3 | -0.003 | 0.18 | 0.00 |
| rs61648765 | 19 | 46381864 | *FOXA3* | C | G | 3 | 0.006 | 4.40E-04 | 0.00 |
| rs34687269 | 9 | 119484132 | *ASTN2* | A | T | Pal | 0.013 | 6.20E-23 | 0.17 |

| **CHR** | Chromosome |
| --- | --- |
| **BP** | Base position |
| **C.GENE** | Closest gene to reported variant |
| **EA** | Effect allele |
| **NEA** | Non-effect allele |
| **EAF** | Effect allele frequency |

### Supplementary Table 5: Colocalisation with GTEx signals for all Cluster 1 SNPs

| SNP | GTEx Gene (Tissue) | Coloc (PP4) |
| --- | --- | --- |
| rs7571789 | TGFA (Fibroblasts) | 0.20 |
|  | TGFA (Brain Amygdala) | 0.95 |
| rs10948155 | SUPT3H (Fibroblasts) | 0.11 |
|  | SUPT3H (Brain - basal ganglia) | 0.91 |
| rs76164690 | EPC1 (Fibroblasts) | 0.00 |
| rs7846438 | N/A | N/A |
| rs34656141 | AP3D1 (Fibroblasts) | 0.00 |
|  | DOT1L (Fibroblasts) | 0.00 |
|  | AMH (Fibroblasts) | 0.00 |
| rs4979342 | COL27A1 (Fibroblasts) | 0.00 |
| rs2236996 | FAM53A (Thyroid) | 0.00 |
|  | FGFR3 (Tibial Nerve) | 0.00 |
|  | TMEM129 (Fibroblasts) | 0.00 |
|  | TACC3 (Skeletal Muscle) | 0.00 |
|  | SLBP (Skeletal muscle) | 0.00 |
| rs2106973 | RP1-213J1P (Skeletal muscle) | 0.00 |
|  | MN1 (Tibial Artery) | 0.07 |
| rs11857461 | ALDH1A2 (Fibroblasts) | 0.02 |
| rs35199713 | TIAM2 (Tibial Artery) | 0.00 |
| rs17172430 | N/A | N/A |

LocusFocus was used to conduct colocalisation with all genes within a 200kb area of the sentinel SNP. This was repeated in all tissues available in GTEx. Fibroblasts and the tissues with the highest expressions are reported here.

### Supplementary Table 6. RegulomeDB output for each independent mJSW SNP.

| chrom | start | end | rsids | probability | ranking | ChIP | DNase | Footprint | Footprint_matched | IC_matched_max | IC_max | PWM | PWM_matched | QTL |
| --- | --- | --- | --- | --- | --- | --- | --- | --- | --- | --- | --- | --- | --- | --- |
| chr5 | 67822619 | 67822620 | rs7711053 | 0.97 | 5 | FALSE | TRUE | FALSE | FALSE | 0 | 0.53 | TRUE | FALSE | FALSE |
| chr19 | 2158227 | 2158228 | rs34656141 | 0.94 | 5 | TRUE | FALSE | FALSE | FALSE | 0 | 1.90 | TRUE | FALSE | FALSE |
| chr1 | 205681369 | 205681370 | rs823097 | 0.81 | 3a | TRUE | TRUE | FALSE | FALSE | 0 | 0.26 | TRUE | FALSE | FALSE |
| chr15 | 67036440 | 67036441 | rs7179372 | 0.76 | 2b | TRUE | TRUE | TRUE | FALSE | 0 | 0.88 | TRUE | FALSE | FALSE |
| chr6 | 7729613 | 7729614 | rs270417 | 0.71 | 2b | TRUE | TRUE | TRUE | FALSE | 0 | 1.63 | TRUE | FALSE | FALSE |
| chr9 | 101761240 | 101761241 | rs1413299 | 0.70 | 4 | TRUE | TRUE | TRUE | FALSE | 0 | 0.00 | FALSE | FALSE | FALSE |
| chr11 | 118486109 | 118486110 | rs45540840 | 0.70 | 4 | TRUE | TRUE | TRUE | FALSE | 0 | 0.00 | FALSE | FALSE | FALSE |
| chr2 | 233872407 | 233872408 | rs10933424 | 0.61 | 4 | TRUE | TRUE | FALSE | FALSE | 0 | 0.00 | FALSE | FALSE | FALSE |
| chr4 | 12897697 | 12897698 | rs981269 | 0.61 | 4 | TRUE | TRUE | FALSE | FALSE | 0 | 0.00 | FALSE | FALSE | FALSE |
| chr5 | 98109984 | 98109985 | rs2545730 | 0.61 | 4 | TRUE | TRUE | FALSE | FALSE | 0 | 0.00 | FALSE | FALSE | FALSE |
| chr6 | 155415592 | 155415593 | rs35199713 | 0.61 | 4 | TRUE | TRUE | FALSE | FALSE | 0 | 0.00 | FALSE | FALSE | FALSE |
| chr7 | 55122649 | 55122650 | rs17172430 | 0.61 | 4 | TRUE | TRUE | FALSE | FALSE | 0 | 0.00 | FALSE | FALSE | FALSE |
| chr9 | 96846060 | 96846061 | rs4744313 | 0.61 | 4 | TRUE | TRUE | FALSE | FALSE | 0 | 0.00 | FALSE | FALSE | FALSE |
| chr9 | 97982668 | 97982669 | rs10739993 | 0.61 | 4 | TRUE | TRUE | FALSE | FALSE | 0 | 0.00 | FALSE | FALSE | FALSE |
| chr15 | 89386651 | 89386652 | rs34949187 | 0.61 | 4 | TRUE | TRUE | FALSE | FALSE | 0 | 0.00 | FALSE | FALSE | FALSE |
| chr19 | 46381863 | 46381864 | rs61648765 | 0.61 | 4 | TRUE | TRUE | FALSE | FALSE | 0 | 0.00 | FALSE | FALSE | FALSE |
| chr2 | 70714792 | 70714793 | rs7571789 | 0.59 | 5 | TRUE | FALSE | FALSE | FALSE | 0 | 0.00 | FALSE | FALSE | FALSE |
| chr11 | 12846728 | 12846729 | rs34651525 | 0.59 | 5 | TRUE | FALSE | FALSE | FALSE | 0 | 0.00 | FALSE | FALSE | FALSE |
| chr17 | 29221276 | 29221277 | rs62070652 | 0.59 | 5 | TRUE | FALSE | FALSE | FALSE | 0 | 0.00 | FALSE | FALSE | FALSE |
| chr19 | 46400442 | 46400443 | rs34717890 | 0.59 | 5 | TRUE | FALSE | FALSE | FALSE | 0 | 0.00 | FALSE | FALSE | FALSE |
| chr9 | 136144296 | 136144297 | rs597974 | 0.57 | 3a | TRUE | TRUE | FALSE | FALSE | 0 | 1.72 | TRUE | FALSE | FALSE |
| chr17 | 67503652 | 67503653 | rs2716212 | 0.52 | 6 | FALSE | FALSE | FALSE | FALSE | 0 | 1.35 | TRUE | FALSE | FALSE |
| chr18 | 46640781 | 46640782 | rs8097746 | 0.34 | 6 | FALSE | FALSE | FALSE | FALSE | 0 | 1.05 | TRUE | FALSE | FALSE |
| chr4 | 1703645 | 1703646 | rs2236996 | 0.33 | 3a | TRUE | TRUE | FALSE | FALSE | 0 | 1.44 | TRUE | FALSE | FALSE |
| chr15 | 81224037 | 81224038 | rs117564279 | 0.30 | 5 | TRUE | FALSE | FALSE | FALSE | 0 | 0.24 | TRUE | FALSE | FALSE |
| chr4 | 145471244 | 145471245 | rs12511230 | 0.20 | 6 | FALSE | FALSE | FALSE | FALSE | 0 | 0.94 | TRUE | FALSE | FALSE |
| chr3 | 98715822 | 98715823 | rs7633464 | 0.18 | 7 | FALSE | FALSE | FALSE | FALSE | 0 | 0.00 | FALSE | FALSE | FALSE |
| chr5 | 115346244 | 115346245 | rs17138646 | 0.18 | 7 | FALSE | FALSE | FALSE | FALSE | 0 | 0.00 | FALSE | FALSE | FALSE |
| chr7 | 128406505 | 128406506 | rs62479589 | 0.18 | 7 | FALSE | FALSE | FALSE | FALSE | 0 | 0.00 | FALSE | FALSE | FALSE |
| chr9 | 119484131 | 119484132 | rs34687269 | 0.18 | 7 | FALSE | FALSE | FALSE | FALSE | 0 | 0.00 | FALSE | FALSE | FALSE |
| chr10 | 32590361 | 32590362 | rs76164690 | 0.18 | 7 | FALSE | FALSE | FALSE | FALSE | 0 | 0.00 | FALSE | FALSE | FALSE |
| chr12 | 66174908 | 66174909 | rs2260671 | 0.18 | 7 | FALSE | FALSE | FALSE | FALSE | 0 | 0.00 | FALSE | FALSE | FALSE |
| chr15 | 58319689 | 58319690 | rs11857461 | 0.18 | 7 | FALSE | FALSE | FALSE | FALSE | 0 | 0.00 | FALSE | FALSE | FALSE |
| chr22 | 28055459 | 28055460 | rs2106973 | 0.18 | 7 | FALSE | FALSE | FALSE | FALSE | 0 | 0.00 | FALSE | FALSE | FALSE |
| chr9 | 119325658 | 119325659 | rs76248879 | 0.16 | 6 | FALSE | FALSE | FALSE | FALSE | 0 | 1.91 | TRUE | FALSE | FALSE |
| chr6 | 44687956 | 44687957 | rs10948155 | 0.13 | 5 | FALSE | TRUE | FALSE | FALSE | 0 | 0.00 | FALSE | FALSE | FALSE |
| chr9 | 16136647 | 16136648 | rs10962293 | 0.13 | 5 | FALSE | TRUE | FALSE | FALSE | 0 | 0.00 | FALSE | FALSE | FALSE |
| chr9 | 116905617 | 116905618 | rs4979342 | 0.13 | 5 | FALSE | TRUE | FALSE | FALSE | 0 | 0.00 | FALSE | FALSE | FALSE |
| chr9 | 119134795 | 119134796 | rs7869550 | 0.13 | 5 | FALSE | TRUE | FALSE | FALSE | 0 | 0.00 | FALSE | FALSE | FALSE |
| chr15 | 68189736 | 68189737 | rs1809360 | 0.13 | 5 | FALSE | TRUE | FALSE | FALSE | 0 | 0.00 | FALSE | FALSE | FALSE |
| chr17 | 54767469 | 54767470 | rs227734 | 0.13 | 5 | FALSE | TRUE | FALSE | FALSE | 0 | 0.00 | FALSE | FALSE | FALSE |
| chr8 | 69578823 | 69578824 | rs7846438 | 0.00 | 6 | FALSE | FALSE | FALSE | FALSE | 0 | 0.03 | TRUE | FALSE | FALSE |

ChIP - ChIP-seq signal, DNase - DNase-seq signal, IC - information content change, PWM - position-weight matrix for transcription factor binding, QTL - quantitative trait loci

RegulomeDB annotes SNPS with known and predicted regulatory elements in non-coding regions of the genome. A probability and ranking score for each SNP is given. The higher the probability the more likely the SNP is a non-coding regulatory SNP.

### Supplementary Table 7. Colocalisation results examining all 42 indepent mJSW SNPs and their associations with gene cis-eQTL expression in human cartilage and synovium.

| GWASInput | eQTLGene | PP.H0.abf | PP.H1.abf | PP.H2.abf | PP.H3.abf | PP.H4.abf |
| --- | --- | --- | --- | --- | --- | --- |
| Less degraded (healthy) cartilage |  |  |  |  |  |  |
| GWAS_mjsw_rs823097_DSTYK_ENSG00000133059_1MbTSS.txt | DSTYK_ENSG00000133059 | 0.00 | 0.00 | 0.01 | 0.99 | 0.00 |
| GWAS_mjsw_rs10933424_MSL3P1_ENSG00000224287_1MbTSS.txt | MSL3P1_ENSG00000224287 | 0.00 | 0.06 | 0.00 | 0.94 | 0.00 |
| GWAS_mjsw_rs10933424_NGEF_ENSG00000066248_1MbTSS.txt | NGEF_ENSG00000066248 | 0.00 | 0.00 | 0.00 | 0.99 | 0.00 |
| GWAS_mjsw_rs10933424_TRPM8_ENSG00000144481_1MbTSS.txt | TRPM8_ENSG00000144481 | 0.00 | 0.05 | 0.07 | 0.88 | 0.00 |
| GWAS_mjsw_rs2236996_CRIPAK_ENSG00000179979_1MbTSS.txt | CRIPAK_ENSG00000179979 | 0.00 | 0.00 | 0.00 | 1.00 | 0.00 |
| GWAS_mjsw_rs2236996_DGKQ_ENSG00000145214_1MbTSS.txt | DGKQ_ENSG00000145214 | 0.00 | 0.02 | 0.00 | 0.98 | 0.00 |
| GWAS_mjsw_rs2236996_FAM53A_ENSG00000174137_1MbTSS.txt | FAM53A_ENSG00000174137 | 0.00 | 0.02 | 0.00 | 0.76 | 0.22 |
| GWAS_mjsw_rs2236996_FGFRL1_ENSG00000127418_1MbTSS.txt | FGFRL1_ENSG00000127418 | 0.00 | 0.39 | 0.00 | 0.61 | 0.00 |
| GWAS_mjsw_rs2236996_TACC3_ENSG00000013810_1MbTSS.txt | TACC3_ENSG00000013810 | 0.00 | 0.00 | 0.00 | 1.00 | 0.00 |
| GWAS_mjsw_rs2236996_UVSSA_ENSG00000163945_1MbTSS.txt | UVSSA_ENSG00000163945 | 0.00 | 0.04 | 0.00 | 0.96 | 0.00 |
| GWAS_mjsw_rs7711053_CCDC125_ENSG00000183323_1MbTSS.txt | CCDC125_ENSG00000183323 | 0.00 | 0.00 | 0.00 | 1.00 | 0.00 |
| GWAS_mjsw_rs7711053_GTF2H2C_ENSG00000183474_1MbTSS.txt | GTF2H2C_ENSG00000183474 | 0.02 | 0.00 | 0.95 | 0.03 | 0.00 |
| GWAS_mjsw_rs7711053_RAD17_ENSG00000152942_1MbTSS.txt | RAD17_ENSG00000152942 | 0.00 | 0.00 | 0.00 | 1.00 | 0.00 |
| GWAS_mjsw_rs17172430_CHCHD2_ENSG00000106153_1MbTSS.txt | CHCHD2_ENSG00000106153 | 0.00 | 0.00 | 0.99 | 0.01 | 0.00 |
| GWAS_mjsw_rs17172430_PSPHP1_ENSG00000226278_1MbTSS.txt | PSPHP1_ENSG00000226278 | 0.00 | 0.00 | 0.15 | 0.85 | 0.00 |
| GWAS_mjsw_rs17172430_PSPH_ENSG00000146733_1MbTSS.txt | PSPH_ENSG00000146733 | 0.00 | 0.00 | 0.18 | 0.82 | 0.00 |
| GWAS_mjsw_rs17172430_ZNF713_ENSG00000178665_1MbTSS.txt | ZNF713_ENSG00000178665 | 0.00 | 0.00 | 0.15 | 0.85 | 0.00 |
| GWAS_mjsw_rs62479589_IRF5_ENSG00000128604_1MbTSS.txt | IRF5_ENSG00000128604 | 0.00 | 0.02 | 0.03 | 0.95 | 0.00 |
| GWAS_mjsw_rs62479589_OPN1SW_ENSG00000128617_1MbTSS.txt | OPN1SW_ENSG00000128617 | 0.00 | 0.00 | 0.00 | 0.09 | 0.90 |
| GWAS_mjsw_rs597974_BRD3_ENSG00000169925_1MbTSS.txt | BRD3_ENSG00000169925 | 0.00 | 0.00 | 0.26 | 0.74 | 0.00 |
| GWAS_mjsw_rs597974_LINC00094_ENSG00000235106_1MbTSS.txt | LINC00094_ENSG00000235106 | 0.00 | 0.00 | 0.26 | 0.74 | 0.00 |
| GWAS_mjsw_rs597974_SURF1_ENSG00000148290_1MbTSS.txt | SURF1_ENSG00000148290 | 0.00 | 0.00 | 0.26 | 0.74 | 0.00 |
| GWAS_mjsw_rs34651525_DKK3_ENSG00000050165_1MbTSS.txt | DKK3_ENSG00000050165 | 0.00 | 0.00 | 0.00 | 1.00 | 0.00 |
| GWAS_mjsw_rs45540840_TRAPPC4_ENSG00000196655_1MbTSS.txt | TRAPPC4_ENSG00000196655 | 0.00 | 0.00 | 0.02 | 0.98 | 0.00 |
| GWAS_mjsw_rs2260671_HELB_ENSG00000127311_1MbTSS.txt | HELB_ENSG00000127311 | 0.00 | 0.00 | 0.00 | 1.00 | 0.00 |
| GWAS_mjsw_rs2260671_WIF1_ENSG00000156076_1MbTSS.txt | WIF1_ENSG00000156076 | 0.00 | 0.02 | 0.00 | 0.98 | 0.00 |
| GWAS_mjsw_rs11857461_AQP9_ENSG00000103569_1MbTSS.txt | AQP9_ENSG00000103569 | 0.00 | 0.07 | 0.00 | 0.93 | 0.00 |
| GWAS_mjsw_rs7179372_SMAD3_ENSG00000166949_1MbTSS.txt | SMAD3_ENSG00000166949 | 0.00 | 0.00 | 0.00 | 1.00 | 0.00 |
| GWAS_mjsw_rs1809360_SMAD3_ENSG00000166949_1MbTSS.txt | SMAD3_ENSG00000166949 | 0.00 | 0.00 | 0.00 | 1.00 | 0.00 |
| GWAS_mjsw_rs8097746_RPL17-C18orf32_ENSG00000215472_1MbTSS.txt | RPL17-C18orf32_ENSG00000215472 | 0.00 | 0.00 | 0.00 | 1.00 | 0.00 |
| GWAS_mjsw_rs34656141_AMH_ENSG00000104899_1MbTSS.txt | AMH_ENSG00000104899 | 0.00 | 0.00 | 0.00 | 1.00 | 0.00 |
| GWAS_mjsw_rs34656141_C19orf24_ENSG00000228300_1MbTSS.txt | C19orf24_ENSG00000228300 | 0.00 | 0.02 | 0.00 | 0.98 | 0.00 |
| GWAS_mjsw_rs34656141_POLR2E_ENSG00000099817_1MbTSS.txt | POLR2E_ENSG00000099817 | 0.00 | 0.00 | 0.98 | 0.01 | 0.00 |
| GWAS_mjsw_rs34656141_ZNF554_ENSG00000172006_1MbTSS.txt | ZNF554_ENSG00000172006 | 0.00 | 0.00 | 0.00 | 1.00 | 0.00 |
| GWAS_mjsw_rs34717890_FKRP_ENSG00000181027_1MbTSS.txt | FKRP_ENSG00000181027 | 0.00 | 0.02 | 0.00 | 0.98 | 0.00 |
| GWAS_mjsw_rs34717890_PNMAL1_ENSG00000182013_1MbTSS.txt | PNMAL1_ENSG00000182013 | 0.00 | 0.00 | 0.00 | 1.00 | 0.00 |
| GWAS_mjsw_rs34717890_VASP_ENSG00000125753_1MbTSS.txt | VASP_ENSG00000125753 | 0.00 | 0.19 | 0.00 | 0.81 | 0.00 |
| Highly degraded (diseased) cartilage |  |  |  |  |  |  |
| GWASInput | eQTLGene | PP.H0.abf | PP.H1.abf | PP.H2.abf | PP.H3.abf | PP.H4.abf |
| GWAS_mjsw_rs823097_DSTYK_ENSG00000133059_1MbTSS.txt | DSTYK_ENSG00000133059 | 0.00 | 0.00 | 0.01 | 0.99 | 0.00 |
| GWAS_mjsw_rs823097_RAB7L1_ENSG00000117280_1MbTSS.txt | RAB7L1_ENSG00000117280 | 0.00 | 0.00 | 0.00 | 0.04 | 0.96 |
| GWAS_mjsw_rs823097_TMEM81_ENSG00000174529_1MbTSS.txt | TMEM81_ENSG00000174529 | 0.00 | 0.03 | 0.01 | 0.96 | 0.00 |
| GWAS_mjsw_rs10933424_TRPM8_ENSG00000144481_1MbTSS.txt | TRPM8_ENSG00000144481 | 0.00 | 0.01 | 0.07 | 0.92 | 0.00 |
| GWAS_mjsw_rs2236996_CRIPAK_ENSG00000179979_1MbTSS.txt | CRIPAK_ENSG00000179979 | 0.00 | 0.00 | 0.00 | 1.00 | 0.00 |
| GWAS_mjsw_rs2236996_TMEM129_ENSG00000168936_1MbTSS.txt | TMEM129_ENSG00000168936 | 0.00 | 0.00 | 0.00 | 1.00 | 0.00 |
| GWAS_mjsw_rs2236996_UVSSA_ENSG00000163945_1MbTSS.txt | UVSSA_ENSG00000163945 | 0.00 | 0.29 | 0.00 | 0.70 | 0.00 |
| GWAS_mjsw_rs7711053_GTF2H2C_ENSG00000183474_1MbTSS.txt | GTF2H2C_ENSG00000183474 | 0.00 | 0.00 | 0.97 | 0.03 | 0.00 |
| GWAS_mjsw_rs7711053_RAD17_ENSG00000152942_1MbTSS.txt | RAD17_ENSG00000152942 | 0.00 | 0.00 | 0.00 | 1.00 | 0.00 |
| GWAS_mjsw_rs10948155_MRPL14_ENSG00000180992_1MbTSS.txt | MRPL14_ENSG00000180992 | 0.00 | 0.36 | 0.00 | 0.64 | 0.00 |
| GWAS_mjsw_rs17172430_CHCHD2_ENSG00000106153_1MbTSS.txt | CHCHD2_ENSG00000106153 | 0.00 | 0.00 | 0.99 | 0.01 | 0.00 |
| GWAS_mjsw_rs17172430_PSPHP1_ENSG00000226278_1MbTSS.txt | PSPHP1_ENSG00000226278 | 0.00 | 0.00 | 0.15 | 0.85 | 0.00 |
| GWAS_mjsw_rs17172430_PSPH_ENSG00000146733_1MbTSS.txt | PSPH_ENSG00000146733 | 0.00 | 0.00 | 0.18 | 0.82 | 0.00 |
| GWAS_mjsw_rs17172430_ZNF713_ENSG00000178665_1MbTSS.txt | ZNF713_ENSG00000178665 | 0.00 | 0.00 | 0.15 | 0.85 | 0.00 |
| GWAS_mjsw_rs62479589_IRF5_ENSG00000128604_1MbTSS.txt | IRF5_ENSG00000128604 | 0.00 | 0.00 | 0.03 | 0.97 | 0.00 |
| GWAS_mjsw_rs62479589_OPN1SW_ENSG00000128617_1MbTSS.txt | OPN1SW_ENSG00000128617 | 0.00 | 0.00 | 0.00 | 0.03 | 0.97 |
| GWAS_mjsw_rs4744313_PTPDC1_ENSG00000158079_1MbTSS.txt | PTPDC1_ENSG00000158079 | 0.00 | 0.01 | 0.01 | 0.98 | 0.00 |
| GWAS_mjsw_rs4979342_DFNB31_ENSG00000095397_1MbTSS.txt | DFNB31_ENSG00000095397 | 0.00 | 0.00 | 0.00 | 1.00 | 0.00 |
| GWAS_mjsw_rs597974_BRD3_ENSG00000169925_1MbTSS.txt | BRD3_ENSG00000169925 | 0.00 | 0.01 | 0.25 | 0.73 | 0.00 |
| GWAS_mjsw_rs597974_SURF1_ENSG00000148290_1MbTSS.txt | SURF1_ENSG00000148290 | 0.00 | 0.00 | 0.26 | 0.74 | 0.00 |
| GWAS_mjsw_rs597974_SURF6_ENSG00000148296_1MbTSS.txt | SURF6_ENSG00000148296 | 0.00 | 0.00 | 0.26 | 0.73 | 0.00 |
| GWAS_mjsw_rs34651525_DKK3_ENSG00000050165_1MbTSS.txt | DKK3_ENSG00000050165 | 0.00 | 0.00 | 0.00 | 1.00 | 0.00 |
| GWAS_mjsw_rs45540840_TRAPPC4_ENSG00000196655_1MbTSS.txt | TRAPPC4_ENSG00000196655 | 0.00 | 0.00 | 0.02 | 0.98 | 0.00 |
| GWAS_mjsw_rs2260671_GNS_ENSG00000135677_1MbTSS.txt | GNS_ENSG00000135677 | 0.23 | 0.02 | 0.68 | 0.06 | 0.01 |
| GWAS_mjsw_rs2260671_HELB_ENSG00000127311_1MbTSS.txt | HELB_ENSG00000127311 | 0.00 | 0.00 | 0.00 | 1.00 | 0.00 |
| GWAS_mjsw_rs2260671_TMBIM4_ENSG00000155957_1MbTSS.txt | TMBIM4_ENSG00000155957 | 0.00 | 0.01 | 0.00 | 0.99 | 0.00 |
| GWAS_mjsw_rs7179372_DIS3L_ENSG00000166938_1MbTSS.txt | DIS3L_ENSG00000166938 | 0.00 | 0.02 | 0.00 | 0.98 | 0.00 |
| GWAS_mjsw_rs7179372_MAP2K5_ENSG00000137764_1MbTSS.txt | MAP2K5_ENSG00000137764 | 0.00 | 0.02 | 0.00 | 0.98 | 0.00 |
| GWAS_mjsw_rs7179372_SMAD3_ENSG00000166949_1MbTSS.txt | SMAD3_ENSG00000166949 | 0.00 | 0.00 | 0.00 | 1.00 | 0.00 |
| GWAS_mjsw_rs1809360_MAP2K5_ENSG00000137764_1MbTSS.txt | MAP2K5_ENSG00000137764 | 0.00 | 0.02 | 0.00 | 0.98 | 0.00 |
| GWAS_mjsw_rs1809360_SMAD3_ENSG00000166949_1MbTSS.txt | SMAD3_ENSG00000166949 | 0.00 | 0.00 | 0.00 | 1.00 | 0.00 |
| GWAS_mjsw_rs62070652_EVI2A_ENSG00000126860_1MbTSS.txt | EVI2A_ENSG00000126860 | 0.00 | 0.00 | 0.00 | 1.00 | 0.00 |
| GWAS_mjsw_rs8097746_RPL17-C18orf32_ENSG00000215472_1MbTSS.txt | RPL17-C18orf32_ENSG00000215472 | 0.00 | 0.00 | 0.00 | 1.00 | 0.00 |
| GWAS_mjsw_rs34656141_AES_ENSG00000104964_1MbTSS.txt | AES_ENSG00000104964 | 0.00 | 0.14 | 0.00 | 0.86 | 0.00 |
| GWAS_mjsw_rs34656141_AMH_ENSG00000104899_1MbTSS.txt | AMH_ENSG00000104899 | 0.00 | 0.00 | 0.00 | 1.00 | 0.00 |
| GWAS_mjsw_rs34656141_C19orf24_ENSG00000228300_1MbTSS.txt | C19orf24_ENSG00000228300 | 0.00 | 0.01 | 0.00 | 0.99 | 0.00 |
| GWAS_mjsw_rs34656141_MUM1_ENSG00000160953_1MbTSS.txt | MUM1_ENSG00000160953 | 0.00 | 0.02 | 0.00 | 0.98 | 0.00 |
| GWAS_mjsw_rs34656141_PLEKHJ1_ENSG00000104886_1MbTSS.txt | PLEKHJ1_ENSG00000104886 | 0.00 | 0.13 | 0.00 | 0.87 | 0.00 |
| GWAS_mjsw_rs34656141_POLR2E_ENSG00000099817_1MbTSS.txt | POLR2E_ENSG00000099817 | 0.00 | 0.00 | 0.98 | 0.01 | 0.00 |
| GWAS_mjsw_rs34656141_ZNF554_ENSG00000172006_1MbTSS.txt | ZNF554_ENSG00000172006 | 0.00 | 0.00 | 0.00 | 1.00 | 0.00 |
| GWAS_mjsw_rs34656141_ZNF77_ENSG00000175691_1MbTSS.txt | ZNF77_ENSG00000175691 | 0.00 | 0.00 | 0.00 | 1.00 | 0.00 |
| GWAS_mjsw_rs34717890_PNMAL1_ENSG00000182013_1MbTSS.txt | PNMAL1_ENSG00000182013 | 0.00 | 0.00 | 0.00 | 1.00 | 0.00 |
| GWAS_mjsw_rs34717890_PPP5C_ENSG00000011485_1MbTSS.txt | PPP5C_ENSG00000011485 | 0.00 | 0.39 | 0.00 | 0.60 | 0.00 |
| GWAS_mjsw_rs7571789_SNRNP27_ENSG00000124380_1MbTSS.txt | SNRNP27_ENSG00000124380 | 0.00 | 0.07 | 0.00 | 0.93 | 0.00 |
| GWAS_mjsw_rs10962293_TTC39B_ENSG00000155158_1MbTSS.txt | TTC39B_ENSG00000155158 | 0.00 | 0.02 | 0.05 | 0.94 | 0.00 |
| GWAS_mjsw_rs34949187_ABHD2_ENSG00000140526_1MbTSS.txt | ABHD2_ENSG00000140526 | 0.00 | 0.15 | 0.00 | 0.84 | 0.01 |
| GWAS_mjsw_rs34949187_AP3S2_ENSG00000157823_1MbTSS.txt | AP3S2_ENSG00000157823 | 0.00 | 0.00 | 0.99 | 0.01 | 0.00 |
| GWAS_mjsw_rs34949187_C15orf38-AP3S2_ENSG00000250021_1MbTSS.txt | C15orf38-AP3S2_ENSG00000250021 | 0.00 | 0.00 | 0.99 | 0.01 | 0.00 |
| GWAS_mjsw_rs34949187_C15orf38_ENSG00000242498_1MbTSS.txt | C15orf38_ENSG00000242498 | 0.01 | 0.00 | 0.98 | 0.01 | 0.00 |
| GWAS_mjsw_rs34949187_ISG20_ENSG00000172183_1MbTSS.txt | ISG20_ENSG00000172183 | 0.00 | 0.30 | 0.00 | 0.70 | 0.00 |
| GWAS_mjsw_rs2716212_ABCA5_ENSG00000154265_1MbTSS.txt | ABCA5_ENSG00000154265 | 0.00 | 0.00 | 0.04 | 0.96 | 0.00 |
| GWAS_mjsw_rs61648765_PNMAL1_ENSG00000182013_1MbTSS.txt | PNMAL1_ENSG00000182013 | 0.00 | 0.00 | 0.00 | 1.00 | 0.00 |
| GWAS_mjsw_rs61648765_PPP5C_ENSG00000011485_1MbTSS.txt | PPP5C_ENSG00000011485 | 0.00 | 0.39 | 0.00 | 0.60 | 0.00 |
| Synovium |  |  |  |  |  |  |
| GWASInput | eQTLGene | PP.H0.abf | PP.H1.abf | PP.H2.abf | PP.H3.abf | PP.H4.abf |
| GWAS_mjsw_rs823097_DSTYK_ENSG00000133059_1MbTSS.txt | DSTYK_ENSG00000133059 | 0.00 | 0.00 | 0.01 | 0.99 | 0.00 |
| GWAS_mjsw_rs823097_NUAK2_ENSG00000163545_1MbTSS.txt | NUAK2_ENSG00000163545 | 0.00 | 0.00 | 0.01 | 0.99 | 0.00 |
| GWAS_mjsw_rs823097_RAB7L1_ENSG00000117280_1MbTSS.txt | RAB7L1_ENSG00000117280 | 0.00 | 0.00 | 0.01 | 0.94 | 0.05 |
| GWAS_mjsw_rs823097_RBBP5_ENSG00000117222_1MbTSS.txt | RBBP5_ENSG00000117222 | 0.00 | 0.13 | 0.01 | 0.86 | 0.00 |
| GWAS_mjsw_rs823097_TMCC2_ENSG00000133069_1MbTSS.txt | TMCC2_ENSG00000133069 | 0.00 | 0.00 | 0.01 | 0.99 | 0.00 |
| GWAS_mjsw_rs10933424_NGEF_ENSG00000066248_1MbTSS.txt | NGEF_ENSG00000066248 | 0.00 | 0.00 | 0.00 | 1.00 | 0.00 |
| GWAS_mjsw_rs2236996_CRIPAK_ENSG00000179979_1MbTSS.txt | CRIPAK_ENSG00000179979 | 0.00 | 0.00 | 0.00 | 1.00 | 0.00 |
| GWAS_mjsw_rs2236996_TNIP2_ENSG00000168884_1MbTSS.txt | TNIP2_ENSG00000168884 | 0.00 | 0.15 | 0.00 | 0.84 | 0.00 |
| GWAS_mjsw_rs2236996_UVSSA_ENSG00000163945_1MbTSS.txt | UVSSA_ENSG00000163945 | 0.00 | 0.01 | 0.00 | 0.99 | 0.00 |
| GWAS_mjsw_rs2236996_ZFYVE28_ENSG00000159733_1MbTSS.txt | ZFYVE28_ENSG00000159733 | 0.00 | 0.00 | 0.00 | 1.00 | 0.00 |
| GWAS_mjsw_rs7711053_RAD17_ENSG00000152942_1MbTSS.txt | RAD17_ENSG00000152942 | 0.00 | 0.00 | 0.00 | 1.00 | 0.00 |
| GWAS_mjsw_rs17172430_PSPHP1_ENSG00000226278_1MbTSS.txt | PSPHP1_ENSG00000226278 | 0.00 | 0.00 | 0.15 | 0.85 | 0.00 |
| GWAS_mjsw_rs17172430_PSPH_ENSG00000146733_1MbTSS.txt | PSPH_ENSG00000146733 | 0.00 | 0.00 | 0.18 | 0.82 | 0.00 |
| GWAS_mjsw_rs4979342_ATP6V1G1_ENSG00000136888_1MbTSS.txt | ATP6V1G1_ENSG00000136888 | 0.00 | 0.00 | 0.00 | 1.00 | 0.00 |
| GWAS_mjsw_rs4979342_DFNB31_ENSG00000095397_1MbTSS.txt | DFNB31_ENSG00000095397 | 0.00 | 0.02 | 0.00 | 0.98 | 0.00 |
| GWAS_mjsw_rs4979342_PRPF4_ENSG00000136875_1MbTSS.txt | PRPF4_ENSG00000136875 | 0.00 | 0.00 | 0.00 | 1.00 | 0.00 |
| GWAS_mjsw_rs597974_SURF1_ENSG00000148290_1MbTSS.txt | SURF1_ENSG00000148290 | 0.00 | 0.00 | 0.26 | 0.74 | 0.00 |
| GWAS_mjsw_rs597974_SURF6_ENSG00000148296_1MbTSS.txt | SURF6_ENSG00000148296 | 0.02 | 0.06 | 0.24 | 0.68 | 0.00 |
| GWAS_mjsw_rs34651525_DKK3_ENSG00000050165_1MbTSS.txt | DKK3_ENSG00000050165 | 0.00 | 0.01 | 0.00 | 0.99 | 0.00 |
| GWAS_mjsw_rs2260671_HELB_ENSG00000127311_1MbTSS.txt | HELB_ENSG00000127311 | 0.00 | 0.01 | 0.00 | 0.99 | 0.00 |
| GWAS_mjsw_rs2260671_WIF1_ENSG00000156076_1MbTSS.txt | WIF1_ENSG00000156076 | 0.00 | 0.00 | 0.00 | 1.00 | 0.00 |
| GWAS_mjsw_rs7179372_MAP2K5_ENSG00000137764_1MbTSS.txt | MAP2K5_ENSG00000137764 | 0.00 | 0.00 | 0.00 | 1.00 | 0.00 |
| GWAS_mjsw_rs1809360_MAP2K5_ENSG00000137764_1MbTSS.txt | MAP2K5_ENSG00000137764 | 0.00 | 0.00 | 0.00 | 1.00 | 0.00 |
| GWAS_mjsw_rs62070652_AK4P1_ENSG00000263535_1MbTSS.txt | AK4P1_ENSG00000263535 | 0.00 | 0.02 | 0.00 | 0.98 | 0.00 |
| GWAS_mjsw_rs62070652_LRRC37BP1_ENSG00000250462_1MbTSS.txt | LRRC37BP1_ENSG00000250462 | 0.00 | 0.00 | 0.00 | 1.00 | 0.00 |
| GWAS_mjsw_rs8097746_RPL17-C18orf32_ENSG00000215472_1MbTSS.txt | RPL17-C18orf32_ENSG00000215472 | 0.00 | 0.00 | 0.00 | 1.00 | 0.00 |
| GWAS_mjsw_rs34656141_AMH_ENSG00000104899_1MbTSS.txt | AMH_ENSG00000104899 | 0.00 | 0.00 | 0.00 | 1.00 | 0.00 |
| GWAS_mjsw_rs34656141_C19orf24_ENSG00000228300_1MbTSS.txt | C19orf24_ENSG00000228300 | 0.00 | 0.00 | 0.00 | 1.00 | 0.00 |
| GWAS_mjsw_rs34656141_LSM7_ENSG00000130332_1MbTSS.txt | LSM7_ENSG00000130332 | 0.00 | 0.00 | 0.00 | 1.00 | 0.00 |
| GWAS_mjsw_rs34656141_ZNF554_ENSG00000172006_1MbTSS.txt | ZNF554_ENSG00000172006 | 0.00 | 0.00 | 0.00 | 1.00 | 0.00 |
| GWAS_mjsw_rs34717890_FKRP_ENSG00000181027_1MbTSS.txt | FKRP_ENSG00000181027 | 0.00 | 0.01 | 0.00 | 0.98 | 0.00 |
| GWAS_mjsw_rs34717890_PNMAL1_ENSG00000182013_1MbTSS.txt | PNMAL1_ENSG00000182013 | 0.00 | 0.00 | 0.00 | 1.00 | 0.00 |
| GWAS_mjsw_rs34717890_STRN4_ENSG00000090372_1MbTSS.txt | STRN4_ENSG00000090372 | 0.00 | 0.01 | 0.00 | 0.99 | 0.00 |
| GWAS_mjsw_rs7633464_DCBLD2_ENSG00000057019_1MbTSS.txt | DCBLD2_ENSG00000057019 | 0.00 | 0.01 | 0.00 | 0.99 | 0.00 |
| GWAS_mjsw_rs7633464_MINA_ENSG00000170854_1MbTSS.txt | MINA_ENSG00000170854 | 0.00 | 0.00 | 0.02 | 0.98 | 0.00 |
| GWAS_mjsw_rs10962293_TTC39B_ENSG00000155158_1MbTSS.txt | TTC39B_ENSG00000155158 | 0.00 | 0.00 | 0.05 | 0.95 | 0.00 |
| GWAS_mjsw_rs34949187_C15orf38-AP3S2_ENSG00000250021_1MbTSS.txt | C15orf38-AP3S2_ENSG00000250021 | 0.02 | 0.00 | 0.97 | 0.01 | 0.00 |
| GWAS_mjsw_rs34949187_ISG20_ENSG00000172183_1MbTSS.txt | ISG20_ENSG00000172183 | 0.00 | 0.00 | 0.00 | 1.00 | 0.00 |

### Supplementary Table 8. PANTHER Output

| GO biological process complete | Homo sapiens - REFLIST (20589) | Client Text Box Input (39) | Client Text Box Input (expected) | Client Text Box Input (over/under) | Client Text Box Input (fold Enrichment) | Client Text Box Input (P-value) |
| --- | --- | --- | --- | --- | --- | --- |
| skeletal system development (GO:0001501) | 515 | **8** | 0.98 | + | 8.2 | 4.63E-02 |
| Unclassified (UNCLASSIFIED) | 2725 | 1 | 5.16 | - | 0.19 | 0.00E+00 |
| Gnene ID | **Mapped IDs** | **Gene Name, Symbol, Persistent ID, Orthologs** | **PANTHER Family/Subfamily** | **PANTHER Protein Class** | **Species** | **MR Cluster** |
| HUMAN\|HGNC=22986\|UniProtKB=Q8IZC6 | ENSG00000196739 | Collagen alpha-1(XXVII) chain;COL27A1;PTN002513112;orthologs | COLLAGEN ALPHA-1(XXVII) CHAIN (PTHR24023:SF844) | extracellular matrix structural protein(PC00103) | Homo sapiens | 1 |
| HUMAN\|HGNC=21317\|UniProtKB=Q7RTS9 | ENSG00000141627 | Dymeclin;DYM;PTN002490062;orthologs | DYMECLIN (PTHR12895:SF9) |  | Homo sapiens | 3 |
| HUMAN\|HGNC=319\|UniProtKB=P16112 | ENSG00000157766 | Aggrecan core protein;ACAN;PTN002504044;orthologs | AGGRECAN CORE PROTEIN (PTHR22804:SF42) | extracellular matrix glycoprotein(PC00100) | Homo sapiens | 2 |
| HUMAN\|HGNC=5009\|UniProtKB=P52926 | ENSG00000149948 | High mobility group protein HMGI-C;HMGA2;PTN002511483;orthologs | HIGH MOBILITY GROUP PROTEIN HMGI-C (PTHR23341:SF4) | endodeoxyribonuclease(PC00093) | Homo sapiens | 3 |
| HUMAN\|HGNC=14866\|UniProtKB=Q96QV1 | ENSG00000164161 | Hedgehog-interacting protein;HHIP;PTN002499831;orthologs | HEDGEHOG-INTERACTING PROTEIN (PTHR19328:SF27) | protein-binding activity modulator(PC00095) | Homo sapiens | 3 |
| HUMAN\|HGNC=7866\|UniProtKB=Q13253 | ENSG00000183691 | Noggin;NOG;PTN002471070;orthologs | NOGGIN (PTHR10494:SF5) | intercellular signal molecule(PC00207) | Homo sapiens | 2 |
| HUMAN\|HGNC=6846\|UniProtKB=P52564 | ENSG00000108984 | Dual specificity mitogen-activated protein kinase kinase 6;MAP2K6;PTN002519608;orthologs | DUAL SPECIFICITY MITOGEN-ACTIVATED PROTEIN KINASE KINASE 6 (PTHR48013:SF12) | non-receptor serine/threonine protein kinase(PC00167) | Homo sapiens | 2 |
| HUMAN\|HGNC=1073\|UniProtKB=P22004 | ENSG00000153162 | Bone morphogenetic protein 6;BMP6;PTN002483823;orthologs | BONE MORPHOGENETIC PROTEIN 6 (PTHR11848:SF137) | growth factor(PC00112) | Homo sapiens | 2 |

Analysis Type:	PANTHER Overrepresentation Test (Released 20221013)

Annotation Version and Release Date:	GO Ontology database DOI: 10.5281/zenodo.6799722 Released 2022-07-01

Analyzed List:	Client Text Box Input (Homo sapiens)

Reference List:	Homo sapiens (all genes in database)

Test Type:	FISHER

Correction:	BONFERRONI

Bonferroni count:	9290

### Supplementary Table 9. FUMA Gene2Func output

| Category | GeneSet | N_genes | N_overlap | p | adjP | genes |
| --- | --- | --- | --- | --- | --- | --- |
| GO_bp | GO_POSITIVE_REGULATION_OF_BIOSYNTHETIC_PROCESS | 1966 | 14 | 7.61E-09 | 5.60E-05 | NUCKS1:EPC1:TEAD1:HMGA2:SMAD6:ATAD5:NOG:MAP2K6:FOXA3:PIK3R1:RGMB:BMP6:SUPT3H:EGFR |
| GO_bp | GO_ENZYME_LINKED_RECEPTOR_PROTEIN_SIGNALING_PATHWAY | 1034 | 11 | 7.89E-09 | 5.80E-05 | NUCKS1:SMAD6:SKOR1:NOG:TGFA:NGEF:HHIP:PIK3R1:RGMB:BMP6:EGFR |
| GO_bp | GO_POSITIVE_REGULATION_OF_RNA_BIOSYNTHETIC_PROCESS | 1592 | 12 | 6.48E-08 | 0.000476 | NUCKS1:EPC1:TEAD1:HMGA2:SMAD6:NOG:FOXA3:PIK3R1:RGMB:BMP6:SUPT3H:EGFR |
| GO_bp | GO_POSITIVE_REGULATION_OF_GENE_EXPRESSION | 1955 | 13 | 6.86E-08 | 0.000504 | NUCKS1:EPC1:TEAD1:HMGA2:ALDH1A2:SMAD6:NOG:FOXA3:PIK3R1:RGMB:BMP6:SUPT3H:EGFR |
| GO_bp | GO_REGULATION_OF_CELL_DEATH | 1697 | 12 | 1.30E-07 | 0.000954 | HMGA2:ALDH1A2:SMAD6:ATAD5:NOG:MAP2K6:NGEF:HHIP:PIK3R1:BMP6:TIAM2:EGFR |
| GO_bp | GO_POSITIVE_REGULATION_OF_TRANSCRIPTION_BY_RNA_POLYMERASE_II | 1178 | 10 | 3.47E-07 | 0.002548 | NUCKS1:EPC1:TEAD1:HMGA2:SMAD6:NOG:FOXA3:PIK3R1:BMP6:EGFR |
| GO_bp | GO_APOPTOTIC_PROCESS | 1956 | 12 | 5.97E-07 | 0.004385 | HMGA2:ALDH1A2:SMAD6:ATAD5:NOG:MAP2K6:NGEF:HHIP:PIK3R1:BMP6:TIAM2:EGFR |
| GO_bp | GO_RESPONSE_TO_ENDOGENOUS_STIMULUS | 1634 | 11 | 8.00E-07 | 0.005876 | NUCKS1:ALDH1A2:SMAD6:SKOR1:NOG:HHIP:PIK3R1:RGMB:BMP6:EGFR:PAPPA |
| GO_bp | GO_RESPONSE_TO_BMP | 165 | 5 | 9.51E-07 | 0.006989 | SMAD6:SKOR1:NOG:RGMB:BMP6 |
| GO_bp | GO_ANIMAL_ORGAN_MORPHOGENESIS | 1030 | 9 | 1.17E-06 | 0.008597 | ALDH1A2:SMAD6:ACAN:NOG:HHIP:BMP6:EGFR:FANCC:COL27A1 |
| GO_bp | GO_SKELETAL_SYSTEM_DEVELOPMENT | 517 | 7 | 1.22E-06 | 0.008954 | HMGA2:ACAN:NOG:DYM:HHIP:BMP6:COL27A1 |
| GO_bp | GO_CARTILAGE_DEVELOPMENT | 207 | 5 | 2.90E-06 | 0.021282 | HMGA2:ACAN:NOG:BMP6:COL27A1 |
| GO_bp | GO_SKELETAL_SYSTEM_MORPHOGENESIS | 238 | 5 | 5.71E-06 | 0.042002 | ACAN:NOG:HHIP:BMP6:COL27A1 |
| GWAScatalog | Height | 898 | 16 | 3.81E-16 | 6.92E-13 | TEAD1:HMGA2:ALDH1A2:ACAN:ATAD5:NOG:DYM:AP3D1:RAB28:HHIP:BMP6:SUPT3H:PTPDC1:COL15A1:COL27A1:PAPPA |
| GWAScatalog | Heel bone mineral density | 834 | 12 | 4.54E-11 | 8.24E-08 | TEAD1:PHLDB1:HMGA2:TGFA:NGEF:RAB28:HHIP:SUPT3H:TIAM2:PTPDC1:PAPPA:ASTN2 |
| GWAScatalog | Hip minimal joint space width | 10 | 4 | 2.43E-10 | 4.41E-07 | TGFA:SLBP:PIK3R1:SUPT3H |
| GWAScatalog | Hip circumference adjusted for BMI | 218 | 7 | 3.44E-09 | 6.25E-06 | HMGA2:ACAN:ATAD5:DYM:SLBP:HHIP:BMP6 |
| GWAScatalog | Birth weight | 278 | 7 | 1.84E-08 | 3.34E-05 | HMGA2:ATAD5:HHIP:PIK3R1:PTPDC1:FANCC:PAPPA |
| GWAScatalog | Infant length | 14 | 3 | 4.21E-07 | 0.000764 | HMGA2:ACAN:HHIP |
| GWAScatalog | Offspring birth weight | 153 | 5 | 6.55E-07 | 0.001189 | HMGA2:HHIP:PIK3R1:FANCC:COL27A1 |
| GWAScatalog | Chronic obstructive pulmonary disease | 144 | 4 | 1.79E-05 | 0.032508 | HHIP:BMP6:COL15A1:ASTN2 |
| Chemical_and_Genetic_pertubation | MEISSNER_BRAIN_HCP_WITH_H3K4ME3_AND_H3K27ME3 | 1055 | 9 | 1.43E-06 | 0.004707 | ALDH1A2:SMAD6:NOG:HHIP:BMP6:EGFR:COL15A1:COL27A1:PAPPA |
| Chemical_and_Genetic_pertubation | BOQUEST_STEM_CELL_CULTURED_VS_FRESH_UP | 426 | 6 | 6.09E-06 | 0.020114 | ACAN:PIK3R1:EGFR:CALU:COL15A1:PAPPA |
| Chemical_and_Genetic_pertubation | BENPORATH_SUZ12_TARGETS | 1031 | 8 | 1.19E-05 | 0.039459 | PHLDB1:SMAD6:ACAN:NOG:HHIP:COL27A1:PAPPA:ASTN2 |
| Chemical_and_Genetic_pertubation | BENPORATH_EED_TARGETS | 1057 | 8 | 1.43E-05 | 0.047234 | NOG:TGFA:HHIP:BMP6:EGFR:COL27A1:PAPPA:ASTN2 |
| GO_cc | GO_EXTRACELLULAR_MATRIX_COMPONENT | 49 | 3 | 2.08E-05 | 0.020786 | ACAN:COL15A1:COL27A1 |
| Immunologic_signatures | GSE39152_CD103_NEG_VS_POS_MEMORY_CD8_TCELL_UP | 200 | 5 | 2.45E-06 | 0.011922 | PHLDB1:HMGA2:ALDH1A2:AP3D1:SURF6 |
| BioCarta | BIOCARTA_HCMV_PATHWAY | 17 | 2 | 0.000153 | 0.044295 | MAP2K6:PIK3R1 |
| BioCarta | BIOCARTA_PTEN_PATHWAY | 18 | 2 | 0.000172 | 0.049798 | PIK3R1:EGFR |
| Wikipathways | ESC Pluripotency Pathways | 117 | 4 | 7.89E-06 | 0.004286 | SMAD6:NOG:MAP2K6:EGFR |
| Wikipathways | Mesodermal Commitment Pathway | 156 | 4 | 2.45E-05 | 0.013315 | TEAD1:HMGA2:SMAD6:NOG |
| Wikipathways | Non-small cell lung cancer | 72 | 3 | 6.61E-05 | 0.035879 | TGFA:PIK3R1:EGFR |
| Wikipathways | Bone Morphogenic Protein (BMP) Signalling and Regulation | 12 | 2 | 7.46E-05 | 0.040527 | SMAD6:NOG |
| Curated_gene_sets | PID_BMP_PATHWAY | 42 | 4 | 1.26E-07 | 0.000695 | SMAD6:NOG:RGMB:BMP6 |
| Curated_gene_sets | REACTOME_GAB1_SIGNALOSOME | 17 | 3 | 7.85E-07 | 0.004318 | TGFA:PIK3R1:EGFR |
| Curated_gene_sets | MEISSNER_BRAIN_HCP_WITH_H3K4ME3_AND_H3K27ME3 | 1055 | 9 | 1.43E-06 | 0.007841 | ALDH1A2:SMAD6:NOG:HHIP:BMP6:EGFR:COL15A1:COL27A1:PAPPA |
| Curated_gene_sets | REACTOME_SIGNALING_BY_EGFR_IN_CANCER | 24 | 3 | 2.32E-06 | 0.012786 | TGFA:PIK3R1:EGFR |
| Curated_gene_sets | BOQUEST_STEM_CELL_CULTURED_VS_FRESH_UP | 426 | 6 | 6.09E-06 | 0.033509 | ACAN:PIK3R1:EGFR:CALU:COL15A1:PAPPA |
| Oncogenic_signatures | ATF2_S_UP.V1_DN | 185 | 4 | 4.77E-05 | 0.009018 | SMAD6:MN1:BMP6:COL15A1 |
| KEGG | KEGG_NON_SMALL_CELL_LUNG_CANCER | 54 | 3 | 2.79E-05 | 0.005181 | TGFA:PIK3R1:EGFR |
| KEGG | KEGG_GLIOMA | 65 | 3 | 4.86E-05 | 0.009048 | TGFA:PIK3R1:EGFR |
| KEGG | KEGG_PANCREATIC_CANCER | 70 | 3 | 6.07E-05 | 0.011297 | TGFA:PIK3R1:EGFR |
| KEGG | KEGG_TGF_BETA_SIGNALING_PATHWAY | 85 | 3 | 0.000108 | 0.020157 | SMAD6:NOG:BMP6 |
| KEGG | KEGG_ERBB_SIGNALING_PATHWAY | 87 | 3 | 0.000116 | 0.0216 | TGFA:PIK3R1:EGFR |
| KEGG | KEGG_PROSTATE_CANCER | 89 | 3 | 0.000124 | 0.023108 | TGFA:PIK3R1:EGFR |
| microRNA_targets | TATTATA_MIR374 | 285 | 5 | 1.37E-05 | 0.003018 | SMAD6:NOG:MAP2K6:TGFA:PAPPA |
| microRNA_targets | CAGTATT_MIR200B_MIR200C_MIR429 | 465 | 5 | 0.000139 | 0.030728 | TEAD1:PHLDB1:NOG:NGEF:CALU |
| TF_targets | CEBP_Q3 | 259 | 5 | 8.61E-06 | 0.005251 | ALDH1A2:MAP2K6:MN1:HHIP:SUPT3H |
| TF_targets | CEBPB_02 | 268 | 5 | 1.02E-05 | 0.006194 | PHLDB1:ALDH1A2:SMAD6:ACAN:MAP2K6 |
| TF_targets | WTTGKCTG_UNKNOWN | 522 | 6 | 1.93E-05 | 0.011743 | HMGA2:SMAD6:MN1:PIK3R1:BMP6:CALU |
| Reactome | REACTOME_GAB1_SIGNALOSOME | 17 | 3 | 7.85E-07 | 0.001177 | TGFA:PIK3R1:EGFR |
| Reactome | REACTOME_SIGNALING_BY_EGFR_IN_CANCER | 24 | 3 | 2.32E-06 | 0.003484 | TGFA:PIK3R1:EGFR |
| Reactome | REACTOME_SIGNALING_BY_EGFR | 49 | 3 | 2.08E-05 | 0.031128 | TGFA:PIK3R1:EGFR |
| Canonical_Pathways | PID_BMP_PATHWAY | 42 | 4 | 1.26E-07 | 0.000278 | SMAD6:NOG:RGMB:BMP6 |
| Canonical_Pathways | REACTOME_GAB1_SIGNALOSOME | 17 | 3 | 7.85E-07 | 0.001726 | TGFA:PIK3R1:EGFR |
| Canonical_Pathways | REACTOME_SIGNALING_BY_EGFR_IN_CANCER | 24 | 3 | 2.32E-06 | 0.005111 | TGFA:PIK3R1:EGFR |
| Canonical_Pathways | REACTOME_SIGNALING_BY_EGFR | 49 | 3 | 2.08E-05 | 0.045664 | TGFA:PIK3R1:EGFR |

FUMA GENE2FUNC output

######################

\# GS.tx

t# Results of gene set enrichment analysis.

\# Only significant gene sets are included in this file.

######################

Category : One of the category from MsigDB

GeneSet : Name of gene set as provided by MsigDB

N_genes : Number of genes in a gene set

N_overlap : Number of input genes overlapping with the gene set

p : Hypergeometric test (upper tail) P-value

adjP : Adjusted P-value (user defined method)

genes : Genes overlapping with the gene set

link : Link to the MsigDB page if available
